# Full stack academic publishing in seizures with artificial intelligence: a cautionary tale

**DOI:** 10.64898/2026.10.07.26363237

**Authors:** Wesley T. Kerr, Jacob Pellinen, Katherine N. McFarlane, Daniel M. Goldenholz

## Abstract

**Significance:** There is potential for generative artificial intelligence (AI) to perform “full stack” academic publishing, including comprehensive manuscript fabrication, “peer”-review, and editorial decision making. This study evaluated the benefits and cautions in using AI for full stack academic publishing in clinical research about seizures.

**Methods:** Three epileptologists each selected 1 published manuscript in *Epilepsia* and produced an AI-fabricated manuscript about seizures. AI tools were then used to perform “peer”-review and make editorial decisions with minimal human supervision. The rates of identifying AI-fabricated manuscripts and editorial decisions among the epileptologists was analyzed, and epileptologists’ observations on how AI differed from typical human “peer”-review and editorial decisions was collected.

**Results:** Four of 6 (67%) pairs of AI-fabricated and human-generated manuscripts were correctly identified. Only 1 AI-fabricated manuscript was editorially rejected, whereas another had a subtle indicator of fabrication. When compared to human content, AI generated content was comprehensive, detailed, and based on the highest standard of academic publication. However, generated content was more verbose, used more structured responses, and more critical feedback.

**Conclusions:** These results demonstrate that the quality of AI-fabricated manuscripts about clinical research in seizures was sufficiently high that the current methods of peer and editorial review could not easily distinguish fabricated content compared to human-written content. Moreover, AI review and editorial decisions were high quality and appeared reasonable in many cases. AI tools have matured to the point that there should be thoughtful policies that permit AI assistance but prohibit insufficiently supervised autonomous scientific submissions, reviews, and editorial decisions.

**Highlights:**

- The maturity of AI tools to assist in manuscript generation, peer-review, and editorial decisions comes with cautions.
- Only 1 of 3 artificial intelligence (AI) fabricated manuscripts about seizures were reliably identified and rejected.
- This study describes characteristics of AI-generated peer-review and editorial comments that had insufficient human oversight.

## 1. Introduction

Generative artificial intelligence (AI) tools, particularly large language models (LLMs), are rapidly reshaping the landscape of academic publishing from manuscript generation, peer-review, and editorial decision making.^1–3^ These technologies also have transformative potential to democratize academic publishing by reducing communication challenges for investigators less familiar with the steps of academic publishing, including increasing access for low and middle-income investigators.^4,5^ In addition, the volume of scientific articles submitted for peer review to academic journals has progressively increased, which substantially increases the burden of peer-reviewers and editors to perform high quality and efficient evaluation of novel manuscripts.^6,7^ By reducing the barriers to creation of manuscripts, AI has increased those challenges as well as enabled greater sophistication of fabricated works.^8,9^

The potential benefits and cautions of utilizing AI for scientific writing have been well described. In addition to reducing the time required to write a manuscript, AI-assisted scientific works can have improved readability scores and improved reference to relevant high-impact manuscripts.^5,10^ However, AI typically has less expertise in the specific field of the work than expert authors, which results in focus on more general and vague insights, as compared to the nuanced contributions that likely are necessary to create meaningful change in the scientific literature.^11,12^ The importance of expert insights is particularly important in the clinically and technically complex field of seizures where there are rapidly evolving definitions of seizures, identification of precision medicine approaches for newly identified genetic epilepsies, and technological advancements in the interpretation of multimodal data (e.g., electroencephalography, wearables, implantable devices, neuroimaging, and clinical data). Because AI is trained on historical knowledge, it can struggle to keep up with these rapidly developing fields.^13^ This work also may be broadly applicable to academic publishing outside of seizures.

Editors and peer-reviewers play key roles in assessing manuscript quality and appropriateness. While there are recognized limitations in the peer-review process without AI, there is great heterogeneity in the AI use policies of journals, including *Epilepsia*, where 59% explicitly prohibit AI use primarily regarding confidentiality safeguards, and others are more permissive.^14^ When policies are permissive, they emphasize the importance of using AI to *assist* human review, as opposed to fully unsupervised autonomous AI review. Human supervision ensures that the expert remains responsible for all aspects of peer review, including adherence to recommendations by the Committee on Publication Ethics (COPE) and the International Committee of Medical Journal Editors (ICMJE).^15,16^

Irrespective of formal policies, AI-generated manuscripts and peer-review are influencing academic publishing in seizures. Editors of *Epilepsia* have identified both purely-AI generated manuscripts and peer-reviews. Clear red flags include single-author manuscripts on topics where the author does not have established expertise and peer-reviews that include headers and footers of LLMs with content unrelated to the review.^8^ This study performed a brief practical evaluation of “full stack” AI in academic publishing regarding seizures, comparing previously published human-generated manuscripts and purely AI-generated manuscripts. With minimal human supervision, manuscripts were evaluated by AI reviewers and editors. This highlights both the final editorial decision as well as observation of less glaring indicators of AI generation.

## 2. Methods

### 2.1 Study Structure and Timeline

For this study, three AI-generated manuscripts and three historical human-generated manuscripts published in *Epilepsia* were used.^17–19^ Each manuscript was reviewed by a set of AI tools. Editorial decisions were made based on reviews using several editorial AI tools.

Due to no collection of data from patients or participants, this work was considered exempt from institution review board (IRB) review.

### 2.2 AI-fabricated Manuscript Generation & Human-Generated Manuscript Selection

Three epileptologists each selected a manuscript published in *Epilepsia* within their field of expertise. To maximize comparability of human-generated and AI-fabricated manuscripts, each epileptologist reformatted the human-generated manuscript into a Word document with the included tables and figures, as well as removing identifying markers by removing title page information and other identifying information in the manuscript text (e.g., institution, IRB review, year of conduct and publication).

Each epileptologist also used AI tools of their choice to create an AI-fabricated manuscript. Allowing epileptologists to choose the specific AI tool for manuscript fabrication highlighted the diversity of opinions regarding the current AI tools. The goal of AI-fabrication was to create a manuscript with minimal human supervision other than ensuring appropriate formatting, reference management, and removal of clear red flags for AI generation, including prompt information and other headers and footers from AI tools (e.g., disclaimers that the text is AI generated and follow up questions for next steps in AI generation). Other than removing those clear red flags and providing an individually curated prompt, all text of the manuscript, including abstract, introduction, methods, results, discussion, and conclusion, were generated by AI tools. These studies were entirely fabricated, including quantitative results. If the AI created tables and figures, these were included in the manuscript without modification. If the AI generated references, these also were included in the manuscript without modification.

### 2.3 AI-generated Peer-Review

Each of the epileptologists sent their reformatted human-generated manuscript and AI-fabricated manuscript to a blinded staff member (KNM), who verified absence of clear red flags of AI generation and ensured comparability of the human-generated versus AI-fabricated manuscript. After verification, the staff member sent the epileptologists the manuscripts that they did not use AI to fabricate (i.e., 4 manuscripts for AI-review per epileptologist). The identity of the epileptologist who generated each manuscript was not revealed to the other epileptologists.

Each epileptologist used an AI tool of their choice and an individually crafted prompt to generate “peer”-review comments regarding each manuscript, without the human reading the manuscript. The human expert also did not modify the AI-generated “peer”-review comments other than removing clear red flags for AI-generation. Each epileptologist used prompts based on their personal preferences and style for peer-review.

When returning the AI-generated review, each epileptologist indicated which 2 of the 4 manuscripts they received that they suspected to be AI-fabricated.

### 2.4 AI Editor Decision Making

Each epileptologist sent the AI-generated “peer”-review comments to the blinded staff member, who verified absence of clear red flags for AI generation. After removal of indicators regarding which epileptologist used AI to generate the review, the staff member sent the 2 AI-generated reviews to the epileptologist who submitted each manuscript. Each epileptologist used an AI of their choice to make an editorial decision (accept, minor revisions, major revisions, or reject) based on the manuscript and the received reviews. The epileptologist did not provide any human supervision.

### 2.5 Descriptive Observations Regarding “Full-Stack” AI

After (1) manuscript generation, (2) “peer”-review, and (3) editorial decision making, the identity of which manuscripts were AI-fabricated or human-generated were revealed to all epileptologists, as well as the content of the manuscripts, AI-generated reviews, and editorial decisions.

While this sample size is small (6 manuscripts, 3 epileptologists), this study describes the number and proportion of epileptologists who correctly identified the AI-fabricated manuscripts, as well as similar numbers to describe the final AI’s editorial decisions.

In addition to those quantitative observations, each epileptologist reviewed the AI-fabricated manuscripts, peer-reviews, and editorial decisions to make qualitative observations. Regarding the AI-fabricated manuscripts, the epileptologists comment on whether AI-fabrication could be easily detected, subtle or clear indicators of AI-fabrication, and overall quality. Regarding the AI-generated “peer”-review comments, the epileptologists comment on whether the review adhered to COPE and ICJME recommendations and whether the use of AI was clear. Lastly, the epileptologists comment on the AI’s final editorial decisions regarding appropriateness and quality.

## 3. Results

### 3.1 AI-fabricated and human-generated manuscript information

The 3 epileptologists in this study generated 6 manuscripts, 3 of which were entirely AI-fabricated and 3 of which were historical presumably human-generated manuscripts published in *Epilepsia* (Table 1). The AI-fabricated manuscripts were created by OpenEvidence (April 8^th^, 2026), OpenAI Codex (GPT 5.4, April 7^th^ 2026), and Anthropic Claude (Opus 4.6, April 9^th^, 2026). Prompt information and the full content of the AI-fabricated manuscripts is available in supplemental information.

**Table 1.** Human-generated and AI-fabricated Manuscript Information. The star, *, indicated that generative artificial intelligence (AI) identified potential irregularities suggesting fabrication. The last row refers to if the other 2 experts correctly identified which manuscript was AI-fabricated and which was human-generated.

| Expert | A |  | B |  | C |  |
| --- | --- | --- | --- | --- | --- | --- |
| AI or Human | AI | Human | AI | Human | AI | Human |
| Manuscript Title | Clinician Attitudes Toward Functional Seizures and Epilepsy: A Comparative Study of Stigma | Re-PROGRAM : The evaluation of a brief intervention program for patients with functional seizures in an outpatient hospital setting | The STOP IT Framework: A Prospective Controlled Study of a Nurse-Led Telephone Counseling Intervention Targeting Sleep Deprivation and Alcohol Use as Seizure Triggers in Adults with Epilepsy | Epilepsies after Pocket Monster Seizures | Randomized Controlled Pilot Trial of Daily Chicken Soup in Drug-Resistant Focal Epilepsy | Early seizure freedom with [18F]fluorodopa positron emission tomography response after isocitrate dehydrogenase inhibition with vorasidenib: First case report |
| Editorial Decision | Major Revision* | Major Revision | Major Revision | Major Revision | Reject | Major Revision |
| Expert Identified? | 1 of 2 |  | 1 of 2 |  | 2 of 2 |  |

The overall proportion of epileptologists correctly identifying the AI-fabricated versus human-generated manuscript was 67% (4/6). One epileptologist misidentified the manuscript regarding clinician stigma in functional/dissociative seizures (FDS) as human-generated in comparison to the publication of FDS treatment.^18^ Another epileptologist misidentified the human-generated manuscript regarding the impact of Pocket Monsters (a.k.a. Pokemon) as AI-fabricated in comparison to misidentifying the AI-fabricated STOP IT Framework as human-generated.^17^

### 3.2 AI-generated “peer”-review

Epileptologists used Microsoft Co-pilot (Corporate Microsoft 365 run on May 12^th^, 2026), OpenAI GPT 5.5 Pro (May 8^th^, 2026), and Anthropic Claude Sonnet 4.6 (May 8^th^, 2026) to generate “peer”-review. The prompts used are described in supplemental information.

Generation of the reviews took 1-10 minutes each to create well-organized responses that accurately identified potential limitations and considered the impact of the study in the context of prior literature. These reviews were comprehensive and equally addressed each section of the work, as compared to human peer reviewers who may defer comments on the details of manuscripts (e.g., typographic errors, consistency of numerical results throughout, figure and table formatting) in favor of focusing on the overall approach.

Observations regarding the AI-generated reviews focused on substantial differences between the format and style as compared to human-generated peer-reviews. The most notable difference between AI-generated reviews and typical human-generated peer-reviews was that each of the AI-generated reviews provided unequivocal recommendations of the editorial decision in the author-facing comments (e.g., major revisions or rejection). In human-generated reviews, ICJME recommends only providing those recommendations in editor-facing (author-blinded) comments to enable the editor to make a consensus judgement across multiple reviews, without the perception of internal disagreement by each reviewer listing their individual recommendation.

Another prominent difference between AI-generated reviews was the volume and format of content. Most human-generated reviews include between 1 to 3 pages of content, whereas the AI-generated content commonly was 6 pages or longer. A portion of that length was associated with more use of complex formatting including colors, lines to separate sections, bullet points, and summaries of the impression both as headers and footers. The AI-generated reviews commonly emphasized the limitations of observation or cross-sectional studies, criticisms of interpreting findings as causal instead of associations, and methodological concerns about the validity of the outcomes.

### 3.3 AI Editorial Decisions

The AI’s editorial decision regarding each manuscript is summarized in Figure 1 and Table 1. One of the 3 AI-fabricated manuscripts was recommended to be rejected, whereas the other 5 manuscripts were recommended for major revisions. Epileptologists used Microsoft Copilot (May 14^th^, 2026), OpenAI GPT 5.5 Pro (May 13^th^, 2026), and Claude Sonnet 4.6 (May 13^th^, 2026) to generate these editorial decisions. Prompt and AI output are available in supplemental information.

**Figure 1.**
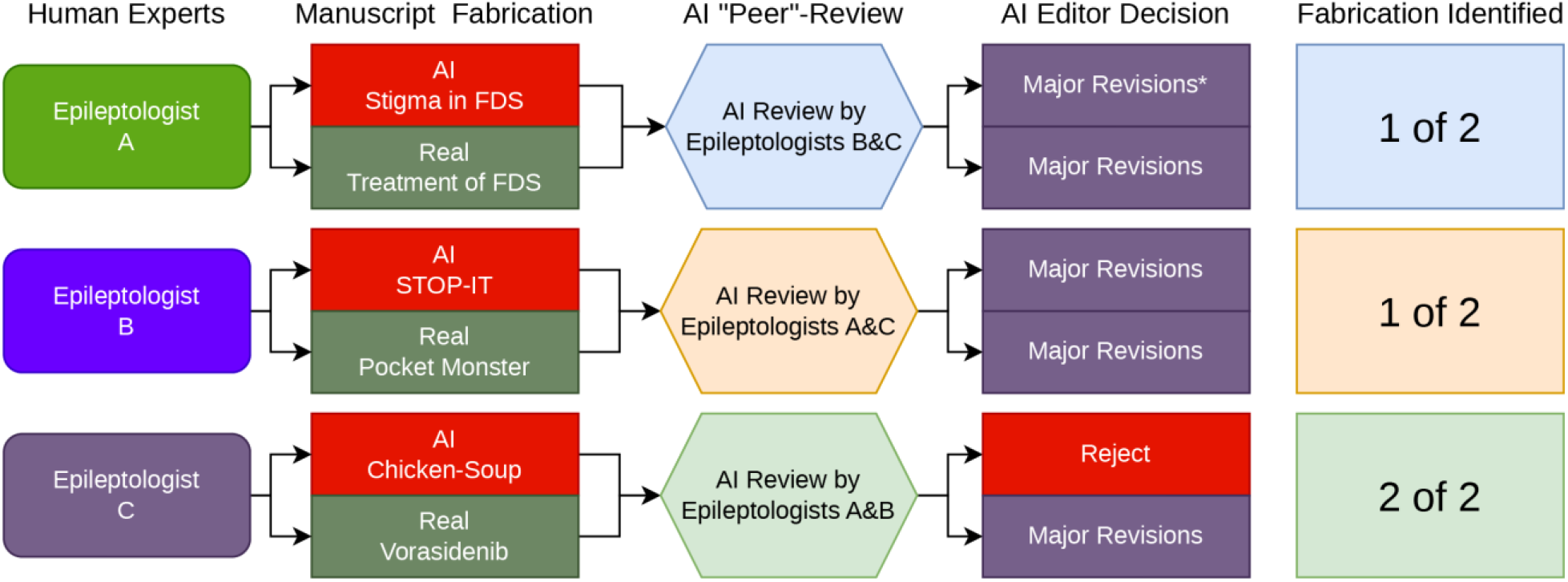
Full Stack generative artificial intelligence (AI) workflow of evaluation and identification. The star, *, indicated that generative artificial intelligence (AI) identified potential irregularities suggesting fabrication.

To describe the epileptologists’ qualitative observations, the AI editors generally did not identify that the AI-fabricated manuscripts were AI-fabricated. An AI editor did identify a yellow flag in one AI-fabricated stigma manuscript that all listed p-values were identical at 0.001. That manuscript was identified as AI-fabricated by 1 of 2 epileptologists.

When qualitatively evaluating the rationale for the AI editor’s decisions, the AI’s phrasing was particularly black and white regarding the benefits and limitations of each manuscript, without appropriate consideration of situations where there was no perfect choice (e.g., emphasis on rule of thumb that there must be 10 participants per characteristic measured in a study). The tone of the rationale also was particularly strong and would be viewed as overly harsh by authors based on phrases of “critical flaw,” “results cannot be trusted,” and “fatal internal inconsistency” despite a decision of major revisions.

The AI editor overemphasized limitations in the manuscripts that were addressed when defining the scope of the work and other acknowledged limitations in the interpretation of the data. For the AI-fabricated manuscript regarding stigma towards FDS, the AI editor criticized a qualitative analysis as “lack[ing] methodological transparency” despite the manuscript clearly indicating that portion of the work was descriptive. Such a situation might be considered “AI hallucination.” Separately in the human-generated manuscript describing the pilot outcomes of treatment for FDS, the authors stated that there were no established outcomes for FDS, which prompted them to use an outcome that they had demonstrated in a prior manuscript. Even though the study was describing a pilot evaluation to prepare for a subsequent clinical trial, the AI agent commented that “outcome measurement is weak and potentially unreliable.” Such a comment suggests a lack of “understanding” the broader context of the study’s place in the medical literature. Additionally, even though prior literature showed a 30% completion rate for FDS treatment, the AI editor commented that a 55% completion rate for this treatment had “severe risk of selection and response bias.” While the concern for selection and response bias is valid, a rejection based on that concern would not be reasonable because of the similarity to prior literature and typical clinical and research practice.

## 4. Discussion

This brief, illustrative study demonstrated both benefits and cautions about the use of AI tools in each step of academic publishing in seizures from manuscript generation, peer-review, and editorial decision making. The quality of the AI-fabricated manuscripts and review commentary was surprisingly high, which demonstrated the importance of this evaluation to both show the utility of these tools and highlight challenges in identifying unethical behavior.^8,12^ While some red flags for AI-fabrication of academic work are obvious,^8^ these results are presented to assist authors, reviewers, and editors in identifying more nuanced indicators of AI-generation in peer review literature in seizures.

The ease with which three fabricated manuscripts were produced was alarming. It is worth noting that many brilliant scientists are not equally adept at communication and writing.^12,20,21^ The advantage of having the ability of AI to assist in producing well-written scientific manuscripts for real scientific work is quite apparent, and there is great potential for AI assistance to improve communication of important science to wider audiences.^22^ When using AI assistance for peer review, scientists with less writing experience must identify when an AI-generated first draft of a review did not adhere to ICJME and COPE recommendations. In response, the scientist must modify the review using human-generated content.^23^ Similarly, the overly strong tone of the editorial decision would be recognized immediately by a human editor, who could either manually change the tone of the text or use prompt engineering to shift the content from purely AI-generated to be AI-assisted.^24^

These results must be viewed in the context of continued exponentially increasing number of peer-reviewed publications, without a commensurate exponential increase in well-trained scientists.^6^ This increasing discrepancy may be associated with an increase in the time from submission to editorial decision, which can be 6 to 12 months in some journals.^25^ Consequentially, the goal of the *Epilepsia* editorial board to maintain a time from submission to decision of 30 days or less is challenging. These results demonstrate that AI-assistance, not AI-generation, has the potential to accelerate scientific communication and bring new discoveries to patients faster.^21^ Human review remains required in all steps of the editorial process, while also recognizing the that cognitive load of reviewing AI-generated material is lower than producing “from-scratch” reviews because during review, our brains engage in passive verification rather than active synthesis.^26,27^

While this study’s illustrative sample was small, “full stack” AI for academic publishing only rejected 1 of the 3 AI-fabricated manuscripts, and the other 2 manuscripts were incorrectly identified as human-generated by 1 of 2 epileptologists. The AI-fabricated manuscript that was rejected was more clearly fabricated because it evaluated chicken soup as an intervention for epilepsy, which was quickly identified as fabricated when read by humans. However, the other 2 AI-fabricated manuscripts were incompletely identified as fabricated by humans and recommended for major revisions by full-stack AI. This identification rate was comparable to a prospective study where the sensitivity was 31% and specificity was 56% to identify a manuscript generated by a much less capable version of generative AI (ChatGPT 3.5 from 2022).^28^ For one of the AI-fabricated manuscripts, the only identified indicator of fabrication was the use of identical p-values of 0.001 throughout the results. Compared to some more blatant red flags of AI-fabrication that have been described previously,^8^ these indicators were much more subtle.

In contrast, AI-generated “peer”-reviews were both qualitatively and quantitatively different from typical human peer-reviews. Even though this study removed obvious evidence of AI-generation in manuscripts, the AI tools used here had clear indicators that have been described outside of the seizure literature. Human peer-reviewers commonly provide relatively succinct comments that focus on the core concerns for manuscripts in a constructive manner, whereas these AI comments were markedly verbose and utilized more complex formatting and structure.^23,29^ Additionally, human peer-reviewers appeared more understanding of the value of well-conducted observational and cross-sectional studies to identify associations in clinical research,^30^ whereas the AI may have overemphasized the need for prospective studies to make causal conclusions using highly validated outcomes.^23,31^ In contrast to AI, human reviewers may be aware of the inherent limitations of clinical research in seizures and thereby not view those criticisms as fundamental flaws.^31^

The differences between AI versus human peer-reviewers also were apparent in the qualitative reasoning of the AI editors. The observation that all 3 human-generated manuscripts and 2 of the AI-fabricated manuscripts had an editorial decision of major revisions was consistent with the non-AI-fabricated literature; after passing the initial editorial screen, almost all quality manuscripts get major revisions after first submission. In many journals, the eventual acceptance rates for manuscripts that were revised after major revisions were 80% or higher.^32,33^ When considering that 2 of these manuscripts were AI-fabricated, that rate of major revisions may indicate that these AI tools were able to fabricate manuscripts that were high enough quality for publication after revisions. After recognizing that AI-detectors had imperfect performance (5-19% false positive rates, 58-74% sensitivity),^10,34,35^ AI-detectors suggest that perhaps 17% of text in academic journals may be largely AI-generated.^36^ Those evaluations focused on identification of AI-generated text, which has markedly different ethical implications than fully AI-fabricated studies. These results substantially raise the concern that the quality of these AI tools was high enough that the typical methods of quality oversight with peer review were not sufficient.

This illustrative study has important limitations. Due to the focus on illustration of the current concerns with the potential benefits and cautions about AI, the sample size of 6 manuscripts evaluated by 3 epileptologists was insufficient for fully powered statistical analysis.^12,37^ As these AI technologies continue to increase in sophistication, the factors indicating AI fabrication here were relevant to the current tools.^38^ Similar to how the previous obvious red flags were not apparent in AI-fabricated work, human peer-reviewers and editors must remain up to date regarding the methods to identify AI-generated or AI-fabricated content, even if they do not use AI tools themselves.^39–41^ Additionally, there are numerous AI tools for text generation and evaluation available which are being updated and released almost every week.^1^ This work represents the current state of AI tools because these rapid developments hinder the ability to comprehensively review all the available tools in real time.^27^

## 5. Conclusions

This study of using full stack AI to fabricate manuscripts, serve as “peer”-reviewers, and make editorial decisions is a cautionary tale demonstrating the maturity of these technologies to create content of sufficient quality to pass as legitimate clinical studies about seizures. Conversely, this demonstrates the opportunity for these tools to reduce barriers to writing manuscripts and increase the speed of human researchers, reviewers and editors, which can have tremendous beneficial effects increasing the pace of clinical innovation and dissemination. Simultaneously, this study identifies the potential for maligned parties to use these same technologies to fabricate manuscripts without clear indicators of AI-fabrication. Additionally, we highlighted non-obvious characteristics of AI-generated peer-reviews that authors and editors can use to maintain the quality of academic publishing in seizures.

## Data Availability

All data produced in the present work are contained in the supplemental materials.

## 6. Funding and Acknowledgements

WTK’s research time was supported, in part, by the National Institute of Neurological Disorders and Stroke (NINDS, K23NS135134). JP has no relevant disclosures. DMG was supported by NIH K23NS124656, R21NS142800, and American Board of Psychiatry and Neurology (ABPN).

## 7. Ethical Publication, Disclosures, & Conflicts of Interest

The authors confirm that they have read the Journal’s position on issues involved in ethical publication and affirm that this report is consistent with those guidelines. Dr. Kerr is an Associate Editor of *Epilepsia*; writes review articles for Medlink Neurology; has received research support from NINDS, SK Life Science, Biohaven Pharmaceuticals, Neurelis, Neuropace, Neumirna, and QurAlis; is a paid consultant for SK Life Sciences, the Epilepsy Study Consortium, EpiTel, UCB Pharmaceuticals, Jazz Pharmaceuticals, and Biohaven Pharmaceuticals; has collaborative or data use agreements with Eisai, Janssen, Radius Health, GSK, and Praxis. Dr. Pellinen has received research support from NIH/NINDS, the American Epilepsy Society, and SK Life Science. Ms. McFarlane is a paid consultant for Neurelis, Neumirna, and Neuropace. Dr. Goldenholz has been provided speaker fees from Harvard Medical School, AAN, AES, ACNS, NNS, AI in Epilepsy and Neurology, Florida Epilepsy Alliance, and UT-Austin. He also previously has been a paid consultant for Neuro Event Labs, IDR, LivaNova, Health Advances, Duke University, Bloom Insights and Wiley. He has received grants from NIH, ABPN, BIDMC and the Lions Club.

## 8. Author Contributions

Dr. Goldenholz proposed the idea for this manuscript to Drs. Pellinen and Kerr. Drs. Goldenholz, Pellinen, and Kerr collaborated on all aspects of the work. Dr. Kerr wrote the first draft. Ms. McFarlane assisted with anonymization and data handling. All authors reviewed manuscript drafts and approved the final version of the work.

## Notes

### Competing Interest Statement

The authors confirm that they have read the Archive's position on issues involved in ethical publication and affirm that this report is consistent with those guidelines. Dr. Kerr is an Associate Editor of *Epilepsia*; writes review articles for Medlink Neurology; has received research support from NINDS, SK Life Science, Biohaven Pharmaceuticals, Neurelis, Neuropace, Neumirna, and QurAlis; is a paid consultant for SK Life Sciences, the Epilepsy Study Consortium, EpiTel, UCB Pharmaceuticals, Jazz Pharmaceuticals, and Biohaven Pharmaceuticals; has collaborative or data use agreements with Eisai, Janssen, Radius Health, GSK, and Praxis. Dr. Pellinen has received research support from NIH/NINDS, the American Epilepsy Society, and SK Life Science. Ms. McFarlane is a paid consultant for Neurelis, Neumirna, and Neuropace. Dr. Goldenholz has been provided speaker fees from Harvard Medical School, AAN, AES, ACNS, NNS, AI in Epilepsy and Neurology, Florida Epilepsy Alliance, and UT-Austin. He also previously has been a paid consultant for Neuro Event Labs, IDR, LivaNova, Health Advances, Duke University, Bloom Insights and Wiley. He has received grants from NIH, ABPN, BIDMC and the Lions Club.

