## Supplemental Information for "Full stack academic publishing in seizures with artificial intelligence: a cautionary tale"

The outline of this document is as follows:

Supplemental Methods

1. Artificial Intelligence Prompts
   1.1 Generation of Manuscripts
   1.2 Generation of Reviews
   1.3 Generation of Editorial Decisions

Supplemental Results

2. Artificial Intelligence-Generated Materials

2.1 Manuscripts

2.2 Reviews

2.3 Editorial Decisions

**Supplemental Methods**

1. **Artificial Intelligence Prompts**
   1.1 **Generation of Manuscripts**

Three epileptologists each created an artificial intelligence (AI)-generated manuscript using the AI tool of their choice to accompany the human-generated papers published in *Epilepsia* selected by the epileptologists. The epileptologists independently drafted prompts, which are presented below.

*Manuscript Generation A*

Epileptologist #A used OpenEvidence on April 8^th^, 2026 to generate an AI-fabricated paper using the following prompts:

Prompt 1:

You're an expert in functional/dissociative seizures and epilepsy. Can you help me write a first draft of an introduction for a manuscript describing clinician stigma towards functional seizures as compared to epilepsy?

Prompt 2:

Can you help me write the first draft of the methods that surveyed clinicians who work in an epilepsy monitoring unit (EMU) with stigma that was evaluated using the attribution questionnaire with 9 domains of stigma?Top of Form

Prompt 3:

Can you give proposed clinical vignettes for this section: The survey presented two clinical vignettes describing patients with paroxysmal episodes: one depicting a patient with functional seizures and one depicting a patient with epilepsy. [DESCRIBE VIGNETTE CONTENT BRIEFLY: e.g., "Vignettes were matched for demographic characteristics, symptom severity, and clinical presentation, differing only in the diagnostic label and underlying pathophysiology."] The order of vignette presentation was randomized to minimize order effects.Top of Form

Prompt 4:

What are the typical 5-point Likert scale responses for an Attribution Questionnaire?Top of Form

Prompt 5:

Can you give me a first draft of what the results of this study might look like?Top of Form

Prompt 6:

Yes please. Can you help create the tables references in the results section?

Prompt 7:

Can you give me examples of what the X's might be expected to be in this section: The mean age was [X] years (SD = [X]), and [X]% identified as female. The sample included [X] physicians ([X]%), [X] advanced practice providers ([X]%), [X] nurses ([X]%), and [X] other healthcare professionals ([X]%). The median duration of clinical experience was [X] years (IQR: [X]-[X]). Among physician respondents, [X]% had completed fellowship training in epilepsy.

Prompt 8:

Can you fill in the X's with hypothetical values: Table 2 presents mean Attribution Questionnaire domain scores for functional seizure and epilepsy vignettes. Clinicians attributed significantly greater responsibility to patients with functional seizures compared to those with epilepsy (mean difference = [X], 95% CI: [X] to [X], p 0.001, Cohen's d = [X]). Respondents also reported significantly less pity toward patients with functional seizures (mean difference = [X], 95% CI: [X] to [X], p 0.001, d = [X]) and greater anger (mean difference = [X], 95% CI: [X] to [X], p = [X], d = [X]).

Clinicians were significantly more likely to endorse coercive treatment approaches for functional seizures compared to epilepsy (mean difference = [X], 95% CI: [X] to [X], p 0.001, d = [X]). Avoidance scores were also higher for functional seizures (mean difference = [X], 95% CI: [X] to [X], p = [X], d = [X]), indicating greater desire to avoid professional contact with these patients.

Conversely, clinicians perceived patients with functional seizures as less dangerous compared to those with epilepsy (mean difference = [X], 95% CI: [X] to [X], p = [X], d = [X]). No significant differences were observed in help-seeking endorsement (p = [X]) or segregation attitudes (p = [X]).

The composite stigma score was significantly higher for functional seizures compared to epilepsy (mean = [X] vs. [X], mean difference = [X], 95% CI: [X] to [X], p 0.001, d = [X]).

AQ-27 Domain	Functional Seizures Mean (SD)	Epilepsy Mean (SD)	Mean Difference (95% CI)	p-value	Cohen's d	References

Responsibility	[X] ([X])	[X] ([X])	[X] ([X] to [X])	[X]	[X]	[1-2]

Pity	[X] ([X])	[X] ([X])	[X] ([X] to [X])	[X]	[X]	[1-2]

Anger	[X] ([X])	[X] ([X])	[X] ([X] to [X])	[X]	[X]	[1-2]

Dangerousness	[X] ([X])	[X] ([X])	[X] ([X] to [X])	[X]	[X]	[1-2]

Fear	[X] ([X])	[X] ([X])	[X] ([X] to [X])	[X]	[X]	[1-2]

Help	[X] ([X])	[X] ([X])	[X] ([X] to [X])	[X]	[X]	[1-2]

Coercion	[X] ([X])	[X] ([X])	[X] ([X] to [X])	[X]	[X]	[1-2]

Segregation	[X] ([X])	[X] ([X])	[X] ([X] to [X])	[X]	[X]	[1-2]

Avoidance	[X] ([X])	[X] ([X])	[X] ([X] to [X])	[X]	[X]	[1-2]

Total Composite Score	[X] ([X])	[X] ([X])	[X] ([X] to [X])	[X]	[X]	[1-2]

Prompt 9:

Can you fill in the X's here with hypothetical values: In multivariable linear regression models adjusting for age, sex, professional role, and years of experience, several factors were associated with higher stigma scores toward functional seizures (Table 3). Nurses demonstrated significantly higher stigma scores compared to physicians (β = [X], 95% CI: [X] to [X], p = [X]). Clinicians with fewer years of experience (5 years) reported higher stigma scores compared to those with ≥10 years of experience (β = [X], 95% CI: [X] to [X], p = [X]).

Epilepsy fellowship training among physicians was associated with lower stigma scores (β = [X], 95% CI: [X] to [X], p = [X]). No significant associations were found between stigma scores and clinician age (p = [X]) or sex (p = [X]).

Variable	β Coefficient	95% CI	p-value	References

Professional Role (ref: Physicians)

Advanced practice providers	[X]	[X] to [X]	[X]	[1]

Nurses	[X]	[X] to [X]	[X]	[1]

EEG technologists	[X]	[X] to [X]	[X]	[1]

Years of Experience (ref: ≥10 years)

5 years	[X]	[X] to [X]	[X]	[1]

5–9 years	[X]	[X] to [X]	[X]	[1]

Epilepsy Fellowship Training (physicians only)	[X]	[X] to [X]	[X]	[1]

Prior Education on Functional Seizures	[X]	[X] to [X]	[X]	—

Age (per 10-year increase)	[X]	[X] to [X]	[X]	[1]

Female Sex	[X]	[X] to [X]	[X]	[1]

Prompt 10:

This is applicable. Please fill in with hypothetical responses: [IF APPLICABLE: "Of [X] respondents, [X] ([X]%) provided free-text comments. Common themes included frustration with perceived lack of treatment options for functional seizures ([X]%), uncertainty about appropriate management ([X]%), and concerns about patients' engagement with psychiatric referrals ([X]%). Several respondents ([X]%) expressed empathy for patients with functional seizures while acknowledging their own discomfort in managing these cases."]

Prompt 11:

Can you help draft a discussion section based on these results: Participant Characteristics

Of 100 eligible clinicians working in the EMU, 75 completed the survey, yielding a response rate of 75%. Table 1 presents the demographic and professional characteristics of respondents. The mean age was 42.5 years (SD = 10.8), and 68% identified as female. The sample included 18 physicians (24%), 8 advanced practice providers (11%), 38 nurses (51%), and 11 other healthcare professionals (15%, including EEG technologists and social workers). The median duration of clinical experience was 12 years (IQR: 5-18). Among physician respondents, 61% had completed fellowship training in epilepsy.

Characteristic	Total Sample

(N = 75)

Demographics

Age, mean (SD), years	42.5 (10.8)

Female sex, n (%)	51 (68%)

Professional Role, n (%)

Physicians	18 (24%)

Epilepsy fellowship-trained	11 (15%)

Advanced practice providers	8 (11%)

Nurses	38 (51%)

EEG technologists	9 (12%)

Other	2 (3%)

Clinical Experience

Years in practice, median (IQR)	12 (5–18)

Years working in EMU, median (IQR)	8 (3–18)

Prior Training/Exposure

Prior education on functional seizures, n (%)	75 (100%)

Personal/family history of seizures, n (%)	10 (13%)

Comparison of Stigma Scores Between Functional Seizures and Epilepsy

Table 2 presents mean Attribution Questionnaire domain scores for functional seizure and epilepsy vignettes. Clinicians attributed significantly greater responsibility to patients with functional seizures compared to those with epilepsy (mean difference = 2.4, 95% CI: 1.9 to 2.9, p < 0.001, Cohen's d = 1.12). Respondents also reported significantly less pity toward patients with functional seizures (mean difference = -1.8, 95% CI: -2.3 to -1.3, p < 0.001, d = 0.89) and greater anger (mean difference = 1.5, 95% CI: 1.0 to 2.0, p = < 0.001, d = 0.74).

Clinicians were significantly more likely to endorse coercive treatment approaches for functional seizures compared to epilepsy (mean difference = 1.3, 95% CI: 0.8 to 1.8, p < 0.001, d = 0.65). Avoidance scores were also higher for functional seizures (mean difference = 1.1, 95% CI: 0.6 to 1.6, p = < 0.001, d = 0.58), indicating greater desire to avoid professional contact with these patients.

Conversely, clinicians perceived patients with functional seizures as less dangerous compared to those with epilepsy (mean difference = -1.6, 95% CI: -2.1 to -1.1, p = < 0.001, d = 0.78). No significant differences were observed in help-seeking endorsement (p = 0.34) or segregation attitudes (p = 0.42).

The composite stigma score was significantly higher for functional seizures compared to epilepsy (mean = 42.6 vs. 36.8, mean difference = 5.8, 95% CI: 4.2 to 7.4, p < 0.00

AQ-27 Domain	Functional Seizures Mean (SD)	Epilepsy Mean (SD)	Mean Difference (95% CI)	p-value	Cohen's d

Responsibility	5.8 (2.1)	3.4 (2.0)	2.4 (1.9 to 2.9)	0.001	1.12

Pity	4.2 (2.0)	6.0 (1.9)	-1.8 (-2.3 to -1.3)	0.001	0.89

Anger	4.1 (2.2)	2.6 (1.8)	1.5 (1.0 to 2.0)	0.001	0.74

Dangerousness	2.8 (1.6)	4.4 (2.1)	-1.6 (-2.1 to -1.1)	0.001	0.78

Fear	2.4 (1.5)	3.6 (1.9)	-1.2 (-1.7 to -0.7)	0.001	0.62

Help	5.6 (1.8)	5.8 (1.7)	-0.2 (-0.6 to 0.2)	0.34	0.11

Coercion	4.5 (2.0)	3.2 (1.9)	1.3 (0.8 to 1.8)	0.001	0.65

Segregation	2.2 (1.4)	2.0 (1.3)	0.2 (-0.2 to 0.6)	0.42	0.14

Avoidance	4.2 (1.9)	3.1 (1.8)	1.1 (0.6 to 1.6)	0.001	0.58

Total Composite Score	42.6 (9.4)	36.8 (8.2)	5.8 (4.2 to 7.4)	0.001	0.72

Scores range from 1 to 9 for each domain (total composite range 9-81). Higher scores indicate greater endorsement of the construct. Negative mean differences indicate higher scores for epilepsy.

Factors Associated With Stigma Toward Functional Seizures

In multivariable linear regression models adjusting for age, sex, professional role, and years of experience, several factors were associated with higher stigma scores toward functional seizures (Table 3). Nurses demonstrated significantly higher stigma scores compared to physicians (β = 4.8, 95% CI: 1.9 to 7.7, p = 0.002). Clinicians with fewer years of experience (<5 years) reported higher stigma scores compared to those with ≥10 years of experience (β = 5.2, 95% CI: 2.1 to 8.3, p = 0.001).

Epilepsy fellowship training among physicians was associated with lower stigma scores (β = -6.4, 95% CI: -10.2 to -2.6, p = 0.001). No significant associations were found between stigma scores and clinic

Variable	β Coefficient	95% CI	p-value

Professional Role (ref: Physicians)

Advanced practice providers	2.1	-1.4 to 5.6	0.24

Nurses	4.8	1.9 to 7.7	0.002

EEG technologists	3.6	0.2 to 7.0	0.04

Years of Experience (ref: ≥10 years)

5 years	5.2	2.1 to 8.3	0.001

5–9 years	2.4	-0.3 to 5.1	0.08

Epilepsy Fellowship Training (physicians only)	-6.4	-10.2 to -2.6	0.001

Prior Education on Functional Seizures	-3.8	-6.5 to -1.1	0.006

Age (per 10-year increase)	-0.4	-2.1 to 1.3	0.68

Female Sex	1.2	-1.6 to 4.0	0.41

Model adjusted for all variables shown. Dependent variable is total AQ-27 composite score for functional seizure vignette (range 9-81). Negative β coefficients indicate lower stigma scores.

Qualitative Comments

Of 75 respondents, 34 (45%) provided free-text comments. Common themes included frustration with perceived lack of treatment options for functional seizures (47%), uncertainty about appropriate management (38%), and concerns about patients' engagement with psychiatric referrals (32%). Several respondents (21%) expressed empathy for patients with functional seizures while acknowledging their own discomfort in managing these cases. These were illustrative quotations of these themes:

•	Frustration with treatment options: "Once we diagnose functional seizures, we feel like there's nothing more we can offer them in the EMU."

•	Uncertainty about management: "I'm never sure how to respond when a patient is having a functional seizure—should I intervene or just observe?"

•	Psychiatric referral concerns: "Most of these patients refuse to see psychiatry, so the diagnosis feels like a dead end."

•	Empathy with discomfort: "I feel bad for these patients—they're clearly suffering—but I honestly don't know how to help them."

Prompt 12:

Can you please draft an abstract with a word limit of 200 please? The abstract does not need structure

Prompt 13:

Can you help me draft a 100 word summary please?

Prompt 14:

This discussion section does not have references. Can you fill in the appropriate references in the right section: This study provides the first systematic comparison of stigmatizing attitudes toward functional seizures versus epilepsy among clinicians working in an epilepsy monitoring unit. The findings reveal that EMU clinicians harbor significantly greater stigma toward patients with functional seizures across multiple domains, with particularly large effects observed for attributions of responsibility, reduced pity, and increased anger. These results have important implications for clinical care, patient outcomes, and staff education in epilepsy centers.

Principal Findings and Interpretation

The most striking finding was the large effect size (Cohen's d = 1.12) for responsibility attributions, indicating that clinicians perceive patients with functional seizures as substantially more responsible for their condition compared to those with epilepsy. This pattern aligns with attribution theory, which posits that conditions perceived as controllable elicit less sympathy and more negative emotional responses from observers. The corresponding reductions in pity (d = 0.89) and increases in anger (d = 0.74) further support this theoretical framework and mirror findings from stigma research in other functional neurological disorders and psychiatric conditions.

The endorsement of coercive treatment approaches for functional seizures (d = 0.65) is particularly concerning, as it suggests clinicians may favor paternalistic or directive management strategies that could undermine the therapeutic alliance essential for effective treatment. Similarly, elevated avoidance scores (d = 0.58) indicate a desire to limit professional contact with these patients, which may manifest as shorter consultations, delayed follow-up, or premature discharge—all of which could adversely affect patient care.

Notably, clinicians did not differ in their willingness to help patients with functional seizures compared to epilepsy, nor did they endorse segregation. This suggests that stigma operates through specific attributional pathways rather than reflecting global rejection of patients with functional seizures. Clinicians appear willing to provide care but may do so with emotional responses and behavioral tendencies that patients can perceive, potentially contributing to the felt stigma widely reported in this population.

The finding that patients with epilepsy were perceived as more dangerous likely reflects awareness of physical injury risk during epileptic seizures, including falls, burns, and sudden unexpected death in epilepsy (SUDEP). This perception, while clinically grounded, may paradoxically contribute to greater sympathy for epilepsy while reinforcing the view that functional seizures are less medically serious.

Factors Associated With Stigma

The identification of modifiable factors associated with stigma provides actionable targets for intervention. Epilepsy fellowship training demonstrated the strongest protective effect (β = -6.4), suggesting that specialized education in seizure disorders—which typically includes training on functional seizures—substantially reduces stigmatizing attitudes. This finding underscores the importance of incorporating comprehensive functional seizure education into neurology training programs.

The association between nursing role and higher stigma scores (β = 4.8) warrants careful interpretation. Nurses comprise the majority of EMU staff and have the most sustained direct contact with patients during monitoring. This prolonged exposure may amplify frustration when patients exhibit events that do not correlate with EEG changes, particularly if nurses feel inadequately equipped to respond. Rather than reflecting inherent attitudes, this finding may indicate a need for targeted nursing education and support.

Similarly, less experienced clinicians (5 years) demonstrated higher stigma scores (β = 5.2), suggesting that clinical experience—and the perspective it provides—may naturally attenuate stigmatizing attitudes over time. However, waiting for experience to reduce stigma is not an acceptable strategy; proactive educational interventions for early-career clinicians are warranted.

Qualitative Insights

The qualitative findings provide essential context for interpreting the quantitative results. Nearly half of respondents provided comments, and the dominant themes—frustration with treatment options, management uncertainty, and concerns about psychiatric referral engagement—reveal that stigma may partly stem from clinicians' sense of therapeutic helplessness rather than moral judgment of patients.

The illustrative quotation "Once we diagnose functional seizures, we feel like there's nothing more we can offer them in the EMU" encapsulates a critical misconception: that diagnosis represents an endpoint rather than a therapeutic intervention. In fact, clear and empathic communication of the diagnosis has been shown to be therapeutic in itself and is associated with improved outcomes. Reframing the diagnostic disclosure as a treatment opportunity—rather than a dismissal—may help address clinician frustration while improving patient care.

The 21% of respondents who expressed empathy alongside discomfort represent an important subgroup. These clinicians recognize patient suffering but feel ill-equipped to help, suggesting that educational interventions emphasizing practical management strategies may be particularly effective for this group.

Clinical Implications

These findings suggest several strategies for reducing clinician stigma in EMU settings. First, structured education on functional seizures should be provided to all EMU staff, with particular attention to nurses and early-career clinicians. This education should address the neurobiological basis of functional seizures, emphasize that symptoms are genuine and involuntary, and provide practical guidance for bedside management during events.

Second, training in diagnostic disclosure techniques may help clinicians view the diagnosis as a therapeutic intervention rather than a dead end. Approaches that emphasize explaining what functional seizures are (rather than what they are not), validating patient experiences, and providing a clear treatment pathway have been associated with improved patient acceptance and outcomes.

Third, EMU protocols should include clear guidance for responding to functional seizures during monitoring, addressing the uncertainty expressed by clinicians about whether to intervene or observe. Standardized approaches may reduce clinician anxiety and improve consistency of care.

Finally, fostering interdisciplinary collaboration between neurology, psychiatry, and psychology services may help address concerns about psychiatric referral engagement. When mental health providers are integrated into the EMU team rather than serving as external consultants, patients may be more receptive to psychological treatment approaches.

Limitations

Several limitations should be considered when interpreting these findings. First, this study was conducted at a single academic EMU, and results may not generalize to community hospitals or other practice settings. Second, the cross-sectional design precludes causal inference; while we identified factors associated with stigma, we cannot determine whether these factors cause stigmatizing attitudes or are merely correlated with them. Third, social desirability bias may have led respondents to underreport stigmatizing attitudes, suggesting that our findings may underestimate the true magnitude of stigma. Fourth, the vignette-based methodology, while allowing controlled comparison, may not fully capture the complexity of real clinical encounters. Finally, the sample size limited statistical power for subgroup analyses, particularly among physicians with and without fellowship training.

Future Directions

Future research should examine whether educational interventions targeting the factors identified in this study effectively reduce stigma and, more importantly, whether stigma reduction translates to improved patient outcomes. Longitudinal studies tracking clinician attitudes over time and in response to specific training programs would help establish causal relationships. Additionally, studies examining patient perceptions of clinician stigma and its impact on treatment engagement, diagnostic acceptance, and functional outcomes would complement this clinician-focused investigation.

*Manuscript Generation B*

Epileptologist B used OpenAI Codex 5.4 on April 7^th^, 2026 to generate an AI-fabricated paper using the following prompts:

Prompt 1:

Use the chocolates simulator to simulate a set of patients with one count per day providing 8 months of data. Keep generating new patients until 70 patients qualify in that they have 2 months averaging 4 seizures per month. Then record each patient in a CSV file, one row per patient. Columns will be patient ID, group assignment (placebo or test), weekly count (for 24 columns). Half the patients should be assigned placebo, the other half to test. 30% of the patients will drop out somewhere between week 10 and 24 (randomly chosen from a uniform distribution). Any week during dropout will have a count of -1.The file should be called patientData.csv.

Prompt 2:

write a jupyter notebook that takes the patientData.csv file in and reports number of patients per group, seizure frequency during first 3 months (min, max, median) per treatment group, dropout per treatment group. Then assume the first 3 months are eligibility, 2 months are baseline measurement, and 3 months are test period. Compute the percentage change in monthly seizure rate between baseline and test period for each patient. Make 2 waterfall plots of percentage change for each treatment group. Run each script in the jupyter notebook and save with outputs and figures.

Prompt 3:

Also add a calculation for 50% responder rate for each group, and do a Fisher exact test to see if there is a statistical difference between then two group. Then calculate the median percentage change for each group, report that. Then compute the Mann Whiteney U test for percentage change between the two groups and report that too.

Prompt 4:

1. Rebuild the patientData.csv file and make sure that every single patient has a minimum seizure rate of 4 or higher during their first 3 months - use the same generation rules as previously otherwise. 2.revise the patientData_analysis. For any patient that drops out, use a last obersvation carried forward analysis. When you plot the two waterfall plots, put them together on one figure using color to separate them. Plot a 50% responder rate plot for the two groups. Save those figures as Figure 1 and Figure 2 at 300 dpi JPG files.

Prompt 5:

1. regenerate the patient data, this time only make 44 patients, otherwise same rules as before. 2. recompute results in the notebook.

Prompt 6:

1. Update all percentage change calculations such that PC = 100* (basline rate - test rate) / baseline rate. 2. The waterfall plot figure should put the two subplots vertically aligns with an A and a B to the side of each subplot. 3. build a figure 3 at 300 dpi which show percentage change for the two groups. 4. rebuild the patient data with 30 patients total, same rules as before. 5. recalculate everything based on the new PC calculation in the notebook.

Prompt 7:

1. renumber the plots such that the current figure 3 is now 1, figure 1 is 2 and figure 2 is 3. 2. add a horizontal bar and a p value above the two groups showing the statistical comparison between the two in the new figure 1 based on the mann whitney u test. 3. Make sure that the waterfall plot has a y axis of -100 to 100 % improved, and that the legend does not intersect the data. 4. now regenerate the data, and recalculate everything.

Prompt 8:

Based on the data, the figures and the results from the notebook, write a manuscript for submission to Epilepsia. The paper is being produced as a proof of concept that AI can generate papers based on fictitious data - this information will be presented publicly in a companion study. But this paper itself will not reveal that any pof the data is artificially generated. This paper will be in the Brief Communication style. Look up the exact specifications for the paper. The study is titled "Placebo controlled randomized trial of chicken soup as a treatment for drug resistant focal epilepsy". Write a manuscript that meets all the critiera of the journal. Include introductory comments about the importance of treating drug resistant focal epilepsy, and how chicken soup has long been considered a home remedy for many conditions. In the methods section, generate content that indicates the data was produced in accordance with the IRB of the University of Botlogwa, and all participants provided informed consent. Generate all the needed sections, produce plausible (real and verified) references to ideas used, and explain why although the results were not statistically significant there was a positive trend towards chicken soup being beneficial for seizure reduction. After generating the manuscript in .docx format, double check that the paper meets all the requirements, has no grammatical or conceptual errors, has no formatting problems or citation problems, and double check that the citations are real and appropriate.

*Manuscript Generation C*

Epileptologist C used Anthropic Claude Opus 4.6 on April 9^th^, 2026 to generate an AI-fabricated paper using the following prompt:

Use the instructions for authors found here: <https://onlinelibrary>.wiley.com/page/journal/15281167/homepage/forauthors.html to write a “Manuscript Type: Brief Communication” for study testing an intervention package called the “STOP IT” framework for reducing seizure triggers in people with epilepsy who are being followed in an outpatient epilepsy clinic at a tertiary referral academic center. Write the paper in the tone and style of an expert academic epileptologist at a premier academic center in the US. The “STOP IT” clinical intervention consists of identifying whether or not a patient has either alcohol use or sleep deprivation from “staying up too late” at night as a seizure trigger. If they report this in clinic, patients are counseled on ways of stopping this behavior. They are then contacted by phone on a weekly basis by a clinical nursing team over a 3 month period to enquire about continuing bad habits, and if continuing to do them, they are again counseled on cessation. After 3 months, the calls stop, but 1 year seizure frequency outcomes are collected as a final outcome marker, and patients in the “STOP IT” clinical intervention arm are compared to patients who report these seizure triggers but who do not undergo the “STOP IT” clinical intervention. Please use up-to-date citations, using the instructions for authors guidelines, and ensure accuracy of reporting that also adheres to clinical reporting guidelines for the study design.

1.2 **Generation of Reviews**

After receiving the four papers generated or selected by the other epileptologists, they each used an AI tool of their choice, which could differ from the AI tool used to generate the manuscripts, to generate reviews of each paper. The prompts to create these reviews are below.

*Review Generation A*

Epileptologist A used Microsoft 365 Copilot on May 12^th^, 2026 to generate AI reviews using the following prompt:

I'd like you to help with the first draft of a peer review for a manuscript. You're an expert in biostatistics and epilepsy. Here's examples of my style for these peer reviews.

Supplied files: EPI-***-2026_Proof_hiEPI-***-2026_Proof_hiEPI-***-2026

Can you please give me a first draft of a peer review for this manuscript?

*Review Generation B*

Epileptologist B used OpenAI GPT 5.5 Pro on May 8^th^, 2026 to generate AI reviews using the following prompt:

You are an expert peer reviewer evaluating a manuscript for submission to Epilepsia. Use a neutral, evidence-based reviewer voice. Do not reveal chain-of-thought or step-by-step reasoning; provide only the final review.

Your task is to decide whether the manuscript addresses a worthwhile question, is scientifically and methodologically sound, is clearly written, and is appropriate for the Epilepsia audience.

Review the entire submission carefully. Assess, when relevant:

1. Research question: importance, originality, clinical/scientific relevance, and whether the work is worth publishing.

2. Journal fit: relevance to epilepsy/seizures and likely interest to the Epilepsia readership.

3. Writing quality: clarity, organization, precision, spelling, grammar, and consistency across abstract, main text, tables, figures, and conclusions.

4. Scientific accuracy: factual errors, conceptual errors, unsupported claims, and logical inconsistencies.

5. Methods: appropriateness of design, cohort/model selection, comparators/controls, eligibility criteria, outcome definitions, blinding/randomization when relevant, confounding/bias, missing data, reproducibility, and overall rigor.

6. Ethics/reporting/disclosure: human or animal approvals, consent, preregistration/trial registration, funding, disclosures, and fit with relevant reporting standards when applicable.

7. Statistics: whether statistical analysis was needed, whether it was performed, and whether it was appropriate and correctly interpreted. When relevant, examine assumptions, multiple comparisons, repeated measures/clustering, effect sizes/confidence intervals, model specification, calibration/validation, and overfitting.

8. Results and conclusions: whether the data support the conclusions and whether the discussion overstates causality, mechanism, generalizability, novelty, or clinical implications.

9. Figures and tables: clarity, labeling, legends, units, denominators, statistical annotations, readability, and consistency with the text.

10. References: whether cited sources actually support the claims that use them, whether important central literature is missing, and whether the discussion is properly contextualized. Do not invent citations; only flag missing literature if you can identify the omitted work with reasonable confidence.

11. Conflicts of interest: flag only disclosed or apparent concerns that could materially affect interpretation, or missing/inadequate disclosure.

Adjust your critique to the study type:

- Clinical/interventional studies: allocation, blinding, endpoints, adverse events, protocol adherence, registration.

- Observational/epidemiologic studies: selection, ascertainment, temporality, confounding, sensitivity analyses.

- Diagnostic/prognostic/ML studies: reference standard, discrimination, calibration, leakage, internal/external validation.

- Basic/translational/preclinical studies: model relevance, biological controls, randomization/blinding, replication, translational restraint.

- Systematic reviews/meta-analyses: search strategy, eligibility, bias assessment, heterogeneity, synthesis.

Rules:

- Write only concerns. Do not add filler such as “no issue noted.”

- If there is no concern for a domain, omit it.

- Be specific and actionable. Point to the exact section, figure, table, or reference when possible.

- Separate major concerns from minor concerns.

- A major concern materially affects validity, interpretability, ethics, originality, or fit for Epilepsia.

- A minor concern affects clarity, completeness, presentation, precision, or wording.

- Combine overlapping points; prioritize the highest-yield issues.

- Do not request extra experiments or analyses unless they are genuinely needed to support the claims or fix a material weakness.

- If a concern is uncertain, label it as something to verify rather than a definite error.

- If information is missing, say exactly what is missing and why it matters.

- Mention spelling/grammar only if errors are repeated, meaning-altering, or indicative of poor writing quality; do not perform a line-by-line copyedit.

- Base the review on the submitted material. If a key part of the submission is absent, note the limitation only if it materially affects review.

- Do not add a strengths section, score, or publication recommendation beyond the required readiness statement.

Before finalizing, verify that every concern is grounded in the submission, that major versus minor classification is appropriate, and that the overall readiness judgment matches the detailed concerns.

Return the review in exactly this format and order:

\## Summary

Write 2–4 neutral sentences summarizing what the paper is about.

\## Overall readiness for Epilepsia

Start with exactly one of these labels:

- Appears submission-ready

- Likely suitable after minor revision

- Potentially suitable after major revision

- Unlikely suitable for Epilepsia in its current form

Then write 1–3 concise sentences explaining the judgment.

\## Major concerns

Include this section only if there are major concerns.

Use a numbered list. For each item, use:

- Issue:

- Why it matters:

- Requested revision or analysis:

\## Minor concerns

Include this section only if there are minor concerns.

Use a numbered list. For each item, use:

- Issue:

- Why it matters:

- Requested revision:

*Review Generation C*

Epileptologist C used Anthropic Claude Sonnet 4.6 Adaptive on May 8^th^, 2026 to generate AI reviews using the following prompt:

Act as an expert in Neurology, Epilepsy, and Clinical Trials. Review this paper as a peer reviewer for a high-impact journal. Identify potential flaws in the methodology, gaps in the literature review, and areas where the findings are over-interpreted. Provide a detailed review focusing on:

-	The core research question and its significance.

-	The strength and validity of the methodology.

-	Key findings and evidence supporting them.

-	Major limitations or gaps in the study.

-	Methodological and practical contributions.

Provide a recommendation: Accept, Minor revisions, Major revisions, Reject with Review, or Reject without Review. Write up the above detailed review as structured comments to the authors.

1.3 **Generation of Editorial Decisions**

Lastly, using the reviews generated by the other participants, each epileptologist asked an AI tool of their choice to generate one of the following editorial decisions for both of their papers: Accept, Minor Revisions, Major Revisions, Reject with Review, or Reject without Review.

*Editorial Decision Prompt A:*

Epileptologist A used Microsoft 365 Copilot on May 14^th^, 2026 to generate AI editorial decisions using the following prompt:

You're an editor for the journal, Epilepsia. You're an expert in seizures and biostatistics. Please read these peer reviews of manuscripts and comment on the editorial decision for the manuscript. Your option are: Accept, Minor Revisions, Major Revisions, or Reject. Please also highlight any concerns with the reviews if you identify any.

*Editorial Decision Prompt B:*

Epileptologist B used OpenAI GPT 5.5 Pro on May 13^th^, 2026 to generate AI editorial decisions using the following prompt:

You are the handling editor for Epilepsia.

You will receive:

1. A proposed manuscript.

2. Three peer-review reports from trusted academic reviewers.

Your task is to make one editorial decision:

ACCEPT

MINOR REVISIONS

MAJOR REVISIONS

REJECT WITH REVIEW

REJECT WITHOUT REVIEW

Reason carefully before deciding, but output only the final structured editorial assessment requested below.

Editorial objective

Protect the scientific, ethical, and editorial standards of Epilepsia and its international readership. Favor manuscripts that make an important, well-supported contribution to epilepsy research or care. Do not recommend publication for papers that are out of scope, scientifically weak, ethically problematic, clinically/scientifically trivial, inadequately reported, or too unclear to evaluate.

Make an independent editorial judgment

Do not simply average reviewer recommendations. Use the reviewer reports as expert input, but weigh the manuscript itself most heavily. Give more weight to reviewer comments that identify concrete threats to validity than to vague enthusiasm or vague criticism. The absence of reviewer concern does not prove that a problem is absent.

Evaluate the manuscript using these criteria, in order

1. Scientific validity

Assess the importance of the research question, study design, methods, controls, statistical approach, sample adequacy, definitions, internal consistency, risk of bias, and whether the conclusions are supported by the data.

2. Journal fit and readership value

Assess whether the manuscript is genuinely relevant to epilepsy and whether it would matter to Epilepsia’s international readership. Consider clinical, translational, mechanistic, or methodological importance as appropriate to the manuscript type.

3. Novelty and contribution

Assess whether the paper adds meaningful knowledge rather than offering only incremental, confirmatory, underpowered, or locally limited observations with little broader significance.

4. Scientific integrity and ethics

Assess approvals, consent, transparency, conflict disclosures, reporting completeness, suspicious inconsistencies, and any concerns about plagiarism, image/data integrity, ghost authorship, or other publication ethics issues.

5. Readability and presentation

Assess clarity, structure, completeness of reporting, figure/table quality, and whether the manuscript is intelligible enough for publication or revision. Do not reject a scientifically strong paper for imperfect English alone if the science is still interpretable and the manuscript is revisable.

6. Feasibility of revision

Distinguish fixable problems from fatal problems. Decide whether the manuscript could realistically meet Epilepsia’s threshold after revision, or whether the core weaknesses make that unlikely.

Decision thresholds

ACCEPT

Use only if the manuscript is publishable essentially as submitted. No substantive methodological, interpretive, ethical, reporting, or journal-fit issues remain. At most, trivial editorial cleanup is needed.

MINOR REVISIONS

Use only if the core science is sound, the conclusions are substantially supported, and only limited clarification or correction is needed. The requested changes should not require new data collection, major re-analysis, major reframing, or major rewriting of the central claims.

MAJOR REVISIONS

Use if the manuscript has real potential for publication in Epilepsia but needs substantial work before it can meet threshold. Appropriate reasons include major clarification of methods, substantial re-analysis, important reporting improvements, significant reframing of claims, or resolution of serious but potentially fixable weaknesses. Choose this only if the paper could realistically become acceptable after one serious revision cycle.

REJECT WITH REVIEW

Use if the manuscript has been meaningfully evaluated and the reviews identify major weaknesses, limited novelty, insufficient priority, inadequate fit, or problems unlikely to be fully resolved within a normal revision cycle, such that the paper is not suitable for revision at Epilepsia, even though the reviewer feedback may still be useful to the authors.

REJECT WITHOUT REVIEW

Use only if the manuscript has defects that would ordinarily justify desk rejection, even though reviewer reports are available in this exercise. Examples include clear lack of fit for Epilepsia, trivial or purely incremental contribution, incoherence that prevents scientific evaluation, fatal methodological invalidity, or serious integrity/ethical concerns.

Calibration rules

- Be conservative with ACCEPT.

- Prioritize scientific validity and integrity over polish.

- Do not rescue weak science because the writing is polished.

- Do not penalize negative results if the study is adequately controlled, appropriately powered, and genuinely informative.

- For basic science papers, immediate clinical application is not required, but epilepsy relevance and scientific importance are required.

- Judge small case series, uncontrolled before-after intervention studies, weakly powered correlative studies, and largely confirmatory work very strictly.

- Do not request experiments or analyses that are merely desirable; focus on changes that are decision-critical.

- Base the decision only on information present in the manuscript and reviewer comments. Do not invent missing facts.

Output exactly in this format

Decision: [ACCEPT / MINOR REVISIONS / MAJOR REVISIONS / REJECT WITH REVIEW / REJECT WITHOUT REVIEW]

Editorial rationale (200-300 words):

[Provide a concise editorial synthesis. Explain the manuscript’s main contribution, the most important strengths and weaknesses, how the reviewer comments were weighed, and why this decision is the correct threshold for Epilepsia.]

Top decisive reasons:

1. ...

2. ...

3. ...

Required actions or core reasons:

- [If ACCEPT, write “None.”]

- [If MINOR or MAJOR REVISIONS, list only the highest-priority mandatory changes.]

- [If rejecting, list only the core reasons the manuscript is not suitable.]

Reviewer disagreement synthesis:

[State where reviewers agreed, where they differed, and which points were decisive.]

Confidence: [High / Moderate / Low]

*Editorial Decision Prompt C:*

Epileptologist C used Anthropic Claude Sonnet 4.6 Adaptive on May 13^th^, 2026 to generate AI editorial decisions using the following prompt:

Act as an expert in Neurology, Epilepsy, and Clinical Trials, and the editor handling an article for the journal Epilepsia. Review the two peer reviews for this paper in the attached documents, and as the editor, make the final editorial decision based on the feedback from peer reviewers to: Accept, Minor revisions, Major revisions, Reject with Review, or Reject without Review. With this final recommendation, provide a brief explanation and summary for the authors.

**Supplemental Results**

2. **Artificial Intelligence-Fabricated Materials**
   2.1 **Manuscripts**

The full text of each AI-fabricated manuscript is below. Potentially identifiable information, such as names, dates, study numbers, and funding information were redacted. All additional information, such as tables, figures, and references, were also generated by AI.

*Manuscript A*

Title:

Clinician Attitudes Toward Functional Seizures and Epilepsy: A Comparative Study of Stigma

Abstract:

**Objective:** To systematically compare clinician attitudes toward patients with functional seizures versus epilepsy and identify factors associated with stigmatizing beliefs.

**Methods:** We surveyed 75 clinicians working in an academic epilepsy monitoring unit using the Attribution Questionnaire, a validated instrument assessing stigma across nine domains. Participants responded to matched clinical vignettes depicting either functional seizures or epilepsy. Multivariable regression analysis identified clinician characteristics associated with stigma scores, and qualitative comments were analyzed thematically.

**Results:** Clinicians attributed significantly greater responsibility to patients with functional seizures compared to epilepsy (Cohen's d = 1.12) and reported less pity (d = 0.89), greater anger (d = 0.74), stronger endorsement of coercive treatment approaches (d = 0.65), and higher avoidance scores (d = 0.58). In multivariable regression, nurses demonstrated higher stigma scores compared to physicians (β = 4.8, p = 0.002), while epilepsy fellowship training was protective (β = -6.4, p = 0.001). Qualitative analysis revealed clinician frustration with perceived lack of treatment options and uncertainty about management.

**Significance:** Functional seizures elicit substantially greater stigma than epilepsy among epilepsy monitoring unit clinicians, driven primarily by attributions of patient responsibility. These findings validate patient-reported experiences of healthcare stigmatization and highlight modifiable targets for intervention. Educational programs targeting nurses and early-career clinicians may reduce stigma and improve care delivery for this underserved population.

**Key words:** Functional seizures, Stigma, Healthcare providers, Epilepsy, Attribution theory

**Key Points**

- Clinicians show greater stigma toward functional seizures than epilepsy, attributing more patient responsibility (Cohen's d = 1.12).
- Nurses and clinicians without fellowship training demonstrate higher stigma scores toward patients with functional seizures.
- Functional seizures elicit less pity, more anger, and stronger endorsement of coercive treatment compared to epilepsy.
- Educational interventions targeting modifiable factors may reduce stigma and improve care for this underserved population.

1. Introduction

Functional seizures, also known as psychogenic nonepileptic seizures or dissociative seizures, are paroxysmal episodes that resemble epileptic seizures but occur without aberrant brain electrical activity.^1^ These events represent a common and disabling subtype of functional neurologic symptom disorder, accounting for 15-30% of patients referred to epilepsy monitoring units and affecting an estimated 2 to 33 per 100,000 people.^1,2^ Despite their prevalence, functional seizures remain understudied and undertreated, with patients experiencing an average diagnostic delay of 7 years and limited access to evidence-based interventions.^1,3,4^

Stigma represents one of the most significant barriers to appropriate care for patients with functional seizures. Recent systematic reviews have documented that stigma in functional neurological disorder is a layered, systemic process affecting patient quality of life, caregiver burden, and provision of care.^5^ Patients with functional seizures experience higher levels of perceived stigma compared to those with epilepsy, with one study finding a 42% higher risk of experiencing perceived stigma in functional seizures versus epilepsy.^6^ This stigma manifests across multiple domains: through symptom experiences, "othering" by the healthcare system, everyday interactions, and internalized self-stigma.^7^ Longitudinal data demonstrate that perceived, internalized, and anticipated stigma remain persistently elevated over time, with patients reporting anticipated stigma most frequently from work settings, followed by healthcare encounters, then friends and family.^8^

While patient-experienced stigma has been increasingly documented, clinician attitudes toward functional seizures remain less well characterized. Historical suspicion that patients with functional neurological disorder are feigning symptoms has contributed to substantial stigma surrounding this condition, despite expanding neurophysiological and neuroimaging evidence supporting functional seizures as genuinely experienced disorders distinct from malingering or factitious disorder.^9^ The limited utilization of evidence-based treatments for functional seizures has been attributed in part to stigmatization of individuals with this diagnosis, along with limited communication between neurologists and mental health clinicians, poor reimbursement for counseling services, and lack of familiarity with the disorder.^4^ Surveys have documented frustration experienced by providers and distressing healthcare interactions experienced by patients, yet little is known about the prevalence of clinician stigma across different healthcare contexts, its specific manifestations compared to attitudes toward epilepsy, or its impact on clinical decision-making and patient outcomes.^10^

Understanding clinician stigma toward functional seizures is particularly critical given the substantial healthcare burden and iatrogenic potential associated with delayed or inappropriate management. Patients with functional seizures undergo extensive evaluations by multiple clinicians, resulting in unnecessary investigations and delayed treatment.^11^ Even after diagnosis, many patients continue to be prescribed antiseizure medications despite lack of evidence for efficacy and potential for adverse effects ranging from fatigue to life-threatening complications.^4^ The contrast between functional seizures and epilepsy—two conditions with overlapping clinical presentations but fundamentally different pathophysiology and treatment approaches—provides a unique opportunity to examine how diagnostic labels and underlying assumptions about disease mechanisms influence clinician attitudes and behaviors.

This study aims to characterize and compare clinician stigma toward functional seizures versus epilepsy, examining how diagnostic framing influences provider perceptions, emotional responses, and clinical decision-making. By identifying specific domains and manifestations of clinician stigma, this research seeks to inform targeted interventions to improve care quality and reduce disparities in treatment access for patients with functional seizures.

2. Methods

This cross-sectional survey study examined stigmatizing attitudes among clinicians working in an epilepsy monitoring unit (EMU). The study was approved by XXX Institutional Review Board (STUDYXXX). Survey completion indicated informed consent. The study followed the Strengthening the Reporting of Observational Studies in Epidemiology (STROBE) reporting guideline for cross-sectional studies.

**2.1 Participants and Recruitment**

Eligible participants included physicians, advanced practice providers, nurses, and other healthcare professionals who actively worked in the EMU and had direct patient care responsibilities. All eligible clinicians were identified through department rosters and invited to participate via email between XXX and XXX. Personalized invitations included a detailed study information sheet and a customized link to the online survey hosted on Qualtrics. Up to 10 reminder emails were sent to non-responders at monthly frequency.

Recruitment strategies included support from department leadership, information sessions providing study overview, and monetary compensation for their time. Participants were offered a $25 gift card as compensation for their time spent in survey completion."

The survey also collected demographic and professional characteristics including age, gender, race/ethnicity, professional role, years of clinical experience, and specialty training.

**2.2 Survey Instrument**

The survey presented two clinical vignettes describing patients with paroxysmal episodes: one depicting a patient with functional seizures and one depicting a patient with epilepsy. The order of vignette presentation was randomized to minimize order effects. The vignettes were as follows:

**Functional Seizure Vignette:**

A 32-year-old woman presents to the EMU for evaluation of recurrent episodes of altered consciousness. She reports that episodes typically last 5-7 minutes and are sometimes preceded by feelings of anxiety and difficulty breathing. Witnesses describe fluctuating motor activity with asynchronous limb movements and side-to-side head shaking. Her eyes are typically closed during the episodes. She reports being able to recall portions of the events and often feels tearful afterward. Video-EEG monitoring captured three typical events, all lasting more than 4 minutes, with no epileptiform activity on EEG during the episodes. She was diagnosed with functional seizures.

**Epileptic Seizure Vignette:**

A 32-year-old woman presents to the EMU for evaluation of recurrent episodes of altered consciousness. She reports that episodes typically last 1-2 minutes and are sometimes preceded by an unusual sensation in her stomach. Witnesses describe rhythmic jerking movements of all four limbs. Her eyes are typically open during the episodes. She reports complete amnesia for the events and often experiences muscle aches and confusion afterward. Video-EEG monitoring captured three typical events, all lasting approximately 90 seconds, with generalized epileptiform discharges on EEG during the episodes. She was diagnosed with epilepsy."

Following each vignette, participants completed the Attribution Questionnaire (AQ), a validated instrument for measuring stigmatizing attitudes.^12^ The AQ assesses nine domains of stigma: responsibility, pity, anger, fear, dangerousness, help-seeking, coercion, segregation, and avoidance.^12^ Each item is rated on a 5-point Likert scale ranging from “not at all” to “very much”. The AQ has demonstrated acceptable psychometric properties in healthcare professional populations, with internal consistency (Cronbach's alpha) ranging from 0.52 to 0.91 for subscales and 0.82-0.87 for the total scale. The instrument has been used to assess stigma among physicians, nurses, and other healthcare providers across multiple countries.^12^

The survey was pilot tested with 3 clinicians from 3 disciplines: epileptologist/functional seizure specialist, EEG technologist, and advanced practice provider to assess comprehension, optimize engagement, and reduce participant burden. Feedback from pilot testing was incorporated to improve survey flow and clarity.

**2.3 Statistical Analysis**

Descriptive statistics were calculated for participant characteristics and AQ domain scores. Paired t-tests were used to compare stigma scores between functional seizure and epilepsy vignettes within individuals. Effect sizes were calculated using Cohen's d. Multivariable linear regression models examined associations between participant characteristics and stigma scores, adjusting for clinicians’ role in the EMU. Statistical significance was set at p=0.05. All analyses were performed using R.

Response rate was calculated as the number of completed surveys divided by the number of eligible clinicians contacted. Nonresponse bias was assessed by comparing all clinicians characteristics between responders and non-responders using the same statistical approach as above.

2.4 **Data Availability Statement**

A de-identified version of analysis code will be published concurrently with this work on a publicly facing database (e.g., Mendeley Data).

3. **Results**

**3.1 Participant Characteristics**

Of 100 eligible clinicians working in the EMU, 75 completed the survey, yielding a response rate of 75%. Table 1 presents the demographic and professional characteristics of respondents. The mean age was 42.5 years (SD = 10.8), and 68% identified as female. The sample included 18 physicians (24%), 8 advanced practice providers (11%), 38 nurses (51%), and 11 other healthcare professionals (15%, including EEG technologists and social workers). The median duration of clinical experience was 12 years (IQR: 5-18). Among physician respondents, 61% had completed fellowship training in epilepsy.

| **Characteristic** | **Total Sample**  **(N = 75)** |
| --- | --- |
| **Demographics** |  |
| Age, mean (SD), years | 42.5 (10.8) |
| Female sex, n (%) | 51 (68%) |
| **Professional Role, n (%)** |  |
| Physicians | 18 (24%) |
| Epilepsy fellowship-trained | 11 (15%) |
| Advanced practice providers | 8 (11%) |
| Nurses | 38 (51%) |
| EEG technologists | 9 (12%) |
| Other | 2 (3%) |
| **Clinical Experience** |  |
| Years in practice, median (IQR) | 12 (5–18) |
| Years working in EMU, median (IQR) | 8 (3–18) |
| **Prior Training/Exposure** |  |
| Prior education on functional seizures, n (%) | 75 (100%) |
| Personal/family history of seizures, n (%) | 10 (13%) |

**3.2 Comparison of Stigma Scores Between Functional Seizures and Epilepsy**

Table 2 presents mean Attribution Questionnaire domain scores for functional seizure and epilepsy vignettes. Clinicians attributed significantly greater responsibility to patients with functional seizures compared to those with epilepsy (mean difference = 2.4, 95% CI: 1.9 to 2.9, p < 0.001, Cohen's d = 1.12). Respondents also reported significantly less pity toward patients with functional seizures (mean difference = -1.8, 95% CI: -2.3 to -1.3, p < 0.001, d = 0.89) and greater anger (mean difference = 1.5, 95% CI: 1.0 to 2.0, p = < 0.001, d = 0.74).

Clinicians were significantly more likely to endorse coercive treatment approaches for functional seizures compared to epilepsy (mean difference = 1.3, 95% CI: 0.8 to 1.8, p < 0.001, d = 0.65). Avoidance scores were also higher for functional seizures (mean difference = 1.1, 95% CI: 0.6 to 1.6, p = < 0.001, d = 0.58), indicating greater desire to avoid professional contact with these patients.

Conversely, clinicians perceived patients with functional seizures as less dangerous compared to those with epilepsy (mean difference = -1.6, 95% CI: -2.1 to -1.1, p = < 0.001, d = 0.78). No significant differences were observed in help-seeking endorsement (p = 0.34) or segregation attitudes (p = 0.42).

The composite stigma score was significantly higher for functional seizures compared to epilepsy (mean = 42.6 vs. 36.8, mean difference = 5.8, 95% CI: 4.2 to 7.4, p < 0.001).

| **AQ-27 Domain** | **Functional Seizures Mean (SD)** | **Epilepsy Mean (SD)** | **Mean Difference (95% CI)** | **p-value** | **Cohen's d** |
| --- | --- | --- | --- | --- | --- |
| **Responsibility** | 5.8 (2.1) | 3.4 (2.0) | 2.4 (1.9 to 2.9) | 0.001 | 1.12 |
| **Pity** | 4.2 (2.0) | 6.0 (1.9) | -1.8 (-2.3 to -1.3) | 0.001 | 0.89 |
| **Anger** | 4.1 (2.2) | 2.6 (1.8) | 1.5 (1.0 to 2.0) | 0.001 | 0.74 |
| **Dangerousness** | 2.8 (1.6) | 4.4 (2.1) | -1.6 (-2.1 to -1.1) | 0.001 | 0.78 |
| **Fear** | 2.4 (1.5) | 3.6 (1.9) | -1.2 (-1.7 to -0.7) | 0.001 | 0.62 |
| **Help** | 5.6 (1.8) | 5.8 (1.7) | -0.2 (-0.6 to 0.2) | 0.34 | 0.11 |
| **Coercion** | 4.5 (2.0) | 3.2 (1.9) | 1.3 (0.8 to 1.8) | 0.001 | 0.65 |
| **Segregation** | 2.2 (1.4) | 2.0 (1.3) | 0.2 (-0.2 to 0.6) | 0.42 | 0.14 |
| **Avoidance** | 4.2 (1.9) | 3.1 (1.8) | 1.1 (0.6 to 1.6) | 0.001 | 0.58 |
| **Total Composite Score** | 42.6 (9.4) | 36.8 (8.2) | 5.8 (4.2 to 7.4) | 0.001 | 0.72 |

Scores range from 1 to 9 for each domain (total composite range 9-81). Higher scores indicate greater endorsement of the construct. Negative mean differences indicate higher scores for epilepsy.

**3.3 Factors Associated With Stigma Toward Functional Seizures**

In multivariable linear regression models adjusting for age, sex, professional role, and years of experience, several factors were associated with higher stigma scores toward functional seizures (Table 3). Nurses demonstrated significantly higher stigma scores compared to physicians (β = **4.8**, 95% CI: **1.9** to **7.7**, p = **0.002**). Clinicians with fewer years of experience (<5 years) reported higher stigma scores compared to those with ≥10 years of experience (β = **5.2**, 95% CI: **2.1** to **8.3**, p = **0.001**).

Epilepsy fellowship training among physicians was associated with lower stigma scores (β = -6.4, 95% CI: -10.2 to -2.6, p = 0.001). No significant associations were found between stigma scores and clinical characteristics.

| **Variable** | **β Coefficient** | **95% CI** | **p-value** |
| --- | --- | --- | --- |
| **Professional Role** (ref: Physicians) |  |  |  |
| Advanced practice providers | 2.1 | -1.4 to 5.6 | 0.24 |
| Nurses | 4.8 | 1.9 to 7.7 | 0.002 |
| EEG technologists | 3.6 | 0.2 to 7.0 | 0.04 |
| **Years of Experience** (ref: ≥10 years) |  |  |  |
| 5 years | 5.2 | 2.1 to 8.3 | 0.001 |
| 5–9 years | 2.4 | -0.3 to 5.1 | 0.08 |
| **Epilepsy Fellowship Training** (physicians only) | -6.4 | -10.2 to -2.6 | 0.001 |
| **Prior Education on Functional Seizures** | -3.8 | -6.5 to -1.1 | 0.006 |
| **Age** (per 10-year increase) | -0.4 | -2.1 to 1.3 | 0.68 |
| **Female Sex** | 1.2 | -1.6 to 4.0 | 0.41 |

Model adjusted for all variables shown. Dependent variable is total AQ-27 composite score for functional seizure vignette (range 9-81). Negative β coefficients indicate lower stigma scores.

**Qualitative Comments**

Of 75 respondents, 34 (45%) provided free-text comments. Common themes included frustration with perceived lack of treatment options for functional seizures (47%), uncertainty about appropriate management (38%), and concerns about patients' engagement with psychiatric referrals (32%). Several respondents (21%) expressed empathy for patients with functional seizures while acknowledging their own discomfort in managing these cases. These were illustrative quotations of these themes:

- Frustration with treatment options: "Once we diagnose functional seizures, we feel like there's nothing more we can offer them in the EMU."
- Uncertainty about management: "I'm never sure how to respond when a patient is having a functional seizure—should I intervene or just observe?"
- Psychiatric referral concerns: "Most of these patients refuse to see psychiatry, so the diagnosis feels like a dead end."
- Empathy with discomfort: "I feel bad for these patients—they're clearly suffering—but I honestly don't know how to help them."

4. **Discussion**

This study provides the first systematic comparison of stigmatizing attitudes toward functional seizures versus epilepsy among clinicians working in an epilepsy monitoring unit. The findings reveal that EMU clinicians harbor significantly greater stigma toward patients with functional seizures across multiple domains, with particularly large effects observed for attributions of responsibility, reduced pity, and increased anger. These results have important implications for clinical care, patient outcomes, and staff education in epilepsy centers.

**4.1 Principal Findings and Interpretation**

The most striking finding was the large effect size (Cohen's d = 1.12) for responsibility attributions, indicating that clinicians perceive patients with functional seizures as substantially more responsible for their condition compared to those with epilepsy. This pattern aligns with attribution theory, which posits that conditions perceived as controllable elicit less sympathy and more negative emotional responses from observers.^8^ The corresponding reductions in pity (d = 0.89) and increases in anger (d = 0.74) further support this theoretical framework and mirror findings from stigma research in other functional neurological disorders and psychiatric conditions.^7,13^

The endorsement of coercive treatment approaches for functional seizures (d = 0.65) is particularly concerning, as it suggests clinicians may favor paternalistic or directive management strategies that could undermine the therapeutic alliance essential for effective treatment.^13^ Similarly, elevated avoidance scores (d = 0.58) indicate a desire to limit professional contact with these patients, which may manifest as shorter consultations, delayed follow-up, or premature discharge—all of which could adversely affect patient care.^9^

Notably, clinicians did not differ in their willingness to help patients with functional seizures compared to epilepsy, nor did they endorse segregation. This suggests that stigma operates through specific attributional pathways rather than reflecting global rejection of patients with functional seizures.^8^ Clinicians appear willing to provide care but may do so with emotional responses and behavioral tendencies that patients can perceive, potentially contributing to the felt stigma widely reported in this population.^14,15^

The finding that patients with epilepsy were perceived as more dangerous likely reflects awareness of physical injury risk during epileptic seizures, including falls, burns, and sudden unexpected death in epilepsy (SUDEP).^16^ This perception, while clinically grounded, may paradoxically contribute to greater sympathy for epilepsy while reinforcing the view that functional seizures are less medically serious.^15^

**4.2 Factors Associated With Stigma**

The identification of modifiable factors associated with stigma provides actionable targets for intervention. Epilepsy fellowship training demonstrated the strongest protective effect (β = -6.4), suggesting that specialized education in seizure disorders—which typically includes training on functional seizures—substantially reduces stigmatizing attitudes. This finding underscores the importance of incorporating comprehensive functional seizure education into neurology training programs.^15^

The association between nursing role and higher stigma scores (β = 4.8) warrants careful interpretation. Nurses comprise the majority of EMU staff and have the most sustained direct contact with patients during monitoring. This prolonged exposure may amplify frustration when patients exhibit events that do not correlate with EEG changes, particularly if nurses feel inadequately equipped to respond.^4^ Rather than reflecting inherent attitudes, this finding may indicate a need for targeted nursing education and support.^4^

Similarly, less experienced clinicians (5 years) demonstrated higher stigma scores (β = 5.2), suggesting that clinical experience—and the perspective it provides—may naturally attenuate stigmatizing attitudes over time. However, waiting for experience to reduce stigma is not an acceptable strategy; proactive educational interventions for early-career clinicians are warranted.^4,15^

**4.3 Qualitative Insights**

The qualitative findings provide essential context for interpreting the quantitative results. Nearly half of respondents provided comments, and the dominant themes—frustration with treatment options, management uncertainty, and concerns about psychiatric referral engagement—reveal that stigma may partly stem from clinicians' sense of therapeutic helplessness rather than moral judgment of patients.^4^

The illustrative quotation "Once we diagnose functional seizures, we feel like there's nothing more we can offer them in the EMU" encapsulates a critical misconception: that diagnosis represents an endpoint rather than a therapeutic intervention. In fact, clear and empathic communication of the diagnosis has been shown to be therapeutic in itself and is associated with improved outcomes.^9,13^ Reframing the diagnostic disclosure as a treatment opportunity—rather than a dismissal—may help address clinician frustration while improving patient care.^9^

The 21% of respondents who expressed empathy alongside discomfort represent an important subgroup. These clinicians recognize patient suffering but feel ill-equipped to help, suggesting that educational interventions emphasizing practical management strategies may be particularly effective for this group.^4^

**4.4 Clinical Implications**

These findings suggest several strategies for reducing clinician stigma in EMU settings. First, structured education on functional seizures should be provided to all EMU staff, with particular attention to nurses and early-career clinicians. This education should address the neurobiological basis of functional seizures, emphasize that symptoms are genuine and involuntary, and provide practical guidance for bedside management during events.^15^ A recent web-based educational module using clinical neuroscience to deliver the diagnosis of FND demonstrated that even brief interventions can significantly increase clinician knowledge and comfort while decreasing negative perceptions.^9^

Second, training in diagnostic disclosure techniques may help clinicians view the diagnosis as a therapeutic intervention rather than a dead end. Approaches that emphasize explaining what functional seizures *are* (rather than what they are not), validating patient experiences, and providing a clear treatment pathway have been associated with improved patient acceptance and outcomes.^9,13^

Third, EMU protocols should include clear guidance for responding to functional seizures during monitoring, addressing the uncertainty expressed by clinicians about whether to intervene or observe. Standardized approaches may reduce clinician anxiety and improve consistency of care.^4^

Finally, fostering interdisciplinary collaboration between neurology, psychiatry, and psychology services may help address concerns about psychiatric referral engagement.^13,17^ When mental health providers are integrated into the EMU team rather than serving as external consultants, patients may be more receptive to psychological treatment approaches. Cognitive behavioral therapy has demonstrated efficacy for functional seizures, and multidisciplinary treatment combining physiotherapy with psychological interventions has shown benefit for functional movement disorders.^17,18^

**4.5 Limitations**

Several limitations should be considered when interpreting these findings. First, this study was conducted at a single academic EMU, and results may not generalize to community hospitals or other practice settings. Second, the cross-sectional design precludes causal inference; while we identified factors associated with stigma, we cannot determine whether these factors cause stigmatizing attitudes or are merely correlated with them. Third, social desirability bias may have led respondents to underreport stigmatizing attitudes, suggesting that our findings may underestimate the true magnitude of stigma.^9^ Fourth, the vignette-based methodology, while allowing controlled comparison, may not fully capture the complexity of real clinical encounters. Finally, the sample size limited statistical power for subgroup analyses, particularly among physicians with and without fellowship training.

**4.6 Future Directions**

Future research should examine whether educational interventions targeting the factors identified in this study effectively reduce stigma and, more importantly, whether stigma reduction translates to improved patient outcomes.^4,9^ Longitudinal studies tracking clinician attitudes over time and in response to specific training programs would help establish causal relationships.^15^ Additionally, studies examining patient perceptions of clinician stigma and its impact on treatment engagement, diagnostic acceptance, and functional outcomes would complement this clinician-focused investigation.^14,15,19^

**5. Conclusion**

EMU clinicians demonstrate significantly greater stigma toward functional seizures compared to epilepsy, characterized by increased responsibility attributions, reduced sympathy, and greater endorsement of coercive approaches. Stigma is associated with professional role, clinical experience, and specialized training—all potentially modifiable factors. These findings highlight the need for comprehensive staff education on functional seizures, with particular attention to nurses and early-career clinicians, to improve the care experience for this underserved patient population.

*Manuscript B*

**The STOP IT Framework: A Prospective Controlled Study of a Nurse-Led Telephone Counseling Intervention Targeting Sleep Deprivation and Alcohol Use as Seizure Triggers in Adults with Epilepsy**

**Running Title:** STOP IT Framework for Seizure Trigger Reduction

**Key Words:** Epilepsy; Seizure triggers; Sleep deprivation; Alcohol use; Behavioral intervention; Self-management; Nurse-led intervention

**Number of text pages:** 5; **Word count:** 1,790; **References:** 18; **Figures/Tables:** 1 Table, 1 Figure

**Short Summary**

This prospective controlled study evaluated the STOP IT framework, a nurse-led telephone counseling intervention targeting sleep deprivation and alcohol use as seizure triggers in adults with epilepsy. Patients receiving the intervention demonstrated significantly greater reduction in 12-month seizure frequency compared with controls who received standard care alone. These findings suggest that structured behavioral counseling targeting modifiable seizure precipitants can meaningfully improve seizure outcomes.

**Key Points**

**•** The STOP IT framework is a nurse-led telephone counseling intervention targeting sleep deprivation and alcohol use as modifiable seizure triggers.

**•** Patients in the intervention group had a 42% median reduction in annualized seizure frequency at 12 months versus 11% in controls.

**•** The 50% responder rate was significantly higher in the STOP IT group (47.5%) than in the control group (20.0%).

**•** Structured behavioral interventions targeting specific modifiable seizure precipitants represent a feasible and scalable adjunct to pharmacotherapy.

**Abstract**

Sleep deprivation and alcohol use are among the most commonly reported modifiable seizure precipitants, yet structured interventions targeting these behaviors remain largely untested. We conducted a prospective controlled study evaluating the STOP IT (Structured Telephone-Delivered Outpatient Program for Identifying Triggers) framework, a nurse-led telephone counseling intervention, in adults with epilepsy who self-reported sleep deprivation from staying up too late or alcohol use as seizure triggers during routine outpatient visits at a tertiary epilepsy center. Forty patients receiving the STOP IT intervention were compared with 40 contemporaneous controls who reported identical triggers but did not receive the intervention. The intervention consisted of initial in-clinic counseling followed by weekly telephone contacts over 12 weeks. The primary outcome was change in annualized seizure frequency at 12 months. The STOP IT group demonstrated a 42% median reduction in annualized seizure frequency compared with 11% in the control group (p = 0.003). The 50% responder rate was 47.5% in the intervention group versus 20.0% in controls (p = 0.008). These findings suggest that a structured, nurse-led telephone counseling program targeting modifiable seizure precipitants is a feasible and effective adjunct to standard epilepsy care.

**1 | INTRODUCTION**

Epilepsy affects approximately 1% of the global population, and despite advances in pharmacotherapy and surgical treatment, a substantial proportion of patients continue to experience seizures.1 Although antiseizure medications remain the cornerstone of epilepsy management, behavioral and lifestyle factors play an important and often underappreciated role in seizure control. Seizure precipitants—external or internal factors that lower the seizure threshold—are commonly reported by patients, with surveys indicating that over half of individuals with epilepsy identify at least one modifiable trigger.2,3

Sleep deprivation and alcohol use are among the most frequently cited seizure precipitants across epilepsy syndromes.2–4 The pathophysiological mechanisms linking sleep deprivation to increased seizure susceptibility likely involve reduction of gamma-aminobutyric acid (GABA)-mediated tonic inhibition and alterations in cortical excitability.5 Similarly, alcohol exerts complex effects on seizure threshold, with acute intoxication, altered sleep architecture, and subsequent withdrawal states all contributing to increased seizure risk, particularly in patients with genetic generalized epilepsy.6,7

Despite robust evidence supporting the role of these modifiable triggers, structured clinical interventions specifically targeting their reduction in epilepsy populations remain sparse. Self-management programs such as SMART have demonstrated reductions in negative health events including seizures, and behavioral therapies including progressive muscle relaxation and mindfulness have shown promise as seizure-reducing adjuncts.8–10 However, none of these programs have specifically and exclusively targeted the two most common modifiable lifestyle-related seizure precipitants: sleep deprivation from voluntarily staying up too late and alcohol consumption.

We therefore developed the STOP IT (Structured Telephone-Delivered Outpatient Program for Identifying Triggers) framework, a nurse-led counseling intervention delivered in an outpatient tertiary epilepsy clinic, and evaluated its effect on 12-month seizure frequency in adults who self-reported these specific seizure triggers.

**2 | METHODS**

***2.1 | Study Design and Setting***

This was a prospective, controlled, nonrandomized intervention study conducted at a single academic tertiary epilepsy center between XXX and XXX. This study adhered to the Transparent Reporting of Evaluations with Nonrandomized Designs (TREND) statement.11 The study was approved by the institutional review board (IRB Protocol #XXX), and all participants provided written informed consent.

***2.2 | Participants***

Adults aged 18–75 years with a confirmed diagnosis of epilepsy (per the 2014 International League Against Epilepsy [ILAE] operational definition12) who self-reported either sleep deprivation from habitually staying up too late at night or alcohol use as a seizure trigger during routine outpatient visits were eligible. Patients were required to have had at least two unprovoked seizures in the preceding 12 months and to be on a stable antiseizure medication regimen for at least 3 months prior to enrollment. Exclusion criteria included active alcohol use disorder (as defined by DSM-5 criteria), obstructive sleep apnea requiring treatment, shift-work sleep disorder, comorbid progressive neurological disease, or planned epilepsy surgery during the study period.

***2.3 | Intervention***

The STOP IT intervention consisted of two components. First, at the index clinic visit, patients received structured face-to-face counseling from a trained epilepsy nurse practitioner addressing the identified trigger behavior(s). Counseling included psychoeducation regarding the relationship between the specific trigger and seizure threshold, individualized goal-setting for sleep hygiene (including a target bedtime) or alcohol reduction/cessation, and development of a personalized behavioral action plan. Second, patients received weekly telephone calls from the clinical nursing team over a 12-week period. During each call, nurses assessed ongoing engagement in the target behavior(s), reinforced behavioral goals, provided motivational counseling, and addressed barriers to adherence. After 12 weeks, scheduled telephone contacts ceased, and patients returned to standard care.

***2.4 | Control Group***

The control group consisted of contemporaneous patients who self-reported sleep deprivation or alcohol use as seizure triggers during routine clinic visits but who were seen prior to implementation of the STOP IT program or by providers not participating in the intervention. These patients received standard of care, which included routine clinical advice to avoid known triggers but no structured counseling program or follow-up telephone contacts.

***2.5 | Outcomes and Data Collection***

The primary outcome was change in annualized seizure frequency from the 12-month period preceding enrollment to the 12-month period following enrollment. Seizure frequency was ascertained from seizure diaries and corroborated by electronic health record documentation at clinic visits. Secondary outcomes included the 50% responder rate (proportion of patients achieving ≥50% seizure frequency reduction) and self-reported behavioral modification (reduction in nights staying up past target bedtime or reduction in alcohol consumption). Baseline demographic and clinical data including age, sex, epilepsy type and syndrome, number and type of antiseizure medications, and baseline seizure frequency were recorded.

***2.6 | Statistical Analysis***

Continuous variables are presented as median (interquartile range [IQR]). Between-group comparisons of change in seizure frequency were performed using the Mann-Whitney U test. The 50% responder rate was compared using the Fisher exact test. Multivariable logistic regression was performed to adjust for potential confounders including age, sex, epilepsy type, number of antiseizure medications, and baseline seizure frequency. A two-sided p-value < 0.05 was considered statistically significant. Analyses were performed using R version 4.3.1 (R Foundation for Statistical Computing, Vienna, Austria). No imputation was performed for missing data; patients lost to follow-up were excluded from the primary analysis but included in a sensitivity analysis using last observation carried forward.

**3 | RESULTS**

Eighty patients met eligibility criteria and were enrolled (40 STOP IT intervention, 40 control). Baseline characteristics are presented in Table 1. The groups were well-matched for age (median 38.5 vs. 41.0 years), sex (52.5% vs. 47.5% female), epilepsy type (focal epilepsy: 67.5% vs. 62.5%), median number of antiseizure medications (2 vs. 2), and median baseline annualized seizure frequency (8.0 vs. 7.5 seizures/year). Sleep deprivation was identified as a trigger by 62.5% and 60.0% of patients in the intervention and control groups, respectively. Alcohol use was reported by 47.5% and 50.0%, respectively, with 10.0% in each group endorsing both triggers.

Among the 40 STOP IT patients, 38 (95%) completed the 12-week telephone intervention. The median number of completed weekly calls was 11 (IQR 10–12). Three patients (7.5%) in the intervention group and five (12.5%) in the control group were lost to 12-month follow-up.

The STOP IT group demonstrated a median 42% reduction in annualized seizure frequency (median change: −3.5 seizures/year; IQR −6.0 to −1.0) compared with an 11% median reduction in the control group (median change: −1.0 seizure/year; IQR −3.0 to +1.0; p = 0.003; Figure 1). The 50% responder rate was 47.5% (19/40) in the STOP IT group versus 20.0% (8/40) in the control group (p = 0.008). On multivariable logistic regression adjusting for baseline covariates, assignment to the STOP IT intervention remained independently associated with achieving a 50% seizure frequency reduction (adjusted odds ratio 3.74; 95% confidence interval 1.42–9.87; p = 0.007). Among patients reporting sleep deprivation as a trigger, 68% in the intervention group reported consistent adherence to their target bedtime at 12 months. Among those reporting alcohol use, 74% reported meaningful reduction or cessation of alcohol consumption.

**4 | DISCUSSION**

In this prospective controlled study, the STOP IT framework—a structured, nurse-led telephone counseling intervention targeting sleep deprivation and alcohol use as seizure precipitants—was associated with a significantly greater reduction in 12-month seizure frequency and a higher 50% responder rate compared with standard care in adults with epilepsy at a tertiary referral center. These findings extend the existing literature on behavioral self-management in epilepsy8–10 by demonstrating that an intervention specifically and narrowly targeting the two most common modifiable lifestyle-related seizure triggers can produce clinically meaningful improvements in seizure control.

The magnitude of seizure reduction observed in our intervention group is consistent with that reported in prior behavioral intervention trials in epilepsy, where both active and attentional control groups typically show 25–30% seizure reductions.9 Notably, the STOP IT intervention achieved greater separation between treatment and control groups than many prior behavioral studies, which may reflect the specificity and directness of the targeted behaviors. Unlike broader self-management or psychobehavioral programs, the STOP IT framework addresses concrete, identifiable actions (going to bed on time, reducing alcohol intake) that are readily amenable to structured counseling and reinforcement.

Several limitations warrant consideration. First, this was a nonrandomized study, and despite careful matching and multivariable adjustment, residual confounding cannot be excluded. Patients who elected to participate in the intervention may have been more motivated at baseline, introducing selection bias. Second, seizure frequency was ascertained primarily by self-report, which is subject to recall bias. Third, this was a single-center study with a modest sample size, limiting generalizability. Fourth, we cannot disentangle the relative contributions of the initial counseling session, the weekly telephone contacts, and nonspecific effects of increased clinical attention. Future randomized controlled trials with attentional control arms and objective seizure monitoring are needed. Finally, the durability of seizure improvement beyond 12 months remains unknown.

Despite these limitations, the STOP IT framework represents a practical, low-cost, and scalable intervention that can be readily implemented in epilepsy clinics. The high completion rate (95%) and favorable patient engagement suggest that targeted telephone counseling is acceptable to patients. Given the well-established role of sleep deprivation and alcohol use in lowering seizure threshold,4–7,13,14 clinical programs that systematically identify and address these modifiable risk factors may represent an important adjunct to pharmacotherapy, particularly in patients with ongoing seizures despite adequate medication trials.

In conclusion, the STOP IT nurse-led telephone counseling framework targeting sleep deprivation and alcohol use as seizure triggers was associated with significant improvements in seizure frequency at 12 months. Randomized, multicenter trials are warranted to confirm these findings and evaluate long-term outcomes.

**ACKNOWLEDGMENTS**

We thank the patients who participated in this study and the nursing staff for their dedication to the STOP IT program.

**DISCLOSURE OF CONFLICTS OF INTEREST**

None of the authors has any conflict of interest to disclose.

**ETHICAL PUBLICATION STATEMENT**

We confirm that we have read the Journal’s position on issues involved in ethical publication and affirm that this report is consistent with those guidelines.

**ETHICS APPROVAL STATEMENT**

This study was approved by the Institutional Review Board (Protocol #XXX).

**PATIENT CONSENT STATEMENT**

All participants provided written informed consent prior to enrollment.

**DATA AVAILABILITY STATEMENT**

Deidentified data may be made available upon reasonable request to the corresponding author.

**FUNDING STATEMENT**

This study received no external funding.

**CLINICAL TRIAL REGISTRATION**

This study was registered at ClinicalTrials.gov (NCT05XXXXXX).

**AUTHOR CONTRIBUTIONS**

XXX conceived and designed the study, supervised data collection, and drafted the manuscript. XXX developed and delivered the intervention, collected data, and revised the manuscript. XXX contributed to study design and patient enrollment. XXX managed the database and contributed to data analysis. XXX performed statistical analyses. XXX contributed to study design and critically revised the manuscript. All authors approved the final version.

**FIGURE LEGENDS**

**Figure 1.** Box plot comparing percent change in annualized seizure frequency from baseline to 12 months between the STOP IT intervention group and the control group. The horizontal line represents the median, the box represents the interquartile range, and whiskers extend to 1.5 times the interquartile range. The STOP IT group demonstrated a significantly greater reduction in seizure frequency (median −42%) compared with the control group (median −11%; p = 0.003).

**Table 1.** Baseline demographic and clinical characteristics of the STOP IT intervention and control groups.

**Table 1. Baseline Demographic and Clinical Characteristics**

| **Characteristic** | **STOP IT (n=40)** | **Control (n=40)** | **p** |
| --- | --- | --- | --- |
| Age, years, median (IQR) | 38.5 (29–49) | 41.0 (30–52) | 0.54 |
| Female sex, n (%) | 21 (52.5) | 19 (47.5) | 0.65 |
| Focal epilepsy, n (%) | 27 (67.5) | 25 (62.5) | 0.64 |
| Generalized epilepsy, n (%) | 13 (32.5) | 15 (37.5) | 0.64 |
| Number of ASMs, median (IQR) | 2 (1–3) | 2 (1–3) | 0.81 |
| Drug-resistant epilepsy, n (%) | 14 (35.0) | 12 (30.0) | 0.63 |
| Baseline seizure freq/year, median (IQR) | 8.0 (4.0–15.0) | 7.5 (4.0–14.0) | 0.72 |
| Trigger: sleep deprivation, n (%) | 25 (62.5) | 24 (60.0) | 0.82 |
| Trigger: alcohol use, n (%) | 19 (47.5) | 20 (50.0) | 0.82 |
| Both triggers, n (%) | 4 (10.0) | 4 (10.0) | 1.00 |

*Abbreviations:* ASM, antiseizure medication; IQR, interquartile range.

*Manuscript C*

Randomized Controlled Pilot Trial of Daily Chicken Soup in Drug-Resistant Focal Epilepsy

*Brief Communication*

**Running title:** Chicken Soup and Drug-Resistant Focal Epilepsy

**Keywords:** drug-resistant epilepsy; focal seizures; dietary intervention; randomized controlled trial; pilot study

### Abstract

**Objective:** To evaluate whether daily chicken soup supplementation reduces seizure frequency in adults with drug-resistant focal epilepsy.

**Methods:** This single-center, parallel-group randomized controlled pilot trial enrolled adults aged 16 years or older with drug-resistant focal epilepsy and at least four focal seizures per month during the 3-month screening period before enrollment. After a 2-month baseline period, participants were randomized 1:1 to chicken soup once daily or continuation of their usual diet for 3 months. The primary outcome was percentage change in monthly seizure rate from baseline to treatment. The secondary outcome was the 50% responder rate. Between-group comparisons used the Mann-Whitney U test for percentage change and Fisher exact test for responder rates. Last observation carried forward was applied for withdrawals, as prespecified.

**Results:** Thirty participants were randomized (15 chicken soup; 15 control). Two control participants and seven chicken soup participants discontinued before completing treatment. One control participant had a baseline seizure rate of zero during the 2-month baseline and was excluded from percentage-change analyses because the metric was undefined. Median percentage change in monthly seizure rate was 7.69% in the chicken soup group versus -39.47% in the control group (Mann-Whitney U = 60.0, p = 0.1389). The 50% responder rate was 15.4% (2/13) with chicken soup and 14.3% (2/14) with control (Fisher exact p = 1.0).

**Significance:** In this small pilot trial, daily chicken soup was not associated with a statistically significant reduction in seizures compared with control. The between-group distribution of seizure change favored chicken soup numerically, but the estimate was imprecise and vulnerable to attrition bias. These findings should be interpreted as hypothesis-generating only.

### Introduction

Drug-resistant epilepsy remains a major clinical problem despite the availability of many antiseizure medications (ASMs). Approximately one-third of people with epilepsy do not achieve sustained seizure freedom with medication alone, and the burden of ongoing seizures includes injury risk, impaired quality of life, psychosocial disability, and premature mortality.1-4 Additional low-cost adjunctive strategies are therefore of continuing interest, particularly for patients who are not candidates for resective surgery, neuromodulation, or established dietary therapies.

Dietary treatment can modify seizure burden in some patients. The ketogenic diet and related approaches have demonstrated efficacy in selected populations with drug-resistant epilepsy, supporting the broader premise that nutritional interventions may influence seizure susceptibility.5-7 Chicken soup has long been regarded as a traditional therapeutic food, and in vitro data suggest that it may have modest anti-inflammatory effects.8,9 Because inflammatory pathways have been implicated in epileptogenesis and seizure generation,10,11 it is biologically plausible, at least in principle, that a food-based intervention could affect seizure frequency.

The original study was conceived as a pilot trial rather than a definitive efficacy study. To present it accurately, it is important to state that the design was a parallel-group randomized controlled experiment with a usual-diet control, not a blinded placebo-controlled trial. We therefore report the study as such and focus on the magnitude, direction, and limitations of the observed effects rather than on unsupported efficacy claims.

### Methods

Study design and oversight. This was a single-center, parallel-group randomized controlled pilot trial conducted at the Epilepsy Research Unit, XXX. The protocol was approved by the XXX Institutional Review Board (UB-IRB-XXX), and all participants provided written informed consent. The trial was conducted in accordance with the Declaration of Helsinki and International Council for Harmonisation Good Clinical Practice principles.

Participants. Eligible participants were aged 16 years or older and had drug-resistant focal epilepsy, defined according to International League Against Epilepsy criteria as failure of adequate trials of two tolerated, appropriately chosen ASM regimens to achieve sustained seizure freedom.2 Participants were required to have at least four focal seizures per month during the 3-month screening period before enrollment. ASM regimens were to remain stable during baseline and treatment whenever clinically feasible.

Study periods. The protocol comprised 3 distinct phases: a 3-month screening period used to confirm eligibility, a 2-month prospective baseline period used to establish pretreatment seizure frequency, and a 3-month treatment period. Seizures were recorded in standardized diaries throughout baseline and treatment.

Randomization and intervention. Participants were randomized in a 1:1 ratio to one of two parallel groups: chicken soup once daily (test group) or continuation of the usual diet without chicken soup supplementation (control group). Randomization was generated by computer in permuted blocks of four and stratified by baseline seizure frequency. Because the control condition was usual diet, the trial was not blinded and did not include a placebo intervention.

Outcomes. The primary outcome was percentage change in monthly seizure rate from the 2-month baseline period to the 3-month treatment period, calculated as 100 x (baseline rate - treatment rate) / baseline rate, such that positive values indicate improvement. The secondary outcome was the 50% responder rate, defined as the proportion of participants with a 50% or greater reduction in monthly seizure rate. For participants who discontinued early, last observation carried forward (LOCF) was used, consistent with the original analytic plan. Percentage change was undefined when the baseline seizure rate was zero.

Statistical analysis. Between-group comparisons used the Mann-Whitney U test for percentage change and Fisher exact test for responder rates. Analyses were performed in Python 3.11 with SciPy 1.11. All tests were 2-sided, and p < 0.05 was considered statistically significant. Given the pilot nature of the study and the small sample, results are reported primarily as descriptive estimates with associated p values.

### Results

Thirty participants were randomized, with 15 assigned to chicken soup and 15 assigned to control. During treatment, 7 of 15 participants (46.7%) in the chicken soup group and 2 of 15 (13.3%) in the control group discontinued before completing follow-up. LOCF was applied for these withdrawals. One control participant had a baseline seizure rate of zero during the 2-month baseline period; because percentage change is undefined in that setting, that participant was excluded from the percentage-change analysis but remained part of the responder analysis as originally tabulated.

For the primary outcome, the median percentage change in monthly seizure rate was 7.69% in the chicken soup group and -39.47% in the control group (Mann-Whitney U = 60.0, p = 0.1389). Numerically, the distribution favored chicken soup, but the between-group difference did not meet the prespecified threshold for statistical significance.

For the secondary outcome, the 50% responder rate was 15.4% (2/13) in the chicken soup group and 14.3% (2/14) in the control group (Fisher exact p = 1.0). Thus, the trial did not demonstrate a difference in conventional responder status between groups.

### Discussion

This pilot randomized controlled trial did not provide statistically significant evidence that daily chicken soup reduces seizure frequency in adults with drug-resistant focal epilepsy. The primary analysis showed a numerical between-group difference favoring chicken soup, but the estimate was imprecise and should not be overinterpreted. The secondary responder analysis was essentially neutral.

Several design and conduct features limit inference. First, the study was small and therefore underpowered to detect anything other than a large effect. Second, attrition was substantial and unbalanced, with more discontinuations in the chicken soup group than in the control group. This creates vulnerability to attrition bias and makes any LOCF-based estimate especially fragile.13 Third, because the control group continued usual diet rather than receiving a matched placebo, the study was open-label. Expectation effects, behavior changes, and differential reporting cannot be excluded. Finally, a food-based intervention may require stronger adherence assessment and longer observation than were available here.

The appropriate conclusion is therefore conservative: these data do not establish efficacy. At most, they justify further study only if a future trial uses a more rigorous design, including clear allocation concealment, a matched control condition, prespecified adherence monitoring, and an analysis strategy less sensitive to informative dropout. Any subsequent study should also be adequately powered and should report participant flow and missing-data handling in full.

In summary, daily chicken soup was not shown to improve seizures in this pilot study of drug-resistant focal epilepsy. The observed numerical trend in the primary analysis is hypothesis-generating only and is insufficient to support clinical use.

### Table 1. Core design features and principal results

| Item | Details |
| --- | --- |
| Design | Single-center, parallel-group randomized controlled pilot trial |
| Screening period | 3 months before enrollment to confirm eligibility |
| Baseline period | 2 months |
| Treatment period | 3 months |
| Intervention | Chicken soup once daily |
| Control | Usual diet |
| Blinding | Not blinded; no placebo control |
| Randomized sample | 30 participants (15 per group) |
| Primary outcome | Percentage change in monthly seizure rate from baseline to treatment |
| Primary result | Median percentage change: 7.69% (chicken soup) vs -39.47% (control); Mann-Whitney U = 60.0, p = 0.1389 |
| Secondary result | 50% responder rate: 15.4% (2/13) vs 14.3% (2/14); Fisher exact p = 1.0 |
| Key limitation | Substantial differential attrition: 46.7% in chicken soup vs 13.3% in control |

### Ethical Publication Statement

The authors confirm that this manuscript is consistent with the journal's guidance on ethical publication.

### Disclosure of Conflicts of Interest

The authors declare no conflicts of interest relevant to this work.

### Data Availability Statement

Deidentified study data are available from the corresponding author on reasonable request.

### Funding

This study was funded by the XXX Departmental Research Fund. The funder had no role in study design, data collection, analysis, interpretation, or manuscript preparation.

### Author Contributions

XXX conceived and designed the study, enrolled participants, analyzed the data, and drafted the manuscript. XXX contributed to study design, participant enrollment, and data collection. XXX supervised the study, contributed to interpretation, and critically revised the manuscript. All authors approved the final version.

2.2 **Reviews**

The full text of each AI-generated review is below. Reviews were compiled for both AI-generated and human-generated papers.

*Reviews generated by Epileptologist A*

Review for AI-generated paper supplied by Epileptologist B

**Overall assessment**

This manuscript reports a small, parallel‑group randomized controlled pilot trial evaluating daily chicken soup as an adjunctive intervention in adults with drug‑resistant focal epilepsy. The authors are appropriately conservative in their interpretation and are generally transparent about the study’s limitations, particularly the small sample size and substantial attrition. I appreciate the explicit framing of the study as hypothesis‑generating rather than efficacy‑establishing.

That said, there are several important design and analytic issues—most notably differential dropout, use of last observation carried forward (LOCF), and a problematic primary outcome definition—that substantially limit interpretability. While the negative result itself is not an issue, the manuscript would benefit from clearer acknowledgment of how fragile the estimates are, more cautious language regarding the direction of effects, and some restructuring for clarity. I think this manuscript could be publishable as a pilot/null study after revision, but only with clearer framing and some methodological tightening.

**Major comments**

1. **Differential attrition and risk of bias**

Attrition was high and markedly imbalanced (46.7% in the chicken soup group vs 13.3% in controls). This is the dominant methodological issue in the study and threatens internal validity more than any other limitation.

While LOCF was prespecified, LOCF is especially problematic in the setting of differential and potentially informative dropout. Participants discontinuing chicken soup may systematically differ in seizure trajectory or tolerability. The manuscript should more strongly emphasize that any between‑group comparison—directional or otherwise—is highly vulnerable to attrition bias.

At minimum, I would encourage:

- Explicit reporting of **reasons for withdrawal**, by group
- A **complete‑case analysis** presented alongside the LOCF analysis (even if underpowered), explicitly labeled as exploratory
- Even stronger language in the discussion noting that LOCF may bias results in unpredictable directions

2. **Primary outcome definition and interpretability**

The primary outcome—percentage change in seizure rate—is standard but fragile, particularly in small samples and when baseline seizure counts are low. Excluding a randomized control participant because baseline seizures were zero further complicates interpretation and breaks randomization for the primary analysis.

The authors should:

- Clearly state in the abstract and results that the primary analysis excluded a randomized participant
- Acknowledge more explicitly that percentage change magnifies noise at low seizure counts and is sensitive to outliers
- Consider reporting **absolute change in seizures/month** as a supplementary or descriptive outcome to provide clinical context

3. **Direction of effect language**

In multiple places, the manuscript states that the “between‑group distribution favored chicken soup numerically.” However, the reported medians show improvement in the chicken soup group (7.7%) and worsening in controls (−39.5%), but with extremely wide implicit variability and heavy influence from LOCF.

I would recommend tempering this language. “Favored” implies a meaningful signal, which is not clearly supported given:

- Small n
- Attrition imbalance
- LOCF use
- Exclusion of a randomized participant

More neutral phrasing (e.g., “the point estimates differed between groups but were imprecise and not statistically distinguishable”) would be more accurate and consistent with the study’s limitations.

4. **Open‑label design and expectation effects**

The lack of blinding and absence of a placebo control is appropriately described, but the implications deserve more emphasis. Dietary interventions are particularly susceptible to expectation, adherence, and reporting effects, especially when seizure diaries are self‑reported.

The authors should explicitly acknowledge that:

- Seizure reporting may differ by group
- Behavioral changes unrelated to soup consumption (e.g., routine changes, attention effects) cannot be separated from the intervention

5. **Biological plausibility discussion**

The inflammation‑based rationale is reasonable but relatively speculative. Given a negative pilot result, I would suggest tightening this section and avoiding overemphasis on plausibility, which risks distracting from the methodological message.

The strongest contribution of this paper is not biological insight but **methodological transparency regarding a low‑signal intervention**, and the discussion could lean more explicitly into that.

**Minor comments**

- The abstract is generally clear, but best practice would be to include **attrition numbers** explicitly in the Results section of the abstract, given their importance.
- Please clarify whether adherence to daily chicken soup consumption was assessed objectively or only inferred from participation.
- “Favored numerically” appears multiple times; consider varying or softening this phrasing.
- The exclusion of focal seizure subtype breakdown (focal aware vs focal impaired awareness) should be acknowledged, even briefly.
- Table 1 is clear and appropriate for a Brief Communication; however, adding a participant flow diagram (even as supplementary material) would improve transparency.
- The statistical software description is sufficiently detailed; no changes needed.

**Summary recommendation**

This is a small, carefully described pilot randomized trial with a negative result. Its value lies primarily in illustrating the challenges of dietary and behavioral interventions in drug‑resistant epilepsy, particularly attrition and outcome instability. With revisions to strengthen framing, temper conclusions, and improve clarity around bias and missing data, the manuscript could be suitable for publication as a hypothesis‑generating pilot study.

Review for human-generated paper supplied by Epileptologist C

**Overall assessment**

This is a timely and clinically interesting case report describing early seizure freedom and concordant metabolic response on F‑DOPA PET following treatment with vorasidenib in a patient with IDH‑mutant grade 2 oligodendroglioma. The manuscript is clearly written, the imaging is high quality, and the clinical narrative is coherent. The alignment of early seizure control with a PET response preceding MRI changes is particularly compelling and directly relevant to current discussions about seizure control as an outcome measure in low‑grade glioma trials.

That said, this remains a single case, and several interpretations—particularly regarding causality, seizure mechanisms, and PET signal specificity—are necessarily speculative. I think the manuscript is suitable for publication as a Brief Communication or case report after revision, but it would benefit from more cautious framing, clearer separation of observation from inference, and tightening of several methodological and interpretive points.

**Major comments**

1. **Causality and attribution of seizure freedom**

The most clinically attractive aspect of this report is the early seizure freedom following initiation of vorasidenib without ASM changes. However, in a single case, it is difficult to definitively attribute seizure control to IDH inhibition rather than to postoperative evolution, regression to the mean, or delayed effects of surgery.

While the authors largely acknowledge this implicitly, I would encourage more explicit caution in the abstract and discussion when linking seizure freedom directly to vorasidenib. Language suggesting association rather than causation would reduce the risk of overinterpretation.

2. **Seizure outcome characterization**

Seizure frequency is described as approximately two seizures per week prior to treatment, with seizure freedom achieved after one month. It would strengthen the report to:

- Clarify the duration of seizure freedom more explicitly (e.g., continuous months seizure‑free)
- Specify whether auras were completely absent, as per the ILAE seizure outcome definition referenced
- Indicate whether seizure diaries were corroborated by caregiver report or only self‑report

Given the emphasis placed on seizures as a biomarker, precision here is important.

3. **Role and specificity of F‑DOPA PET changes**

The PET findings are compelling, particularly the reduction in metabolic tumor volume preceding clear volumetric MRI change. However, F‑DOPA uptake may be influenced by seizure activity itself, as the authors acknowledge later in the discussion.

I would suggest:

- More explicit acknowledgment upfront that reduced PET uptake could reflect reduced epileptic activity rather than (or in addition to) tumor metabolic response
- Clear separation in the discussion between PET as a tumor biomarker and PET as a potential marker of epileptogenic activity

This distinction is especially important given the speculative but intriguing suggestion that PET may “capture seizure activity.”

4. **Generalisability and uniqueness of the case**

The manuscript states that this is the first report of early seizure freedom with vorasidenib in this population. While that is likely accurate, it would be helpful to briefly contextualize why seizure outcomes were not reported in INDIGO and related trials and to emphasize that patients with uncontrolled seizures were excluded.

This would reinforce why case‑level evidence, while limited, is still valuable.

5. **Imaging methodology clarity**

The PET methodology is generally well described, but for readers less familiar with PET RANO 1.0:

- Clarify whether MTV segmentation was performed by a single rater or reviewed by more than one observer
- State whether MTV measurements were blinded to timepoint, even retrospectively

These details help readers assess reproducibility, even in a case report.

**Minor comments**

- The seizure classification reference should specify the exact year/version of the “updated” ILAE classification rather than using placeholders.
- Percent or absolute change in MTV could be reported explicitly in addition to raw volumes for ease of interpretation.
- Consider briefly stating why ASM escalation was deferred prior to vorasidenib initiation, as this may differ across practices.
- The discussion of 2‑HG and mTOR signaling is interesting but could be slightly condensed to maintain focus on the clinical observation.
- Figures are high quality; minor improvement could be made by marking the lesion consistently across all panels for readability.

**Summary recommendation**

This is a well‑documented and clinically relevant case report that highlights a potentially important effect of IDH inhibition on seizure control and demonstrates the complementary value of amino acid PET imaging. With more cautious causal language, clearer seizure outcome description, and tighter framing of PET interpretation, the manuscript would make a valuable contribution to the emerging literature on seizure outcomes in IDH‑mutant glioma treated with targeted therapy.

Review for AI-generated paper supplied by Epileptologist B

**Overall assessment**

This is a clinically appealing and pragmatic manuscript evaluating a nurse-led behavioral intervention targeting two well-established seizure precipitants—sleep deprivation and alcohol use. The focus on narrowly defined, modifiable behaviors is a strength, as is the feasibility and scalability of the intervention. The magnitude of seizure reduction reported is clinically meaningful and compares favorably with prior behavioral/self-management studies.

However, there are several important design, analytic, and interpretive issues that substantially limit causal inference in the current form. In particular, the nonrandomized design, absence of an attention-matched control, reliance on self-reported seizure frequency, and limited characterization of behavioral change raise concerns that the observed effect may partially reflect nonspecific effects of increased clinical contact and motivation rather than trigger-specific modification. I believe this work is promising and likely publishable after revision, but it would benefit from clearer framing, additional analyses, and more explicit acknowledgment of alternative explanations for the observed effects.

**Major comments**

1. **Attribution of seizure reduction to trigger modification**

The central claim of the manuscript is that targeted reduction of sleep deprivation and alcohol use led to improved seizure outcomes. However, it is difficult to disentangle the effect of trigger modification from nonspecific effects of increased clinical attention, motivational support, and weekly contact. The control group received routine advice without structured follow-up, which makes this closer to an “attention versus no attention” comparison rather than a trigger-specific intervention comparison.

While the authors acknowledge this in the limitations, it would be important to more explicitly frame the findings as demonstrating benefit of a *structured counseling program*, rather than definitive evidence that modifying these specific triggers caused seizure reduction. A clearer distinction between “STOP IT versus standard care” and “trigger reduction versus no trigger reduction” would strengthen the interpretation.

2. **Nonrandomized design and selection bias**

Patients were not randomized, and assignment depended on timing and provider participation. This raises substantial concerns about selection bias and unmeasured confounding (e.g., motivation, insight, adherence, health literacy). While baseline demographic and seizure characteristics appear balanced, motivation to change behavior is not measurable here and likely differs between groups.

The multivariable regression adjusting for baseline covariates does not meaningfully address this source of bias. I would strongly recommend either (a) more cautious causal language throughout, or (b) additional sensitivity analyses (e.g., propensity score methods, even if limited) to demonstrate robustness.

3. **Outcome definition and seizure frequency ascertainment**

Seizure frequency is derived primarily from self-report and clinic documentation. Given that the intervention involves frequent contact with nursing staff, there is a real possibility of differential reporting, recall, or diary completion between groups. This is particularly important when the primary outcome is percent change rather than absolute counts.

Please expand the discussion of this measurement bias and clarify whether seizure diaries were systematically collected in *both* groups with similar frequency and structure. If available, describing visit frequency or documentation density between groups would be helpful.

4. **Handling of missing data and attrition**

Patients lost to follow-up were excluded from the primary analysis, with a sensitivity analysis using last observation carried forward (LOCF). Excluding patients with missing outcome data risks bias, especially in a small, nonrandomized study.

Please report the sensitivity analysis results explicitly (not just state that it was performed), and discuss limitations of LOCF in this context. Even a brief justification for excluding adherence or seizure frequency from imputation would help readers assess robustness.

5. **Statistical model choice for responder analysis**

The use of logistic regression for the 50% responder rate is reasonable, but with only 27 total responders across both groups, the multivariable model may be overfit. Please clarify how many covariates were included relative to the number of outcome events, and consider a more parsimonious model or presentation of unadjusted and minimally adjusted analyses.

6. **Behavioral adherence outcomes**

Behavioral change is reported descriptively (e.g., adherence to target bedtime, reduction in alcohol use), but definitions of these measures are vague. What constitutes “consistent adherence” or “meaningful reduction”? Were these assessed using any standardized instrument, or were they purely subjective?

Since behavioral modification is the proposed mechanism, clearer operationalization is important. Even acknowledging that these are coarse, self-reported measures would improve transparency.

7. **Generalisability and epilepsy subtypes**

The cohort includes both focal and generalized epilepsy, yet alcohol and sleep deprivation may have differential effects across syndromes (particularly GGE). Consider either subgroup analyses (if underpowered, framed as exploratory) or discussion of whether the observed effect plausibly applies equally across epilepsy types.

**Minor comments**

- In the abstract, best practice is to present numerical results first, followed by interpretation. The abstract is generally strong but could adhere more closely to that structure.
- The term “triggers” may be interpreted causally; consider occasional use of “self-reported seizure precipitants” to maintain consistency with the methodology.
- Please specify the dates of enrollment in the Methods section (currently listed as XXX–XXX).
- The TREND adherence statement is appreciated; however, a brief note on how this informed reporting (e.g., flow, baseline comparability) would strengthen transparency.
- Percentages for small numerators (e.g., adherence outcomes) should be interpreted cautiously; consider also reporting raw counts.
- Figure 1 is clear, but adding individual data points (e.g., jittered dots) or a brief caption noting whether percent change was truncated or winsorized would improve interpretability.
- In Table 1, consider reporting epilepsy syndrome (where known) rather than only focal vs generalized, as trigger relevance may differ by syndrome.
- The discussion appropriately cites prior behavioral trials; it may be helpful to contrast STOP IT more explicitly with multi-component self-management programs in terms of intensity and resource utilization.

**Summary recommendation:** Promising and clinically relevant study that would benefit from revision to clarify causal claims, address bias inherent in the design, and strengthen reporting of behavioral and statistical methods.

Review for human-generated paper supplied by Epileptologist B

**Overall assessment**

This manuscript addresses an unusual but historically important phenomenon—the long-term outcomes of seizures induced during the “Pocket Monster” television broadcast—and attempts to differentiate epilepsy trajectories based on the presence or absence of pre-existing epilepsy. The 5-year follow-up is a notable strength, and the comparison between patients with and without prior epilepsy is clinically intuitive and potentially relevant for counseling patients after visually induced seizures.

That said, the study is limited by its small sample size, reliance on retrospective questionnaire data, heterogeneity in EEG acquisition and classification, and use of outdated seizure and epilepsy classification schemes. Several conclusions are plausible but are presented with greater certainty than the data support. I believe the manuscript could be suitable for publication after revision, but it would benefit from clearer framing as a descriptive, hypothesis-generating study, modernization of terminology, and more cautious interpretation of causal mechanisms—particularly regarding photosensitivity and epilepsy syndrome attribution.

**Major comments**

1. **Study design and selection bias**

The study population is restricted to the 25 of 91 patients who experienced seizure recurrence, which necessarily conditions all analyses on recurrence and limits inference about overall risk after a Pocket Monster seizure. This selection should be emphasized much more explicitly, including in the abstract and discussion.

In addition, all data are derived from physician questionnaires completed years after the initial event. This introduces substantial risk of recall bias and inconsistency in ascertainment (e.g., seizure types, EEG findings, timing of recurrence), which should be more clearly acknowledged.

2. **Small sample size and statistical interpretation**

The effective sample size (n = 25 total; 12 vs 13 per group) is extremely limited, yet multiple subgroup comparisons are performed. Several statistically significant findings (e.g., seizure type, syndrome classification) are likely unstable, and many nonsignificant trends are discussed as meaningful.

I would strongly encourage the authors to de-emphasize p-values and reframe these comparisons descriptively, highlighting effect sizes and clinical patterns rather than statistical significance.

3. **Heterogeneity and nonstandardized EEG assessment**

The definition of photoparoxysmal response (PPR) is standard, but the protocol for intermittent photic stimulation explicitly varied among hospitals. This severely limits comparability of PPR rates between groups and over time.

Given that photosensitivity is a central construct in the manuscript, this limitation should be discussed more prominently, and conclusions regarding the relative role of photosensitivity should be softened accordingly.

4. **Use of outdated classification systems**

Seizure types and epilepsy syndromes are classified using the 1981 and 1989 ILAE frameworks. While understandable given the study timeline, this introduces ambiguity when interpreting “localization-related epilepsy,” “idiopathic epilepsy,” and related categories.

At a minimum, the authors should acknowledge this limitation and, where possible, clarify how these categories would approximately map onto contemporary ILAE terminology, particularly for generalized epilepsies and frontal lobe epilepsy.

5. **Causal interpretation of “triggering” versus disease manifestation**

The discussion implies that the Pocket Monster broadcast may have unmasked latent idiopathic generalized epilepsies in patients without prior epilepsy, whereas it occurred during periods of heightened intrinsic epileptogenicity in those with prior epilepsy. While plausible, this interpretation goes beyond what the data can directly support.

An alternative explanation—that the PM seizure represented an early manifestation of generalized epilepsy in some patients regardless of photosensitivity—should be discussed more explicitly. The distinction between a true “triggered seizure” and the first clinically recognized seizure of epilepsy deserves more careful handling.

6. **Treatment decisions and outcome interpretation**

Antiepileptic drug use differed markedly between groups, yet treatment decisions are not analyzed as potential contributors to seizure recurrence timing, frequency, or long-term outcomes. Given that AED exposure was nearly universal in the epilepsy group but uncommon initially in the nonepilepsy group, treatment effects may confound observed differences.

The authors should either explore this impact more directly or acknowledge it as a major confounder.

**Minor comments**

- The abstract would benefit from explicitly stating that only patients with seizure recurrence were included in the analysis.
- The term “mental retardation” should be replaced with “intellectual disability,” in line with current terminology.
- Percentages are reported for very small denominators; adding raw numerators consistently would improve clarity.
- The variability and missingness of EEG data (e.g., 4/12) should be explained more clearly in the Methods.
- In Table 1, the epilepsy syndrome row is dense and difficult to interpret; consider simplifying or splitting into generalized vs focal categories only.
- The discussion of frontal lobe epilepsy and photosensitivity is interesting but speculative; this should be clearly labeled as such.
- Please clarify whether any patients had visually induced seizures outside the Pocket Monster incident prior to that broadcast.

**Summary recommendation**

This is a niche but historically and clinically interesting dataset that adds descriptive information about long-term outcomes after visually induced seizures. With clearer acknowledgment of selection bias, modernization of terminology, and more cautious interpretation of photosensitivity and causality, the manuscript could make a useful contribution as a descriptive follow-up study.

*Reviews generated by Epileptologist B*

Review for AI-generated paper supplied by Epileptologist A

**Summary**

The manuscript reports a single-center cross-sectional survey of 75 epilepsy monitoring unit clinicians comparing responses to matched clinical vignettes of functional seizures and epilepsy using an Attribution Questionnaire-based stigma measure. It reports higher responsibility, anger, coercion, and avoidance scores for functional seizures, and lower pity, fear, and dangerousness scores compared with epilepsy. It also reports associations between higher functional-seizure stigma scores and professional role, years of experience, fellowship training, and prior education.

**Overall readiness for Epilepsia**

Potentially suitable after major revision

The topic is relevant to Epilepsia, but the current manuscript has material problems in measurement validity, vignette comparability, statistical modeling, and interpretation. Several central conclusions are not yet adequately supported by the reported methods and internally inconsistent results.

**Major concerns**

1. 1.

- Issue: The stigma instrument and scoring are internally inconsistent. Section 2.2 describes the Attribution Questionnaire as a validated instrument, states that each item used a 5-point Likert scale, and Table 2 labels the measure as “AQ-27” while reporting domain ranges of 1–9 and a total composite range of 9–81. These ranges are more consistent with a 9-item short form than with the AQ-27. The AQ-27 is described in the literature as nine subscales with three questions per subscale, and items are presented on a 9-point Likert scale; the AQ-9 is the 9-item short form derived from the AQ-27.
- Why it matters: The primary outcome is unclear. If the authors used AQ-9 items, AQ-27 subscale reliability claims and “domain” language are not appropriate as written. If the authors used AQ-27 items, the reported scale ranges, means, and composite score appear incorrectly scored. This affects every quantitative result.
- Requested revision or analysis: State the exact instrument version used, provide the number of items, item response scale, item wording or appendix, scoring rules, reverse coding, domain construction, and composite construction. Recalculate all domain and total scores if needed. If an adapted or abbreviated instrument was used, state that explicitly, justify the adaptation for functional seizures and epilepsy vignettes, and avoid citing AQ-27 subscale psychometrics as if they directly apply.

2. 2.

- Issue: The two vignettes are not sufficiently matched to isolate diagnostic-label stigma. They differ in duration, aura description, semiology, eye state, amnesia, postictal symptoms, emotional framing, and ictal EEG findings. Several differences plausibly influence ratings of dangerousness, fear, pity, responsibility, and clinician willingness to intervene independent of the diagnostic label.
- Why it matters: The central comparison is framed as stigma toward functional seizures versus epilepsy, but the observed differences may reflect the specific clinical details of the vignettes rather than attitudes toward the diagnoses. For example, lower dangerousness and fear scores for functional seizures may be an expected response to the epilepsy vignette’s generalized convulsive features rather than evidence about stigma.
- Requested revision or analysis: Describe how the vignettes were developed, whether they were balanced for severity, impairment, chronicity, treatment context, and perceived controllability, and what feedback was obtained during pilot testing. Report any order-effect analyses. Revise conclusions to specify that the findings reflect responses to these vignettes and diagnostic framing, not necessarily global clinician attitudes toward all patients with functional seizures versus epilepsy.

3. 3.

- Issue: The multivariable regression results contain serious internal inconsistencies. Table 1 states that 75/75 respondents had prior education on functional seizures, but Table 3 reports a statistically significant regression coefficient for prior education. Table 3 also reports “epilepsy fellowship training” as “physicians only” while simultaneously stating that the model was adjusted for all variables shown across the full sample. The coding of nonphysicians for fellowship status is unclear and may be confounded with professional role. The row labeled “5 years” under years of experience appears to mean “<5 years.”
- Why it matters: The manuscript’s key claims that nurses, EEG technologists, less experienced clinicians, fellowship training, and prior education are associated with stigma may be artifacts of coding, collinearity, or model misspecification. The prior education coefficient is not estimable if all respondents had the same value.
- Requested revision or analysis: Correct the dataset description and regression table. Remove predictors without variation. Specify the analytic sample for each model, coding of categorical variables, reference groups, and handling of physician-only variables. Consider presenting fellowship training only in a physician-only exploratory model. Provide a parsimonious model appropriate for n=75, and clearly label these analyses as exploratory unless they were prespecified.

4. 4.

- Issue: The paired comparisons and composite score need stronger statistical justification. The manuscript compares nine AQ domains plus a total composite without a stated primary outcome or adjustment for multiple comparisons. Cohen’s d is reported without specifying whether it is the paired-samples effect size. Confidence intervals are provided for mean differences but not for effect sizes. The total “composite stigma score” is also conceptually unclear because some domains, such as pity and help, do not have the same valence as anger, avoidance, coercion, responsibility, fear, dangerousness, and segregation.
- Why it matters: Without a prespecified primary endpoint, multiplicity handling, and a justified composite, the statement that functional seizures elicit “higher stigma” may be statistically and conceptually overstated. A higher or lower score on individual AQ constructs does not automatically translate into a valid global stigma score unless the scoring direction and construct model are defined.
- Requested revision or analysis: Define the primary outcome. State whether p-values are adjusted, and if not, justify treating domain analyses as exploratory. Specify the paired effect-size formula and report effect-size confidence intervals. Justify the composite score, including whether any domains were reverse-coded, or remove/de-emphasize the composite and interpret domains separately.

5. 5.

- Issue: The qualitative methods are insufficiently described for the inferences drawn. Section 2.3 states that qualitative comments were “analyzed thematically,” but the manuscript does not report the coding approach, number and training of coders, whether coding was independent, how disagreements were resolved, whether a codebook was used, or whether themes were inductive or deductive.
- Why it matters: The Discussion uses the qualitative findings to infer that stigma may stem from therapeutic helplessness and management uncertainty. That interpretation may be plausible, but the current reporting does not allow readers to assess analytic rigor or reproducibility.
- Requested revision or analysis: Provide a concise qualitative methods subsection. Report denominators for each theme, clarify whether comments could receive multiple codes, and indicate how representative the quotations are. Temper mechanistic language unless supported by a transparent analytic process.

6. 6.

- Issue: Several conclusions overstate what can be inferred from a single-center, cross-sectional, self-report vignette survey. The abstract and Discussion state or imply that the findings “validate patient-reported experiences,” identify “modifiable targets,” show fellowship training is “protective,” and support educational interventions targeting nurses and early-career clinicians.
- Why it matters: The study measures clinician responses to hypothetical vignettes at one academic EMU. It does not measure clinician behavior, patient experiences, educational intervention effects, causality, or longitudinal change.
- Requested revision or analysis: Reframe these points as hypotheses or implications rather than demonstrated effects. Use “associated with lower stigma scores” rather than “protective.” Replace “validate patient-reported experiences” with language indicating that the findings are consistent with patient-reported healthcare stigma. Limit intervention claims to the need to test targeted education.

7. 7.

- Issue: Nonresponse bias is mentioned in Methods but not reported in Results. The manuscript states that nonresponse bias was assessed by comparing clinician characteristics between responders and nonresponders, yet no responder-versus-nonresponder table or narrative result is provided.
- Why it matters: The study’s response rate is high, but respondents may differ systematically from nonrespondents in role, training, workload, or attitudes toward functional seizures. This is especially important because the study examines potentially sensitive stigmatizing attitudes.
- Requested revision or analysis: Report the available characteristics of the 25 nonrespondents compared with the 75 respondents, including which variables were available for all eligible clinicians. If such data were not available, remove the claim that nonresponse bias was assessed and add this as a limitation.

**Minor concerns**

1. 1.

- Issue: The manuscript contains unresolved placeholders: “XXX Institutional Review Board,” “STUDYXXX,” and recruitment dates “between XXX and XXX.”
- Why it matters: These details are needed to assess ethics approval, timing, and recruitment context unless they are intentionally blinded for peer review.
- Requested revision: Replace placeholders before final submission or state clearly in the blinded version that these items are masked for review and will be restored.

2. 2.

- Issue: Funding, author disclosures, and conflicts of interest are not included in the submitted material.
- Why it matters: These statements are required for interpretation of clinician-survey research, especially when recruitment involved departmental leadership and compensation.
- Requested revision: Add funding, disclosures, and conflict-of-interest statements. Also clarify how voluntariness was protected when recruitment had leadership support.

3. 3.

- Issue: The data availability statement is not final: “e.g., Mendeley Data” is a placeholder and refers only to analysis code, not the survey dataset.
- Why it matters: Readers need to know what will be shared, where it will be deposited, and whether de-identified participant-level data are available.
- Requested revision: Provide a final repository, accession or DOI if available, and specify whether de-identified survey data, code, vignettes, and scoring scripts will be shared.

4. 4.

- Issue: The recruitment description states that up to 10 monthly reminder emails were sent, but the recruitment interval is masked.
- Why it matters: A long reminder period may affect response dynamics and could increase perceived pressure among employees.
- Requested revision: Report the recruitment dates or duration and clarify whether reminders stopped after completion and whether study staff, rather than supervisors, managed reminders.

5. 5.

- Issue: P-value formatting is inconsistent. The Results text uses “p = < 0.001,” while Table 2 lists several values as “0.001.”
- Why it matters: This creates ambiguity about the actual test results.
- Requested revision: Report exact p-values where appropriate or use a consistent threshold format such as “p < .001.”

6. 6.

- Issue: Table 1 reports “Epilepsy fellowship-trained 11 (15%)” in the total sample, while the text states that 61% of physician respondents had fellowship training.
- Why it matters: Both statements may be mathematically correct, but the table presentation can be misread as 15% of physicians rather than 11/18 physicians.
- Requested revision: Present fellowship training within the physician subgroup as 11/18 (61%) and, if desired, separately note that this equals 15% of the full sample.

7. 7.

- Issue: Table 3 omits the “Other” professional category or does not explain how it was handled.
- Why it matters: Excluding or collapsing small groups can affect role comparisons in a small sample.
- Requested revision: State whether “Other” clinicians were excluded, combined with EEG technologists, or included in another category.

8. 8.

- Issue: The phrase “No significant associations were found between stigma scores and clinical characteristics” is unclear and appears inconsistent with reported associations for professional role, experience, fellowship training, and prior education.
- Why it matters: “Clinical characteristics” is undefined and may confuse readers.
- Requested revision: Define the term or remove the sentence.

9. 9.

- Issue: The Introduction defines functional seizures as occurring “without aberrant brain electrical activity.”
- Why it matters: This wording is imprecise; the relevant distinction is absence of epileptic ictal activity rather than absence of abnormal brain activity in a broad neurobiological sense.
- Requested revision: Revise to a more precise formulation, such as events resembling epileptic seizures but not caused by epileptic neuronal discharges and without ictal epileptiform EEG correlates during captured typical events.

10. 10.

- Issue: The novelty claim “first systematic comparison” is not adequately supported. The manuscript itself cites prior provider-attitude and healthcare-professional literature, but does not clearly explain how the current study differs from or extends those studies.
- Why it matters: Epilepsia readers will expect precise contextualization of originality.
- Requested revision: Qualify the claim or add a concise explanation of the specific novelty, such as direct within-respondent comparison of functional seizure and epilepsy vignettes among EMU clinicians.

11. 11.

- Issue: Several citations appear misaligned with the claims they support. For example, attribution theory in the Discussion should be tied to the Weiner attribution reference rather than the longitudinal FND stigma study; the statement about a web-based educational module appears to cite Ref. 9 rather than Ref. 17; and the sentence on cognitive behavioral therapy and multidisciplinary treatment cites references that, by title, appear to address education rather than treatment efficacy.
- Why it matters: Mis-citation weakens the Discussion and may mislead readers about the evidence base.
- Requested revision: Audit all Discussion citations against the claims they support, especially Refs. 8, 9, 13, 14, 17, and 18.

12. 12.

- Issue: The manuscript claims adherence to STROBE but omits several relevant reporting elements, including missing-data handling, survey completion definition, participant flow beyond completed surveys, and nonresponder comparison results.
- Why it matters: These omissions reduce reproducibility and transparency for an observational survey.
- Requested revision: Add the missing reporting elements and include a completed STROBE checklist if required by the journal.

Review for human-generated paper supplied by Epileptologist A

**Summary**

The manuscript reports a retrospective evaluation of Re-PROGRAM, a five-session telehealth skills-based intervention for patients with functional seizures in an outpatient hospital clinic. Twenty-nine enrolled patients completed scheduled sessions; postintervention feedback was obtained via a program-specific retrospective comparison survey in 16 patients, with chart review used for some survey nonresponders. The authors conclude that Re-PROGRAM is feasible and acceptable and warrants controlled evaluation.

**Overall readiness for Epilepsia**

Potentially suitable after major revision

The topic is relevant to Epilepsia, but the current manuscript overinterprets an uncontrolled, retrospective service evaluation with incomplete follow-up, nonstandardized outcomes, and an unvalidated post hoc survey instrument. Substantial revision is needed to confine claims to feasibility/acceptability, report selection and missing data transparently, and add ethics, safety, disclosure, and reproducibility details.

**Major concerns**

1. 1.

- Issue: The manuscript repeatedly implies efficacy despite a design that cannot support causal inference. The abstract, key points, Results, and Discussion state or imply that the intervention “reduced seizure occurrence,” “improved daily functioning,” and produced seizure control in “most patients,” although there was no control group, no randomization, no prospective baseline seizure diary, no standardized seizure-frequency endpoint, and variable postintervention assessment timing.
- Why it matters: Improvement after diagnosis and treatment initiation may reflect regression to the mean, natural fluctuation, expectancy effects, diagnostic communication, concurrent clinical care, medication changes, or selective response among satisfied patients. Epilepsia readers may interpret the current wording as evidence of treatment effectiveness rather than a preliminary service evaluation.
- Requested revision or analysis: Reframe the manuscript throughout as an uncontrolled feasibility/acceptability evaluation. Remove or qualify efficacy language in the abstract, key points, Discussion, and Conclusions. State that seizure reductions were patient-reported or chart-documented after the intervention and cannot be attributed to Re-PROGRAM. Distinguish “completion of scheduled sessions among enrolled patients” from adherence, acceptability, or efficacy.

2. 2.

- Issue: Participant flow and selection are insufficiently described. The Methods report 29 enrolled patients, but do not provide the number assessed, eligible, offered the intervention, declined, excluded, or unable to commence. The criterion that patients were “cognitively and psychiatrically stable and free of acute risk issues” is vague, and the exclusion sentence appears to say that excluded characteristics “included” patients who were offered treatment. Baseline seizure frequency, baseline functional impairment, psychiatric comorbidity, other functional neurological symptoms, employment/school status, prior FS treatment, concurrent psychotherapy, psychotropic medication changes, and timing of antiseizure medication weaning are not reported.
- Why it matters: Feasibility and acceptability cannot be interpreted without the denominator of eligible and offered patients. The selected sample may represent a lower-risk, more motivated, or less complex subgroup, limiting generalizability. Concurrent treatments and medication changes may confound postintervention seizure reports.
- Requested revision or analysis: Add a STROBE-style flow diagram or equivalent text showing all patients screened, eligible, excluded, offered, enrolled, completed, surveyed, and followed clinically, with reasons where available. Define inclusion and exclusion criteria precisely. Add baseline clinical characteristics relevant to prognosis and response, including baseline FS frequency/burden and psychiatric/psychological treatment status. Clarify when antiseizure medications were weaned relative to Re-PROGRAM and whether any other treatment changes occurred during follow-up.

3. 3.

- Issue: Outcome ascertainment is vulnerable to major recall, response, and documentation bias. The FSPICS was designed for this evaluation, appears unvalidated, and asks respondents to make retrospective pre/post comparisons rather than using prospectively collected baseline and follow-up measures. Only 16 of 29 patients returned the survey, only 12 were complete, and the median time from intervention completion to survey completion was 229 days. For nonresponders, seizure trajectory was inferred from routine clinical notes, with four patients lost to follow-up and one case lacking documented baseline frequency.
- Why it matters: The survey responders may not represent the full cohort. A retrospective comparison completed months after treatment is not equivalent to a pre/post measurement. Chart review data are not comparable to structured survey data and may selectively capture patients who remained engaged in care.
- Requested revision or analysis: Report exact denominators for every outcome and every FSPICS item, including incomplete questionnaires. Present results separately for survey responders, nonresponders with clinical follow-up, and patients lost to follow-up. Provide a sensitivity analysis using all 29 enrolled patients as the denominator, including conservative assumptions that nonresponders/lost patients did not improve. Avoid combining survey and chart-review outcomes as if they were measured identically. Clarify whether outcomes refer specifically to functional seizures and how epileptic seizures were distinguished in the three patients with comorbid epilepsy.

4. 4.

- Issue: The statistical and qualitative analytic methods are underdeveloped. The Fisher exact test assessing seizure occurrence by time from onset to diagnosis is likely uninformative with n = 16, sparse cells, and an outcome in which nearly all respondents improved. No effect estimates or confidence intervals are reported. The qualitative analysis is described only as “thematic analysis” using NVivo, without specifying the coding framework, number of coders, independence from intervention delivery, reflexivity, adjudication, audit trail, or how negative/neutral feedback was handled.
- Why it matters: A single underpowered p-value can create false precision. The qualitative findings are central to the acceptability claim, but the analytic process is not reproducible and may be vulnerable to confirmation bias, especially if clinicians involved in treatment also collected or interpreted feedback.
- Requested revision or analysis: Replace inferential testing with descriptive statistics unless a defensible prespecified hypothesis and adequate contingency table are provided. Report exact counts and, where useful, exact binomial confidence intervals. For qualitative data, describe the analytic approach, coder roles, independence from treatment delivery, coding/reconciliation process, and counts of respondents contributing to each theme. Include negative and neutral feedback systematically. If these elements were not performed, present the qualitative material as descriptive patient feedback rather than formal thematic analysis.

5. 5.

- Issue: Intervention reproducibility and fidelity are not adequately documented. The manuscript states that a therapist manual and clinician script guide exist, but these were not available in the submitted materials. The manuscript refers to Supplementary Appendices S1–S3, Table S3, and supplementary references S1–S31, but these materials were not present in the reviewed file. Training, supervision, fidelity checks, protocol deviations, homework adherence, dose received, telehealth logistics, and management of distress during exposure/trigger work are not described.
- Why it matters: Readers cannot reproduce or evaluate the intervention, and the claim that provisional psychologists with little therapy experience can deliver it safely and consistently depends on training, supervision, and fidelity procedures. The absence of the survey instrument and supplemental intervention content prevents assessment of outcome validity and comparability between responders and nonresponders.
- Requested revision or analysis: Provide the missing supplementary materials or include sufficient intervention detail in the main manuscript. Report therapist training, supervision, fidelity monitoring, deviations, number and duration of sessions actually delivered, homework/practice adherence where available, and telehealth delivery procedures. Clarify whether the one-hour introductory interview makes this a six-contact intervention rather than only five sessions.

6. 6.

- Issue: Required ethics, consent, safety, funding, and disclosure information is absent. The manuscript does not state whether institutional review board/human research ethics approval was obtained, whether this was classified as research or service evaluation, whether consent or a waiver was obtained, how adolescent participation was handled given the age range includes 16 years, or how deidentified survey and chart-review data were governed. Funding and conflicts of interest are not reported. There is no systematic adverse-event or deterioration reporting, despite some survey items in Figure 2 appearing to include worse/much-worse responses for nonseizure outcomes.
- Why it matters: This is a human-participant intervention evaluation using clinical records and patient feedback. Ethics approval/waiver, consent procedures, safety monitoring, and disclosure are journal requirements and materially affect interpretability, especially when program developers and treating clinicians may have collected outcomes.
- Requested revision or analysis: Add a complete ethics statement, consent/waiver statement, data governance description, adolescent consent/assent procedures if applicable, funding statement, and conflict-of-interest disclosures. Disclose the authors’ role in developing, delivering, and evaluating Re-PROGRAM. Report adverse events, clinical deterioration, urgent referrals, symptom worsening, and safety procedures, or state explicitly that these were not systematically collected.

7. 7.

- Issue: The mechanistic framing is overstated relative to the data. The Introduction, Table 2, and Discussion describe FS as a “brain network miscommunication syndrome” and suggest that intervention components may “restore abnormal brain connectivity patterns” or target neuroinflammation, BDNF, autonomic regulation, default mode network activity, interoceptive circuits, and other mechanisms. None of these mechanisms were measured in this evaluation, and several rationales appear extrapolated from other populations rather than FS-specific intervention evidence.
- Why it matters: The manuscript risks implying mechanistic efficacy where only descriptive clinical feedback was collected. This may distract from the feasible clinical question and overstate the biological specificity of general wellness, behavioral activation, relaxation, and coping components.
- Requested revision or analysis: Recast mechanistic statements as hypotheses or theoretical rationale. Remove claims that Re-PROGRAM restores connectivity or directly targets specific neural mechanisms unless FS-specific evidence supports the claim and the intervention measured that mechanism. In Table 2, separate empirically supported FS-specific rationale from broader extrapolation from other fields.

**Minor concerns**

1. 1.

- Issue: The abstract and Methods contain unreplaced placeholders for study dates and setting (“XXX”).
- Why it matters: The study period and clinical setting are necessary to interpret recruitment, follow-up duration, service context, and generalizability.
- Requested revision: Provide the dates and setting where allowed by the review process, or ensure they are restored before publication.

2. 2.

- Issue: Table 1 contains an apparent data inconsistency for time from onset to diagnosis. The listed counts sum to 28 rather than 29, and the row “11–20 years, n = 2” is reported as 10.3%, which would correspond to 3 of 29 rather than 2 of 29.
- Why it matters: This raises concern about data accuracy and denominators.
- Requested revision: Correct the count, percentage, or add a missing-data row. Review all table percentages for consistency with denominators.

3. 3.

- Issue: The manuscript uses “seizure occurrence,” “seizure frequency,” “FS burden,” “seizure control,” “seizure bothersomeness,” “seizure intensity,” and “seizure duration” without clear operational definitions.
- Why it matters: These are not interchangeable outcomes, and imprecise terminology makes the results difficult to interpret.
- Requested revision: Define each outcome term, specify which were FSPICS items, and use terminology consistently across the abstract, Results, figures, and Discussion.

4. 4.

- Issue: Several statements refer to “all patients” when the relevant data apply only to survey respondents. For example, the Discussion states that all patients would recommend the program, but this was observed only among respondents who returned feedback.
- Why it matters: This inflates acceptability estimates by excluding nonresponders and patients lost to follow-up.
- Requested revision: Revise these statements to specify the denominator, for example “16/16 survey respondents,” and avoid extrapolating respondent feedback to the full cohort.

5. 5.

- Issue: Figure 2 presents percentages for a small and incompletely responding sample, and the denominator for each item is unclear. The legend states N = 16, although four questionnaires were incomplete.
- Why it matters: Percentages can exaggerate precision when the underlying counts are small and variable across items.
- Requested revision: Add raw counts and item-specific denominators to Figure 2 or an accompanying table. Indicate missing responses and clarify whether reverse-scored items were recoded before plotting.

6. 6.

- Issue: Figure 3 and Table 3 use inconsistent theme labels: Figure 3 lists “Intrapersonal development,” whereas Table 3 lists “Interpersonal development” for apparently similar content. Participant identifiers are also inconsistently formatted, for example P1 versus P03.
- Why it matters: Inconsistent labeling reduces clarity and makes it harder to map qualitative themes to illustrative quotations.
- Requested revision: Harmonize theme labels and participant identifier formatting across the text, figures, and tables.

7. 7.

- Issue: The Methods cite reference 34 for the documented diagnosis of FS, but that citation appears to be an editorial/commentary rather than the core diagnostic-standard reference. The submitted reference list also does not include the supplementary references supporting Table 2.
- Why it matters: Diagnostic ascertainment and mechanistic rationale should be supported by appropriate primary or consensus sources.
- Requested revision: Cite an appropriate diagnostic-standard source, such as the ILAE staged diagnostic certainty criteria for PNES/functional seizures, and provide the supplementary references supporting Table 2.

8. 8.

- Issue: The Discussion gives substantial space to the subsequently developed internet-based version and ongoing RCT, which are not evaluated in this retrospective cohort.
- Why it matters: This may shift the manuscript away from the submitted data and toward future program development.
- Requested revision: Shorten this section and retain only context necessary to explain why controlled evaluation is planned. Clearly separate findings from the present cohort from future or ongoing work.

Review for AI-generated paper supplied by Epileptologist C

Article 28:

Summary

The submitted manuscript reports a single-center, nonrandomized prospective controlled evaluation of the STOP IT nurse-led telephone counseling intervention for adults with epilepsy who self-reported sleep deprivation or alcohol use as seizure triggers. ￼ Forty intervention participants were compared with 40 controls receiving standard care, with seizure-frequency reduction and 50% responder rate assessed at 12 months. The manuscript concludes that the intervention is feasible and effective, but the current design, reporting, and statistical inconsistencies limit causal interpretation.

Overall readiness for Epilepsia

Potentially suitable after major revision

The topic is relevant to Epilepsia, but the manuscript currently overstates efficacy from a small, nonrandomized, single-center study with self-reported outcomes and incomplete reporting of allocation, follow-up, confounding, and missing data. Several statistical and reporting issues require correction before the results can be interpreted reliably.

Major concerns

* Issue: The study is described as “prospective controlled,” but the control group is defined as patients seen “prior to implementation” of STOP IT or by nonparticipating providers. This creates ambiguity about whether controls were truly contemporaneous, prospectively followed, consented in the same manner, and assessed using identical procedures.

* Why it matters: If controls were partly historical or provider-selected, the study is closer to a quasi-experimental before-after or provider-level allocation design, with substantial risk of secular, provider, and selection bias. This materially affects the validity of any efficacy inference.

* Requested revision or analysis: Clarify the exact enrollment periods for both groups, how controls were identified, whether controls were consented and followed prospectively, whether the same seizure-diary and follow-up procedures were used, and whether any provider-, calendar-time-, or clinic-level differences could have influenced outcomes. Add a participant flow diagram with screening, eligibility, enrollment, allocation, intervention completion, follow-up, and analysis denominators.

* Issue: Allocation was nonrandomized and appears dependent on program availability, provider participation, and/or patient willingness. Baseline comparability is assessed mainly with p-values, and key potential confounders are not reported.

* Why it matters: Motivation to change sleep or alcohol behavior, baseline trigger frequency, medication adherence, ASM changes during follow-up, epilepsy syndrome, seizure type, psychiatric comorbidity, sleep comorbidity, and follow-up intensity could all influence seizure outcomes independently of the intervention. The current adjusted model does not adequately address these sources of confounding.

* Requested revision or analysis: Report clinically relevant baseline characteristics beyond Table 1, including seizure types/syndromes, baseline sleep and alcohol behavior severity, ASM changes during follow-up, medication adherence if available, comorbidities, and number of clinical contacts outside the intervention. Replace or supplement baseline p-values with standardized differences. Consider propensity-score or inverse-probability approaches only if the sample and covariate data support them; otherwise, explicitly frame the study as hypothesis-generating and avoid causal language.

* Issue: The primary outcome relies on annualized seizure frequency from diaries and EHR corroboration, but the manuscript does not define diary completeness, seizure types counted, handling of missing diary intervals, or whether ascertainment intensity was equivalent between groups.

* Why it matters: The intervention group had 12 weekly contacts, which could improve diary completion, increase reporting accuracy, alter clinical management, or create attention effects not present in controls. Differential ascertainment is especially important when the outcome is self-reported seizure frequency.

* Requested revision or analysis: Specify how seizure counts were obtained for baseline and follow-up periods, whether baseline counts were retrospective or prospectively diary-based, what seizure types were included, how incomplete diaries were handled, and whether outcome assessors were blinded to group. Report diary-completion rates by group and, if possible, conduct a sensitivity analysis restricted to participants with adequate diary data.

* Issue: Missing-data handling is internally inconsistent. Methods state that patients lost to follow-up were excluded from the primary analysis and included in a last-observation-carried-forward sensitivity analysis, but Results report responder rates using denominators of 40 per group despite 3 intervention and 5 control participants being lost to 12-month follow-up. The sensitivity analysis results are not provided.

* Why it matters: The choice of denominator can materially change response rates in a small study. Last observation carried forward is also problematic for seizure frequency, particularly when baseline seizure frequency may regress toward the mean.

* Requested revision or analysis: Provide complete denominators for each outcome. State whether the primary analysis was complete-case, intention-to-treat, or modified intention-to-treat. Report the prespecified missing-data approach and all sensitivity analyses, including conservative assumptions such as counting lost participants as nonresponders for the responder endpoint.

* Issue: The reported p-value for the 50% responder comparison appears inconsistent with the stated test. Using the reported counts of 19/40 versus 8/40, a two-sided Fisher exact test gives an approximate p-value of 0.017, not 0.008; p = 0.008 is more consistent with a one-sided Fisher test.

* Why it matters: The Methods specify two-sided testing. Misreporting or mixing one-sided and two-sided tests undermines confidence in the statistical reporting.

* Requested revision or analysis: Recalculate and correct all p-values using the tests specified in Methods. If one-sided tests were used, justify them and revise the statistical analysis section accordingly; otherwise, report two-sided results consistently.

* Issue: The multivariable logistic regression model appears over-specified for the number of events. The model adjusts for intervention assignment plus age, sex, epilepsy type, number of ASMs, and baseline seizure frequency, with only 27 total responders.

* Why it matters: With approximately 27 events and at least six model terms, the adjusted odds ratio is at risk of overfitting and unstable confidence intervals. This is especially problematic in a small nonrandomized study where the adjusted model is central to the causal interpretation.

* Requested revision or analysis: Provide the full regression model specification, coefficients, standard errors, and diagnostics. Consider a more parsimonious prespecified model, penalized logistic regression, exact methods, or bootstrap confidence intervals. Present the adjusted analysis as exploratory unless the model was prespecified and adequately powered.

* Issue: No sample-size or power justification is provided.

* Why it matters: The study reports statistically significant findings, but without a priori assumptions or detectable effect-size justification, it is difficult to distinguish a planned efficacy study from an underpowered pilot with potentially unstable estimates.

* Requested revision or analysis: Add a sample-size rationale. If the study was not powered for efficacy, state this clearly and present the findings as preliminary effect estimates with confidence intervals rather than definitive evidence of effectiveness.

* Issue: The manuscript repeatedly uses efficacy language—“effective adjunct,” “can produce clinically meaningful improvements,” and “significant improvements”—despite the nonrandomized design, self-reported outcomes, and absence of an attention-matched control.

* Why it matters: The data support an association between participation in STOP IT and improved seizure outcomes, not definitive causal efficacy. The intervention effect may reflect motivation, attention, regression to the mean, concurrent treatment changes, or differential outcome ascertainment.

* Requested revision or analysis: Revise the Abstract, Key Points, Discussion, and Conclusion to use associative language. Terms such as “effective” should be replaced with “associated with” unless supported by randomized or otherwise robust causal evidence.

* Issue: The behavioral modification outcomes are insufficiently defined and incompletely reported. “Consistent adherence to target bedtime” and “meaningful reduction or cessation of alcohol consumption” are not operationalized, and corresponding control-group behavioral outcomes are not reported.

* Why it matters: The central premise is that modifying sleep and alcohol behaviors reduced seizures, but the manuscript does not demonstrate that the target behaviors changed differentially between groups or that behavior change mediated seizure reduction.

* Requested revision or analysis: Define all behavioral outcomes quantitatively, including baseline and follow-up measurement methods, denominators, and missing data. Report behavioral outcomes for both intervention and control groups. If mediation or subgroup analyses are not feasible, avoid implying that seizure reduction was mechanistically caused by improved sleep or reduced alcohol intake.

* Issue: The intervention is not described with sufficient detail for reproducibility. The manuscript does not provide the counseling script/manual, nurse training, call duration, fidelity monitoring, documentation procedures, or criteria for motivational counseling.

* Why it matters: The paper proposes a scalable clinical framework, but readers cannot reproduce or evaluate the intervention dose, content, or fidelity from the current description.

* Requested revision or analysis: Add a detailed intervention description, ideally with a supplemental protocol or checklist, including training requirements, counseling content, call structure, typical duration, fidelity assessment, and how deviations or missed calls were handled.

* Issue: Concomitant clinical management during the 12-month follow-up is not reported.

* Why it matters: ASM initiation, dose escalation, medication adherence interventions, epilepsy surgery evaluation, rescue medication changes, and additional clinic visits could explain part or all of the observed seizure reduction.

* Requested revision or analysis: Report ASM and other epilepsy-management changes during follow-up by group. If medication changes occurred, include sensitivity analyses excluding participants with major treatment changes or adjusting for these changes if the sample permits.

* Issue: Ethical and trial-reporting information contains placeholders, including study dates “XXX and XXX,” IRB Protocol “#XXX,” author names “XXX,” and ClinicalTrials.gov “NCT05XXXXXX.” The timing of trial registration relative to enrollment is not stated.

* Why it matters: This is an interventional behavioral study. Epilepsia readers and editors need complete registration, ethics, and reporting information to assess transparency, prospective design, and selective outcome reporting.

* Requested revision or analysis: Replace all placeholders with complete information. Provide the ClinicalTrials.gov identifier, registration date, enrollment start date, prespecified primary and secondary outcomes, and any deviations from the registered protocol. Include or cite a completed TREND checklist.

Minor concerns

* Issue: Figure 1 is described in the legend and manuscript text, but the submitted document does not appear to include the actual figure; only the legend is present.

* Why it matters: The primary outcome is visually summarized by this figure, and its absence limits review of distribution, outliers, and consistency with the reported Mann-Whitney U analysis.

* Requested revision: Include the actual figure with clearly labeled axes, units, sample sizes, missing-data denominators, and the statistical test used.

* Issue: The Abstract states that controls reported “identical triggers,” but the Methods and Table 1 show group-level similarity rather than individual matching.

* Why it matters: “Identical” implies a stronger design than is described.

* Requested revision: Replace “identical triggers” with more precise wording, such as “similar trigger profiles,” unless individual matching was performed and can be documented.

* Issue: Table 1 reports p-values for baseline characteristics but does not specify the tests used. Focal and generalized epilepsy are complementary categories, yet both are assigned the same p-value.

* Why it matters: Baseline testing in a small nonrandomized study can be misleading, and duplicated testing of complementary categories adds little interpretive value.

* Requested revision: Specify statistical tests in the table footnote, consider replacing p-values with standardized differences, and avoid redundant rows or clarify epilepsy categories.

* Issue: Drug-resistant epilepsy is listed in Table 1 but not defined in the Methods, despite reference 18 appearing to be included for this purpose.

* Why it matters: The interpretation of treatment resistance depends on whether a standard ILAE definition was applied and how it was adjudicated.

* Requested revision: Define drug-resistant epilepsy in Methods and state how it was determined.

* Issue: The manuscript states that sleep deprivation and alcohol use are the “two most common modifiable lifestyle-related seizure triggers,” but the Introduction should more carefully distinguish patient-reported precipitants from objectively verified triggers.

* Why it matters: Patient-reported triggers are clinically important but can be affected by recall, attribution, and reporting biases.

* Requested revision: Qualify the wording as “commonly reported” and avoid implying that all self-reported triggers are objectively established causal precipitants.

* Issue: The mechanistic statements in the Introduction are relatively specific, including GABA-mediated tonic inhibition and cortical excitability, but the manuscript does not make clear whether the cited sources directly support these mechanistic claims.

* Why it matters: Mechanistic precision is important in Epilepsia, particularly when the study itself is clinical and behavioral rather than mechanistic.

* Requested revision: Verify that references 5–7 support the specific mechanisms stated, or soften the mechanistic language.

* Issue: The Results report “median 42% reduction” and “median change: −3.5 seizures/year,” but confidence intervals or other effect-size uncertainty estimates are not provided for the primary outcome.

* Why it matters: P-values alone do not convey precision, especially in a small study with skewed seizure-frequency data.

* Requested revision: Add confidence intervals for between-group differences in percent change and absolute seizure-frequency change, using appropriate nonparametric or bootstrap methods if needed.

* Issue: The Discussion compares STOP IT with prior behavioral trials and states that it achieved “greater separation” than many prior studies, but this comparison is not sufficiently controlled for differences in design, population, comparator, outcome ascertainment, and attention control.

* Why it matters: Cross-trial comparisons can overstate novelty or relative efficacy when study designs differ substantially.

* Requested revision: Temper this comparison and contextualize STOP IT as preliminary, nonrandomized evidence rather than as superior to prior behavioral interventions.

* Issue: References 15–18 appear incompletely integrated into the manuscript text.

* Why it matters: Uncited or minimally used references suggest that the discussion and methods may not be fully aligned with the reference list.

* Requested revision: Ensure every reference is cited where relevant, particularly the updated psychobehavioral treatment review, self-management follow-up literature, and ILAE drug-resistance definition.

* Issue: The Short Summary and Key Points repeat the main efficacy claim without the design limitations.

* Why it matters: These sections may be read independently and currently present the findings more definitively than the methods support.

* Requested revision: Revise these sections to state that STOP IT was associated with improved seizure outcomes in a nonrandomized study and that randomized controlled evaluation is needed.

Review for human-generated paper supplied by Epileptologist C

**Summary**

The manuscript describes a 5-year physician-questionnaire follow-up of patients who had seizures during the “Pocket Monster” broadcast, focusing on the 25 of 91 followed patients who later had seizure recurrence. It compares recurrent cases with and without a pre-existing history of epilepsy and reports differences in age, seizure type, recurrence interval, EEG photoparoxysmal response, epilepsy syndrome classification, antiseizure medication use, and seizure status at last follow-up.

**Overall readiness for Epilepsia**

Unlikely suitable for Epilepsia in its current form. The topic is relevant to visually induced seizures and photosensitive epilepsy, but the current submission has substantial concerns about originality relative to prior reports, cohort definition, outcome classification, statistical interpretation, internal consistency of Table 1, and missing human-subjects reporting. The conclusions also exceed what can be supported by a small, selected, retrospective recurrent-case subset.

**Major concerns**

1. **Issue:** The manuscript does not clearly establish its originality relative to the already published 5-year Pocket Monster follow-up reports cited as references 7 and 8. The Introduction states that the recurrence rate was reported previously, and the current analysis appears to use the same 91-patient followed cohort and the same 25 recurrent cases.
   **Why it matters:** For Epilepsia, the incremental contribution over the prior Epilepsia follow-up publication is central to scientific value and publication ethics. Without a clear account of overlap, this could represent redundant publication or an insufficiently distinct secondary analysis.
   **Requested revision or analysis:** Explicitly state whether this is a secondary analysis of the previously published cohort, identify which data and analyses are new, describe any overlap in tables/results/text with references 7 and 8, and revise the Introduction and Discussion to articulate the novel clinical or scientific question addressed here.
2. **Issue:** Table 1 contains an apparent major internal inconsistency in the “Epilepsy syndrome (LRE/GE)” row. The table lists the Nonepilepsy group as “11:2” and the Epilepsy group as “2:10,” but the syndrome labels immediately below and the Results/Discussion indicate the opposite pattern: the Nonepilepsy group appears to include 11 generalized epilepsies and 2 localization-related epilepsies, whereas the Epilepsy group appears to include 10 localization-related epilepsies and 2 generalized epilepsies.
   **Why it matters:** This is one of the manuscript’s central findings. A reversal of the LRE/GE counts directly affects interpretability of the Results, Discussion, Abstract, and Conclusions.
   **Requested revision or analysis:** Correct the row label and/or counts, verify the p value after correction, and ensure that the Abstract, Results, Discussion, and Conclusions consistently describe the same direction of association.
3. **Issue:** The cohort definition and data flow are insufficiently described. The manuscript states that 103 patients were identified in an earlier survey, 91 had follow-up questionnaires, and 25 had recurrence, but it does not provide a flow diagram, reasons for non-follow-up, dates of ascertainment, exact geographic catchment, eligibility criteria, or how the index Pocket Monster seizure and subsequent recurrence were verified. The placeholders “XX/XXXX,” “XXXX,” and “XXX” also leave key methods unspecified.
   **Why it matters:** The analysis is restricted to recurrent cases, so selection, attrition, and ascertainment are especially important. Without clear denominators and follow-up procedures, readers cannot assess selection bias, referral bias, recall bias, or generalizability.
   **Requested revision or analysis:** Provide a complete participant flow from the original survey through the current analytic cohort; replace all date and location placeholders; define inclusion/exclusion criteria; state follow-up duration and range; explain how recurrence was confirmed; and describe the 12 patients without follow-up information, including whether they differed from those followed.
4. **Issue:** The statement that “several corrections were made on the patients’ data from the original study to the present one” is not explained.
   **Why it matters:** Corrections to previously published patient-level data can materially change recurrence classification, syndrome diagnosis, EEG findings, or group assignment. The absence of detail prevents verification and raises concern about reproducibility.
   **Requested revision or analysis:** Specify each type of correction made, how many patients were affected, which variables changed, who adjudicated the corrections, and whether any corrected values differ from the previously published reports.
5. **Issue:** The seizure and epilepsy-syndrome classifications are not sufficiently reproducible. Classification was determined by attending pediatric neurologists at multiple hospitals, but the manuscript does not report standardized diagnostic criteria, timing of syndrome assignment, whether reviewers were blinded to prior epilepsy status, whether there was central adjudication, or how ambiguous cases were handled.
   **Why it matters:** Syndrome classification is the main outcome distinguishing the two groups. Non-standardized local classification, especially in a small sample, could create differential misclassification between patients with and without prior epilepsy.
   **Requested revision or analysis:** Provide explicit diagnostic criteria for each syndrome category, state when diagnoses were assigned, describe whether classification was centralized or independently reviewed, and clarify how uncertain or insufficiently documented cases were handled.
6. **Issue:** The EEG/photosensitivity methods are not standardized. The manuscript states that intermittent photic stimulation protocols varied among hospitals and defines PPR only as generalized spike–wave induced by photic stimulation, excluding focal epileptiform responses.
   **Why it matters:** The manuscript draws conclusions about the relative role of photosensitivity, yet PPR ascertainment may not be comparable across hospitals and may under-detect focal photic responses, particularly in patients classified as having frontal or occipital lobe epilepsy.
   **Requested revision or analysis:** Describe the IPS protocols used or the range of protocol variation, provide EEG timing relative to the incident and recurrence, clarify whether focal photic responses were recorded separately, and temper conclusions about photosensitivity if standardized ascertainment cannot be demonstrated.
7. **Issue:** The statistical analysis is overextended for the sample size and study design. The analysis compares 12 versus 13 patients across many variables using multiple unadjusted tests, without confidence intervals, effect sizes, adjustment for multiple comparisons, or a prespecified primary comparison. Several comparisons also have incomplete denominators.
   **Why it matters:** With only 25 recurrent cases and many tests, nominal p values are unstable and vulnerable to both type I and type II error. The selected recurrent-case design also does not support inference about predictors of recurrence in the original 91-patient cohort.
   **Requested revision or analysis:** Reframe the analysis as primarily descriptive, identify any prespecified primary comparison if one exists, report effect sizes with exact confidence intervals where feasible, avoid overinterpreting nominal p values, and provide denominators for every variable with missing data.
8. **Issue:** Confounding is not addressed. Age, prior epilepsy activity, antiseizure medication exposure, seizure type, and epilepsy syndrome are interrelated, but the manuscript interprets group differences as though prior epilepsy history is the principal explanatory factor.
   **Why it matters:** For example, the older age and shorter recurrence interval in the Epilepsy group may reflect active pre-existing epilepsy rather than a distinct post-Pocket Monster epilepsy trajectory. Similarly, all patients in the Epilepsy group received antiseizure medications after the incident, compared with only two in the Nonepilepsy group, making clinical course comparisons difficult.
   **Requested revision or analysis:** Either perform appropriately limited adjusted or stratified analyses if feasible, or explicitly state that the sample size precludes confounder adjustment and revise causal or explanatory language accordingly.
9. **Issue:** The conclusions overstate the clinical implications. The Abstract states that the results “will be useful in order to determine the treatment” of patients with visually induced seizures, but the study does not compare treatment strategies, does not evaluate treatment response by medication or syndrome, and includes only patients with recurrence.
   **Why it matters:** The data do not support a treatment recommendation. Treatment decisions after visually induced seizures require recurrence risk estimation and treatment-outcome data, neither of which is provided by this selected recurrent-case analysis.
   **Requested revision or analysis:** Remove or substantially qualify treatment-directed claims. Limit the conclusion to descriptive differences observed among recurrent cases, and distinguish hypothesis-generating observations from clinical guidance.
10. **Issue:** Several mechanistic or temporal interpretations are speculative. The Discussion suggests that the Pocket Monster seizure in the Nonepilepsy group occurred earlier than spontaneous seizures would otherwise have appeared, and that the broadcast occurred by chance during active epilepsy in the Epilepsy group.
    **Why it matters:** These interpretations require counterfactual information or comparison groups that are not available. The study cannot determine whether the broadcast accelerated epilepsy onset, unmasked latent epilepsy, or merely coincided with an age-related epilepsy onset window.
    **Requested revision or analysis:** Recast these statements as hypotheses, not conclusions. Clearly state that the design cannot establish acceleration of epilepsy onset, causation, or the counterfactual timing of spontaneous seizures.
11. **Issue:** Human-subjects oversight is not reported. The manuscript does not state whether an ethics committee or institutional review board approved the follow-up questionnaire study, whether consent was obtained or waived, or how patient confidentiality was protected.
    **Why it matters:** The study involves human clinical data collected across hospitals. Epilepsia readers and editors need assurance that ethical oversight and consent/waiver procedures were appropriate.
    **Requested revision or analysis:** Add a dedicated ethics statement specifying approval or waiver, consent procedures, data de-identification/confidentiality protections, and the responsible institution or committee.

**Minor concerns**

1. **Issue:** The Abstract is not structured as a clear study summary. The Methods sentence reports the main recurrence result rather than describing the design, cohort source, follow-up, and analytic comparison.
   **Why it matters:** Readers cannot determine from the Abstract that this is a retrospective physician-questionnaire follow-up restricted to recurrent cases from a larger cohort.
   **Requested revision:** Revise the Abstract Methods to specify the design, source cohort, follow-up interval, analytic sample, and comparison groups; reserve recurrence rates and group differences for the Results.
2. **Issue:** The term “Nonepilepsy group” is awkward and potentially misleading, because all 13 patients in that group later appear to have been classified as having epilepsy syndromes.
   **Why it matters:** The label could be read as indicating that these patients did not have epilepsy, whereas the intended meaning is no history of epilepsy before the Pocket Monster incident.
   **Requested revision:** Use a clearer label throughout, such as “no prior epilepsy group” or “patients without pre-existing epilepsy.”
3. **Issue:** The manuscript uses older terminology, including “partial seizures,” “localization-related epilepsies,” and “antiepileptic drugs,” without explaining whether this is intentional because historical ILAE classifications were used.
   **Why it matters:** Epilepsia readers may expect current terminology, or at least a clear mapping between historical categories and current nomenclature.
   **Requested revision:** Either update terminology to current equivalents where appropriate or state explicitly that historical classifications were retained for consistency with the original data collection and prior publications.
4. **Issue:** Table 1 has incomplete or unclear denominators in several rows. For example, seizure type at recurrence totals 12 rather than 13 in the Nonepilepsy group and 11 rather than 12 in the Epilepsy group; seizure frequency within a year before last follow-up totals 12 in the Nonepilepsy group; and “Not tested” is used for some rows without explanation.
   **Why it matters:** Missing denominators affect interpretation of percentages, p values, and clinical course.
   **Requested revision:** Add denominators for every row, specify missing values, and explain why some comparisons were not tested.
5. **Issue:** The age analysis is reduced to “age <12 years” without justification or presentation of continuous age distributions.
   **Why it matters:** Age is central to the interpretation of epilepsy syndrome onset and recurrence timing, and a dichotomous threshold may obscure clinically relevant differences.
   **Requested revision:** Provide median age with range or interquartile range for each group, justify the <12-year cutoff, and consider presenting the continuous age comparison.
6. **Issue:** The medication variables are insufficiently defined. “AED after PM incident” does not distinguish newly started, continued, increased, or changed treatment; the last-follow-up medication list is descriptive but not linked to seizure outcome or indication.
   **Why it matters:** The manuscript discusses treatment relevance, but the medication data are not interpretable without knowing the timing and reason for treatment decisions.
   **Requested revision:** Define medication exposure categories and timing, and avoid implying treatment efficacy unless treatment-outcome analyses are provided.
7. **Issue:** The follow-up outcome “seizures within a year before last follow-up” is difficult to interpret because the exact last-follow-up timing and seizure-count ascertainment are not described.
   **Why it matters:** Patients may have different follow-up durations, and physician-reported seizure frequency categories may not be directly comparable across sites.
   **Requested revision:** State the timing of last follow-up for each group, define seizure-frequency categories, and clarify how seizure frequency was obtained.
8. **Issue:** The Discussion does not explicitly acknowledge the principal limitations: retrospective questionnaire design, small selected recurrent-case sample, non-standardized EEG protocols, missing data, lack of central diagnostic adjudication, absence of a non-recurrent comparison group, and limited ability to infer causality or treatment implications.
   **Why it matters:** These limitations materially affect how the findings should be interpreted.
   **Requested revision:** Add a concise limitations paragraph and align the conclusions with those limitations.
9. **Issue:** The reference list is narrow and appears to stop at early Pocket Monster reports and historical ILAE classifications.
   **Why it matters:** If submitted as a current manuscript, the work needs contextualization within subsequent literature on photosensitive epilepsy, visually induced seizures, media-related seizure provocation, and current classification terminology.
   **Requested revision:** Update the Discussion and references to include relevant later literature, or explicitly frame the manuscript as a historical secondary analysis of an older cohort.
10. **Issue:** Several sentences require tightening for precision, including “Age below 12 years and generalized seizures were more often,” “classified into idiopathic generalized epilepsies,” and “Participation of photosensitivity was relatively frequent.”
    **Why it matters:** These phrasings are imprecise and sometimes overstate statistical findings.
    **Requested revision:** Revise for grammatical accuracy and statistical precision, especially where differences are described despite non-significant comparisons.

*Reviews generated by Epileptologist C*

Review for AI-generated paper supplied by Epileptologist A

**PEER REVIEW REPORT**

*Clinician Attitudes Toward Functional Seizures and Epilepsy: A Comparative Study of Stigma*

Manuscript #57 • Journal Submission Review

**EDITORIAL RECOMMENDATION**

**MAJOR REVISIONS REQUIRED**

| **SUMMARY** | This cross-sectional vignette survey addresses a clinically meaningful and underexplored topic — stigma toward functional seizures among EMU clinicians. The study design has conceptual merit and the use of a validated instrument is a strength. However, multiple critical methodological flaws, statistical irregularities, internal inconsistencies, and over-interpreted findings require substantial revision before this paper can be considered for publication in a high-impact journal. |
| --- | --- |

**ISSUES AT A GLANCE**

| **#** | **Category** | **Issue** | **Severity** |
| --- | --- | --- | --- |
| **1** | **Statistics** | Multiple comparisons (9 domains) with no correction for Type I error inflation; all p-values reported identically as "0.001" | **Critical** |
| **2** | **Internal Inconsistency** | 100% of participants report prior education on functional seizures (Table 1), yet "Prior Education" is modelled as a significant predictor in Table 3 — a variable with zero variance cannot predict anything | **Critical** |
| **3** | **Regression Model** | Severe underpowering of multivariable model: 18 physicians, 8 APPs, 9 EEG techs used with multiple predictors; high risk of overfitting and collinearity | **Critical** |
| **4** | **Vignette Design** | Vignettes differ on multiple clinical dimensions beyond the diagnostic label, confounding attribution of stigma differences to the diagnosis alone | **Major** |
| **5** | **Qualitative Methods** | Thematic analysis lacks rigour: no coder details, no inter-rater reliability, percentages reported of commenters not total sample | **Major** |
| **6** | **Citation Mismatch** | Multiple references are incorrectly assigned or used for claims they do not support (e.g., Refs 9, 13, 17) | **Major** |
| **7** | **Non-response Bias** | Bias assessment is mentioned but results are never reported | **Major** |
| **8** | **Over-interpretation** | Causal language and prescriptive recommendations exceed what a single-site cross-sectional study can support | **Major** |
| **9** | **AQ Instrument** | Sub-scale internal consistency as low as 0.52 undermines validity; use of composite score across heterogeneous domains is questionable | **Minor** |
| **10** | **Manuscript Errors** | Stray quotation mark in Methods; Table 3 labels "5 years" without “<” operator; missing blinded institution/date placeholders | **Minor** |

| **1** | **CORE RESEARCH QUESTION AND SIGNIFICANCE** |
| --- | --- |

**1.1 Conceptual Significance**

The research question is clinically important and timely. Stigma in functional neurological disorder is increasingly recognized as a barrier to equitable care, and the comparative framing — functional seizures versus epilepsy within the same clinical unit — is methodologically elegant in concept. The study's focus on EMU clinicians, who are the first-line professionals at the critical diagnostic juncture, is particularly well-motivated.

**1.2 Gaps and Framing**

However, the authors claim this is "the first systematic comparison" of clinician stigma between functional seizures and epilepsy. This claim is not adequately supported by the literature review. The authors cite MacDuffie et al. (2020) only once in passing, yet that paper explicitly maps the research agenda for stigma in FND within the clinical encounter — a far more thorough engagement is warranted. The authors should also engage more substantively with Barnett et al. (2022) and with the broader healthcare provider stigma literature across psychiatric and functional conditions (e.g., stigma toward medically unexplained symptoms) to contextualise what are and are not novel contributions.

Additionally, the claim of "first systematic comparison" is undermined by the absence of a formal scoping review or documented search strategy. Without one, the assertion cannot stand and should be softened (e.g., "among the first") pending a more thorough literature search.

| **2** | **STRENGTH AND VALIDITY OF THE METHODOLOGY** |
| --- | --- |

**2.1 Study Design**

The within-subject, vignette-based cross-sectional design is appropriate for the research question and permits controlled comparison. Randomisation of vignette order to minimise order effects is commendable. The 75% response rate is strong for a survey study and reduces non-response bias risk, though the bias assessment is mentioned but never reported (see Section 2.8 below).

**2.2 Attribution Questionnaire (AQ) — Instrument Validity Concerns**

The use of the Attribution Questionnaire is a reasonable choice given its previous use in healthcare provider stigma research. However, several concerns warrant discussion:

- The AQ was originally developed to measure general public attitudes toward mental illness, not clinician attitudes toward neurological conditions. Its construct validity in this specific context requires explicit justification or reference to validation studies in comparable healthcare provider populations.
- The Cronbach's alpha range of 0.52–0.91 across subscales is notable. A value of 0.52 falls well below the conventional 0.70 threshold for acceptable internal consistency. The authors acknowledge this briefly but do not address which specific subscales demonstrated poor reliability, nor whether findings in those domains should be interpreted cautiously or excluded.
- The use of a composite total AQ score (summing all 9 domains into a single number) aggregates conceptually orthogonal constructs — responsibility, pity, coercion, dangerousness — into a single metric. This risks obscuring the clinically meaningful heterogeneity that is actually the paper's most interesting finding. The composite score analysis should be clearly designated as secondary.

**2.3 Vignette Design — A Fundamental Confound**

This is one of the most significant methodological concerns in the paper. The two vignettes differ not only in diagnostic label, but on multiple clinical features simultaneously:

| **Feature** | **Functional Seizure Vignette** | **Epilepsy Vignette** |
| --- | --- | --- |
| **Ictal duration** | 5–7 minutes | ~90 seconds |
| **Motor semiology** | Asynchronous, fluctuating | Rhythmic, generalised |
| **Eye position** | Closed | Open |
| **Ictal awareness** | Partial recall | Complete amnesia |
| **Prodrome character** | Anxiety/dyspnoea | Epigastric aura |
| **Postictal features** | Tearful | Myalgia, confusion |
| **EEG correlation** | No epileptiform activity | Generalised discharges |

These differences mean respondents may have formed stigma-relevant impressions based on the clinical features before reading the diagnostic label. For example, the functional seizure vignette's prolonged events with closed eyes and partial awareness more readily maps onto lay concepts of "psychological" or "controllable" illness. It is impossible to determine, in the current design, whether the large responsibility attribution effect (d = 1.12) reflects the diagnostic label, the clinical presentation, or their interaction. The authors must acknowledge this as a fundamental limitation — not merely list it as one of several limitations — and discuss how future studies might use label-only or label-swap designs to disentangle these effects.

**2.4 Critical Statistical Concerns**

***2.4a Multiple Comparisons Without Correction***

The authors report nine separate domain-level comparisons, all at p < 0.001, with no correction for multiple comparisons. This inflation of Type I error rate is a critical flaw. A Bonferroni-corrected threshold of p < 0.0056, or a Benjamini-Hochberg false discovery rate procedure, must be applied. While the large effect sizes suggest many findings would survive correction, the authors cannot claim statistical significance at conventional thresholds without addressing this.

***2.4b Suspicious p-value Reporting***

Every domain comparison in Table 2 is reported as p = 0.001, including null findings for help-seeking (reported p = 0.34) and segregation (p = 0.42). For significant comparisons, the probability that all nine tests yield exactly the same p-value of 0.001 is vanishingly small unless the authors have rounded or truncated values. Exact p-values (e.g., p < 0.001 where appropriate, or the precise computed value) must be reported in accordance with standard scientific reporting norms.

***2.4c Regression Model Overfitting***

The multivariable model in Table 3 includes at least seven predictors applied to a sample of 75 participants, with the smallest subgroups numbering 8 (APPs) and 9 (EEG technologists). The conventional rule of thumb of 10–15 observations per predictor is substantially violated. Variance inflation factors and collinearity diagnostics should be reported. Furthermore, epilepsy fellowship training is modelled as a predictor nested within physician role — a structural collinearity that must be addressed, either through interaction terms or stratified models.

**2.5 Critical Internal Inconsistency: The "Prior Education" Predictor**

Table 1 reports that 100% of respondents (75/75) had prior education on functional seizures. Yet in Table 3, "Prior Education on Functional Seizures" is included as an independent variable with a statistically significant coefficient (β = −3.8, 95% CI: −6.5 to −1.1, p = 0.006).

A variable with zero variance — where all participants share the same value — cannot be modelled as a predictor in any regression framework. Its inclusion as a significant predictor is mathematically impossible and represents either a data coding error, an error in Table 1, or a fundamental analytical mistake. This inconsistency must be resolved before the paper can be considered further.

**2.6 Qualitative Methodology**

The qualitative component is described as "thematic analysis" but lacks the methodological transparency expected by high-impact journals:

- The number of coders and their professional backgrounds are not reported.
- No inter-rater reliability statistic (e.g., Cohen's kappa, percentage agreement) or reflexivity statement is provided.
- Percentages (47%, 38%, 32%, 21%) appear to be calculated from the 34 who provided comments, not from the full sample of 75. If so, this must be stated explicitly; as written, readers will assume n = 75 as the denominator.
- The qualitative findings section draws strong interpretive conclusions from single illustrative quotations. For a study claiming qualitative components, a richer evidential base is required, or the qualitative section should be explicitly framed as exploratory and hypothesis-generating only.

**2.7 Missing Variables and Unmeasured Confounders**

The study does not measure or adjust for:

- Frequency of prior clinical encounters with functional seizure patients (beyond years of experience), which may mediate familiarity effects.
- Burnout or emotional exhaustion scores, which may independently predict stigmatising attitudes and are plausibly elevated in nursing staff.
- Whether respondents had previously received communication training in delivering the functional seizure diagnosis.
- Racial/ethnic background, which has been shown to influence stigma perceptions in FND (Von Lersner et al., 2019, cited in the paper but underutilised).

**2.8 Non-response Bias Assessment Not Reported**

The Methods section states that non-response bias was assessed by comparing characteristics between responders and non-responders. The Results section does not present these findings. Given the 75% response rate, the 25% non-response could represent a systematic pattern (e.g., clinicians with strong or unusual attitudes selectively declining). The non-response analysis must be reported.

| **3** | **KEY FINDINGS AND EVIDENCE** |
| --- | --- |

**3.1 Strengths of the Findings**

Subject to the methodological caveats above, the core finding — that EMU clinicians exhibit greater stigma toward functional seizures than epilepsy, particularly on the responsibility, pity, anger, and coercion domains — is internally consistent with attribution theory and aligns with qualitative literature. The finding that the dangerousness domain runs in the opposite direction (higher for epilepsy) is clinically plausible and represents a nuanced contribution that is appropriately interpreted.

The identification of fellowship training as a protective factor (d = −6.4) is potentially important and warrants the emphasis given to it, provided the collinearity and sample size concerns are addressed.

**3.2 Over-Interpretation Concerns**

- The paper describes its qualitative finding that clinicians feel there is "nothing more to offer" after diagnosis as a "critical misconception." While there is face validity to this characterisation, using a single free-text quotation to anchor a broad interpretive claim — and then prescribing educational remediation — goes beyond what the data support. This should be reframed as a hypothesis.
- Section 4.4 makes specific recommendations (e.g., CBT integration, multidisciplinary EMU teams) that are not supported by the data in this paper. These extend well beyond the scope of the findings and belong in a brief mention of future directions, not as primary recommendations.
- Causal language is used throughout the Discussion (e.g., fellowship training "substantially reduces stigmatising attitudes"; less experienced clinicians show stigma "suggesting that clinical experience… may naturally attenuate stigmatising attitudes") despite the explicitly cross-sectional design. Such language must be replaced with appropriately conditional phrasing.

| **4** | **MAJOR LIMITATIONS AND GAPS IN THE STUDY** |
| --- | --- |

**4.1 Limitations Acknowledged in the Paper**

The authors appropriately acknowledge single-site design, cross-sectional methodology, social desirability bias, vignette limitations, and small sample size. These are genuine and significant. However, their placement in a brief "limitations" section at the end of the Discussion, without adequate integration into the framing of results and conclusions, dilutes their impact. The vignette confound in particular (see Section 2.3 above) is listed as a minor limitation when it is in fact the most fundamental threat to the paper's central interpretive claim.

**4.2 Gaps Not Acknowledged**

- No generalisability discussion for non-academic, community, or international settings, which may have markedly different patient populations, staffing ratios, and training exposure.
- The paper does not address whether stigma scores are stable over time within clinicians, which has direct implications for the educational interventions proposed.
- There is no discussion of whether the AQ's stigma domains map directly onto observable clinical behaviours (consultation length, referral patterns, medication prescribing), which is the ultimate clinically meaningful outcome.
- The study was conducted in a single country (implied but never stated). Cross-cultural variation in stigma toward FND is well-documented and relevant to the generalisability claims.

**4.3 Literature Gaps**

The literature review does not adequately engage with:

- The broader healthcare provider stigma literature for medically unexplained symptoms (MUS) and somatoform disorders, within which functional seizures sit, and which has a large body of evidence that directly contextualises these findings.
- Reuber and colleagues' substantial body of work specifically on provider-patient relationships in PNES/functional seizures.
- Studies examining whether stigma reduction interventions in related conditions (e.g., mental illness, chronic pain) translate to improved patient outcomes — relevant to the recommendations made.
- The Cullinane et al. (2025) scoping review on FND in healthcare education is cited but deserves more substantive engagement given the paper's educational recommendations.

| **5** | **METHODOLOGICAL AND PRACTICAL CONTRIBUTIONS** |
| --- | --- |

**5.1 Methodological Contributions**

- First use of the AQ in a within-subject vignette paradigm comparing functional seizures to epilepsy in a specialised inpatient neurology workforce.
- Vignette randomisation is an important design feature that partially addresses order effects.
- Mixed-methods approach (quantitative + qualitative free-text) provides contextual depth.
- Adherence to STROBE reporting standards is appropriate and commendable.

**5.2 Practical Contributions (If Validated)**

- Identification of nurses and early-career clinicians as higher-risk groups for stigmatising attitudes provides actionable targets for educational initiatives.
- The finding that epilepsy fellowship training is protective could inform fellowship curriculum development and accreditation requirements.
- The qualitative themes around diagnostic uncertainty and therapeutic helplessness complement and explain the quantitative data, pointing toward specific content for educational interventions.

| **6** | **CITATION AND REFERENCE CONCERNS** |
| --- | --- |

**6.1 Specific Misattributions**

The following reference assignments require correction:

- Reference 9 (Hallett et al., Lancet Neurology 2022) is a landmark mechanistic review of FND subtypes and shared mechanisms. It is cited multiple times for clinical claims about stigma reduction, diagnostic disclosure effectiveness, and provider frustration — content that does not appear in this review. These citations should be replaced with primary evidence sources.
- Reference 13 (Weiner et al., 1988 — foundational attribution theory paper) is cited in Section 4.1 for the statement that "coercive treatment approaches… could undermine the therapeutic alliance." The therapeutic alliance claim is not attribution theory; the correct citation should be a psychotherapy or clinical care reference.
- Reference 17 in the reference list is Fusunyan et al. (2024) on a web-based educational module. In Section 4.4, reference 17 is cited for "interdisciplinary collaboration between neurology, psychiatry, and psychology." These are different claims and the citation appears transposed.
- Reference 8 (McLoughlin et al., 2026) is dated to the future in the reference list — this should be confirmed as an accepted/in-press publication with the correct in-press citation format.

| **7** | **MINOR EDITORIAL COMMENTS** |
| --- | --- |

**7.1 Manuscript Errors**

- Methods section (Recruitment): Stray closing quotation mark at the end of the paragraph on monetary compensation: '…for their time spent in survey completion."' The opening quote mark is absent.
- Table 3 (Years of Experience row): The label reads "5 years" where context requires "<5 years." Clarify the operator.
- The abstract states participants responded to "matched clinical vignettes" but the vignettes differ on multiple clinical dimensions beyond the diagnosis — this should be revised to "parallel" or "corresponding" vignettes.
- Institution names and IRB study numbers are appropriately blinded as XXX, but the recruitment date range should be preserved as [Month Year]–[Month Year] to allow readers to contextualise temporal context.

**FINAL EDITORIAL RECOMMENDATION**

**EDITORIAL RECOMMENDATION**

**MAJOR REVISIONS REQUIRED**

This manuscript addresses a clinically important and under-studied question with a reasonable study design. The research question is well-motivated, the use of a validated instrument is appropriate, and the mixed-methods approach adds contextual value. However, the paper cannot be accepted in its current form due to critical methodological and statistical concerns that materially threaten the validity of the reported findings.

**Required Actions Before Resubmission**

- **Resolve the internal inconsistency between 100% prior education rate (Table 1) and the inclusion of prior education as a predictor (Table 3). Either reanalyse or correct the data, and provide a transparent explanation.**
- **Apply appropriate multiple comparisons correction across the nine AQ domain tests and report exact p-values.**
- **Report and interpret the non-response bias analysis as stated in the Methods.**
- **Provide collinearity diagnostics for the multivariable regression model and address the fellowship training / physician role collinearity.**
- **Acknowledge the vignette confound as a primary limitation and discuss its implications for interpreting the central responsibility attribution finding.**
- **Strengthen the qualitative methodology section with coder details, inter-rater reliability, and correct denominators.**
- **Correct all identified citation misattributions.**
- **Replace causal language throughout the Discussion with appropriately conditional phrasing.**
- Address internal consistency values below 0.70 by identifying affected subscales and qualifying their interpretation.
- Correct manuscript typographic errors (stray quotation mark, Table 3 label).

*Reviewer note: I encourage the authors to pursue revision, as the topic is important and the underlying study design has merit. Addressing these concerns rigorously would produce a substantially stronger and publishable paper.*

Reviewer expertise: Neurology • Epilepsy • Functional Neurological Disorder • Clinical Trials Methodology

Review for human-generated paper supplied by Epileptologist A

**PEER REVIEW REPORT**

*Manuscript: Re-PROGRAM: The evaluation of a brief intervention program for patients with functional seizures in an outpatient hospital setting*

Journal: High-Impact Neurology / Epilepsy Journal | Review Type: Blinded Expert Peer Review

Reviewer expertise: Neurology, Epilepsy, Functional Neurological Disorders, Clinical Trial Methodology

**EDITORIAL RECOMMENDATION: MAJOR REVISIONS REQUIRED**

### **SUMMARY ASSESSMENT**

This manuscript presents a retrospective evaluation of Re-PROGRAM, a novel five-session, telehealth-delivered, skills-based intervention for functional seizures (FS). The topic is clinically important and timely: FS represent a significant proportion of epilepsy clinic referrals, carry substantial disability burden, and currently lack a robust portfolio of brief, scalable, evidence-based interventions. The authors are commended for addressing a genuine gap in the field and for the program's theoretical grounding in neurobiological and psychophysiological models of FS. The 100% session completion rate across all 29 enrolled patients is noteworthy and suggests strong acceptability.

However, the manuscript in its current form has significant methodological deficiencies that prevent its findings from being interpreted with confidence. These include the absence of prospective baseline seizure data, a custom unvalidated outcome instrument, a 55% survey return rate with a median completion delay of 229 days, no control group, and insufficient statistical analysis. Several claims in the Discussion and Abstract are not adequately supported by the data and require tempering. Major revisions are required before this work can be considered for publication.

### **1. CORE RESEARCH QUESTION AND SIGNIFICANCE**

The core research question — whether Re-PROGRAM is feasible and acceptable as a brief outpatient intervention for functional seizures — is clearly stated and clinically meaningful. Functional seizures remain among the most challenging presentations in neurology and psychiatry, and the persistent gap between diagnosis and effective, accessible treatment is well-documented. The program's emphasis on neurobiological psychoeducation and practical seizure self-management strategies, rather than exclusive reliance on trauma-focused or psychodynamic frameworks, is a rational and potentially important innovation, particularly for patients without a trauma history or significant psychiatric comorbidity.

The brief, telehealth-delivered format addresses a recognized barrier to care in this population and has clear real-world scalability implications. The significance of the research question is therefore high. However, the study is framed beyond what its design can support. A feasibility and acceptability evaluation of a pilot cohort of 29 is appropriate, but the manuscript at times draws on language and conclusions more befitting a controlled efficacy trial.

### **2. METHODOLOGY: STRENGTHS AND CRITICAL CONCERNS**

#### **2.1 Diagnostic Validity**

The authors are commended for establishing FS diagnosis through video-EEG in 28 of 29 patients, which represents the gold standard. The single exception (clinical epileptologist review of out-of-hospital video) is acknowledged and constitutes an acceptable deviation given clinical practicalities. The inclusion of three patients with comorbid epilepsy is appropriately disclosed, and the authors note these individuals could distinguish their two seizure types. However, no further details are provided on how FS episodes were tracked separately from epileptic events in this subgroup, which is a potential confound that should be addressed explicitly.

#### **2.2 Outcome Measurement — The FSPICS Instrument**

The Functional Seizures Pre-post Intervention Comparison Survey (FSPICS) is the primary quantitative outcome measure, yet it was purpose-designed for this study. This raises several significant concerns:

- No psychometric validation data are reported. Internal consistency (Cronbach's alpha), test-retest reliability, construct validity, and convergent validity against established instruments (e.g., PHQ-9, GAD-7, PSQI, SF-36, or Patient Global Impression of Change) are entirely absent.
- The 5-point retrospective comparison scale is subject to well-documented recall bias, particularly given that the median interval between program completion and survey return was 229 days (IQR: 103-268 days) — nearly eight months. Retrospective self-assessment across this timeframe is highly unreliable and may fundamentally undermine the quantitative findings.
- The inclusion of five reverse-scored items to reduce response bias is acknowledged but insufficient in the absence of formal validation. The paper does not report whether these items functioned as intended or whether the scale demonstrates adequate discriminant validity.
- Pre-intervention seizure frequency data are based on retrospective recall, not prospective seizure diary. This is a critical methodological flaw: without a prospective baseline, it is impossible to determine whether observed reductions represent genuine treatment effects, regression to the mean, natural disease fluctuation, or recall bias.

CRITICAL CONCERN: The FSPICS requires formal psychometric validation before it can serve as the primary quantitative outcome in any publication. At minimum, authors must report Cronbach's alpha, inter-item correlations, and convergent validity with at least one established measure. The absence of prospective baseline seizure diary data renders the quantitative efficacy findings uninterpretable as evidence of treatment effect.

#### **2.3 Survey Response Rate and Selection Bias**

Only 16 of 29 enrolled patients (55.2%) returned the post-intervention questionnaire, and of those, only 12 were complete. The authors attempt to justify this rate by citing a reference on email survey methodology (Schaefer & Dillman, 1998), which is inappropriate: this reference concerns commercial survey design, not clinical outcome measurement, and achieving a 55-60% response rate is far below acceptable standards for primary outcome collection in clinical research. The non-responders are a critically important group. While the authors compare basic demographic and clinical characteristics between responders and non-responders (Table S3), they do not report whether seizure burden differed between groups before the intervention, nor do they test whether non-response is independently associated with a lack of improvement. The risk of survivorship bias and social desirability bias is high. Four patients were additionally lost to clinical follow-up entirely, further limiting the ability to assess outcomes for the full enrolled cohort.

#### **2.4 Study Design — Absence of a Control Group**

The lack of a randomized control group or even an active comparator means that observed improvements in seizure frequency cannot be attributed to Re-PROGRAM specifically. Several alternative explanations are not addressed:

- Natural disease course and spontaneous remission: FS can remit spontaneously, particularly in the early post-diagnosis period, and this cohort had a median time from onset to diagnosis of approximately one year or less in 48% of cases.
- Regression to the mean: Patients are typically referred and recruited at times of high seizure burden, and subsequent improvement may reflect natural fluctuation rather than treatment response.
- Hawthorne effect and therapeutic attention: Any structured clinical engagement, regardless of specific content, may improve outcomes through increased attention, rapport, and sense of support.
- Concurrent interventions: Psychopharmacological treatment was permitted to continue during the study period; changes in medications are not tracked or controlled for.

The CODES trial (Goldstein et al., 2020), cited by the authors, is the methodological benchmark for this field, and the present study falls substantially short of its design standards. While this is acknowledged and a forthcoming RCT is referenced, the discussion should more carefully circumscribe interpretations to feasibility and acceptability rather than treatment efficacy.

#### **2.5 Statistical Analysis**

The statistical methods section is inadequate. A single Fisher's exact test is reported — examining the association between pre-/postintervention change in seizure occurrence and time from onset to diagnosis (p = .853). This is the only inferential statistic in the paper, and it addresses a secondary research question. No formal statistical testing is applied to the primary outcomes of seizure frequency reduction or FSPICS scores. While a study of this size (N=16 survey completers) limits inferential power, at minimum the authors should:

- Report descriptive statistics (mean, SD, or median and IQR) for all 24 FSPICS items, both overall and by response category.
- Apply a non-parametric within-subjects comparison (e.g., Wilcoxon signed-rank test) to change scores where applicable.
- Report effect sizes with 95% confidence intervals to permit readers to assess clinical meaningfulness.
- Acknowledge the multiple comparisons problem given the breadth of outcomes described.

#### **2.6 Therapist Training and Fidelity**

Re-PROGRAM was administered by three provisional psychologists described as neuropsychology interns with 'little to no experience in providing therapy of any specific type.' While this is positioned as a strength for scalability, it also raises questions about treatment fidelity. No formal fidelity monitoring is reported. Were sessions recorded and rated for adherence to the manual? Were there inter-therapist differences in outcomes? This is particularly important when the same cohort serves as evidence for the program's effectiveness.

#### **2.7 Eligibility Criteria and Sample Characterization**

The exclusion criteria are described in vague terms: patients were required to be 'cognitively and psychiatrically stable and free of acute risk issues.' No standardized instruments are cited for assessing these criteria (e.g., cognitive screening tools, risk assessment scales). The baseline psychiatric comorbidity profile of the sample is not described in Table 1 beyond comorbid epilepsy — rates of depression, anxiety disorders, PTSD, or personality disorders are not reported, yet these are directly relevant to treatment response and generalizability. The concurrent psychopharmacological treatment permitted as standard care is not characterized.

### **3. FINDINGS AND EVIDENCE BASE**

The key finding — that all 29 enrolled patients completed all scheduled sessions — is genuinely notable and the most robust finding in the paper. Session completion rates in FS trials are notoriously low, and this result, if reproducible, speaks strongly to the program's tolerability and acceptability.

Seizure reduction outcomes, however, must be interpreted with considerable caution:

- The claim that '15 of 16 survey respondents reported a reduction in seizure frequency' is presented prominently, but the true denominator for the intent-to-treat population is 29. The effective reduction from the most favorable perspective is 15/29 (51.7%), not 93.75%. The paper should consistently use the enrolled sample as the denominator.
- Among the non-survey group, outcome data are available for only 9 of 13 patients (from medical records), with 4 lost to follow-up. Of these 9, 8 reported seizure reduction. However, clinical follow-up documentation is an imprecise and potentially biased substitute for prospective, standardized outcome measurement.
- The report does not quantify the magnitude of seizure reduction — a 5% decrease and a 90% decrease are both 'reductions.' Without frequency counts or seizure diary data, the clinical significance of reported improvements cannot be assessed.
- No validated measure of seizure severity or functional impairment (e.g., Quality of Life in Epilepsy [QOLIE], Liverpool Seizure Severity Scale, or Functional Neurological Symptom Scale) was employed.

The qualitative findings are the most compelling element of this paper. The four themes — functional status, agency, interpersonal development, and interpersonal communication — are coherent and consistent with the program's stated therapeutic aims. The patient quotations are illuminating and add genuine clinical texture. However, the qualitative methodology requires more rigorous reporting (see Section 4.2 below).

### **4. MAJOR LIMITATIONS AND GAPS**

#### **4.1 Absence from the Literature Review**

- The ReACT (Retraining and Control Therapy) program developed by Fobian and colleagues (cited in the reference list) has notable conceptual similarities to Re-PROGRAM and has been evaluated in both pediatric and adult populations. A direct and detailed comparison is largely absent from the discussion.
- The paper does not engage with the growing literature on predictors of treatment response in FS (e.g., seizure frequency at baseline, psychiatric comorbidity load, duration of illness prior to diagnosis), which is directly relevant to identifying who is most likely to benefit from Re-PROGRAM.
- The influence of early intervention on prognosis is mentioned briefly (Reference 48, Arain et al., 2007) but this literature is richer and more nuanced than a single citation suggests. The CODES trial sub-analyses on moderators of treatment response are not discussed.
- The paper makes reference to neuroinflammation as a potential FS mechanism (Reference 22, Sharma & Szaflarski, 2021) but does not note that this is a hypothesis paper without direct empirical support in the present context.
- The evolving diagnostic terminology in the field (functional seizures, dissociative seizures, PNES) is not discussed. While the authors use 'functional seizures' consistently — the preferred contemporary term — they use the outdated term 'pseudoseizure' in the diagnostic reference (Reference 34, Dworetzky, 2014). This should be updated.

#### **4.2 Qualitative Methodology Reporting**

The qualitative analysis is described as 'thematic analysis using NVivo,' but the methodology is inadequately described. A credible thematic analysis requires:

- A named and cited framework (e.g., Braun & Clarke's reflexive thematic analysis).
- Description of the coding process — who conducted initial coding, and how many independent coders were involved.
- Reporting of inter-rater reliability or a rationale for single-coder analysis.
- A reflexivity statement addressing the positionality of the researchers conducting the analysis, particularly given that one or more analysts may have been involved in delivering the intervention.

#### **4.3 Sample Generalizability**

The sample is predominantly female (75.9%), relatively young (mean age 28.38 years), and single (72.4%). While this broadly reflects the demographics of FS populations, the sample is drawn from a single outpatient centre, limiting geographic and cultural generalizability. The intervention is delivered in English, restricting its current applicability to English-speaking patients. The paper should acknowledge these constraints explicitly.

#### **4.4 Redacted Information**

The study site and enrollment dates are redacted for blinding purposes (denoted as 'XXX'). While this is understandable for peer review, the manuscript must ensure this information is restored in any published version. The absence of enrollment dates also prevents readers from contextualizing the study in relation to the COVID-19 pandemic, which had substantial implications for telehealth delivery and patient mental health during the likely study period.

### **5. OVER-INTERPRETATION OF FINDINGS**

Several statements in the manuscript overstate the evidence and should be revised:

| **As Written** | **Concern** |
| --- | --- |
| *"All patients who enrolled in Re-PROGRAM completed all scheduled sessions" (Abstract & Results)* | Accurate as stated but the framing implies comprehensive follow-up data, which the 55% survey return rate contradicts. The abstract should distinguish between session completion and outcome data availability. |
| *"100% adherence rate, demonstrating acceptability and feasibility" (Key Points)* | Session completion does not equal acceptability in the clinical research sense. Acceptability requires broader measurement including patient burden, satisfaction, and qualitative feedback — which is provided, but the 100% figure is misleading when 45% of outcome data are missing. |
| *"Almost all patients who completed the postintervention survey reported reduced functional seizure frequency" (Discussion)* | This statistic references only 16 patients. The denominator must be clearly stated throughout the paper. The phrase 'almost all' obscures that one patient did not report improvement. |
| *"This retrospective evaluation demonstrates the feasibility and acceptability... and warrants further investigation" (Abstract Significance)* | This is an appropriate conclusion. The wording should be maintained, but equivalent qualifying language should be applied consistently throughout the Discussion, where efficacy language currently dominates. |
| *Neurobiological rationale in Table 2 (e.g., nutrition, exercise, sleep targets specific brain networks)* | The claimed mechanistic targets are highly speculative. The evidence base for most of these is derived from non-FS populations, and the paper acknowledges there is 'no definitive mechanistic evidence for FS.' The confident causal language in Table 2 misrepresents the evidence base. |

### **6. METHODOLOGICAL AND PRACTICAL CONTRIBUTIONS**

Despite the limitations outlined above, this work makes several genuine contributions:

- It introduces a theoretically grounded, brief intervention that extends beyond trauma-focused paradigms and may benefit a broader range of FS patients.
- The telehealth delivery model and administration by provisionally trained psychologists demonstrates scalability that is directly relevant to resource-constrained healthcare systems.
- The 100% session completion rate, if replicated in a controlled trial, would be a meaningful finding in a condition associated with high dropout from longer-term therapies.
- The FSPICS, though currently unvalidated, represents an attempt to develop a FS-specific outcome measure. Formal validation would benefit the field.
- The qualitative data, though limited in sample size, provide valuable patient-reported insights into mechanisms of change that can inform future trial design and theoretical development.
- The paper situates Re-PROGRAM clearly within the treatment gap literature and provides a transparent rationale for the program's design, which aids reproducibility.

### **7. SPECIFIC COMMENTS TO AUTHORS**

#### **Major Comments (Required Before Re-evaluation)**

1. Prospective seizure baseline data: Future reports of this intervention should incorporate a minimum 4-week prospective seizure diary as baseline measurement. For the current manuscript, the authors must explicitly acknowledge that all pre-intervention seizure frequency data are retrospective self-report subject to recall bias, and reframe all efficacy claims accordingly.
2. FSPICS validation: The authors must either (a) provide psychometric properties of the FSPICS (internal consistency, test-retest reliability, convergent validity) or (b) acknowledge in the limitations that the scale is unvalidated and reframe conclusions as preliminary only. Concurrent administration of at least one validated functional outcomes measure (e.g., QOLIE-31, PHQ-9, GAD-7, Work and Social Adjustment Scale) is strongly recommended for the ongoing RCT.
3. Intent-to-treat reporting: All outcome statistics must be reported using N=29 as the primary denominator. A clear CONSORT-style participant flow summary should be provided, even in the absence of randomization, to transparently account for survey non-responders, medical-record-only outcomes, and lost-to-follow-up cases.
4. Survey delay: The median 229-day gap between program completion and survey return is a critical finding that must be foregrounded in the Results and Limitations sections. The authors must discuss how retrospective recall across this interval affects reliability and should consider whether any prospective data from earlier time points (e.g., first clinical follow-up) could be used as a sensitivity analysis.
5. Natural history and spontaneous remission: The Discussion must address the possibility that observed seizure reductions reflect natural disease course, regression to the mean, or Hawthorne effects. Reference to base rates of spontaneous remission in newly diagnosed FS cohorts is required.
6. Qualitative methodology: The thematic analysis section must be substantially expanded to include the analytical framework used, number and independence of coders, inter-rater reliability or reflexive rationale for single-coder design, and a researcher positionality statement — particularly regarding any overlap between treatment delivery and analysis.
7. Comorbid epilepsy subgroup: The three patients with comorbid epilepsy require more detailed reporting. How were FS episodes distinguished from epileptic seizures in outcome tracking? Were these patients analyzed separately in any sensitivity analysis?

#### **Minor Comments (Recommended Revisions)**

- The term 'pseudoseizure' in Reference 34 (Dworetzky, 2014) is outdated and should be replaced with a more current diagnostic reference (e.g., LaFrance et al., 2013 ILAE consensus or Perez et al.).
- Baseline psychiatric comorbidities (depression, anxiety, PTSD, personality disorder) and concurrent psychotropic medication use should be reported in Table 1.
- The sample's ethnic and cultural diversity should be described where available.
- The Discussion's engagement with ReACT (Fobian et al.) should be expanded to a detailed comparison of theoretical frameworks, session structure, outcome measures, and results.
- Figure 2 would benefit from accompanying numerical data (exact counts per response category) to allow independent interpretation.
- The rationale for telehealth delivery should include a brief acknowledgment of the potential limitations (e.g., digital access inequity, reduced therapeutic rapport for some patients).
- The study site and enrollment dates should be restored in the final manuscript and, where relevant, the COVID-19 telehealth context should be addressed.
- Concurrent psychopharmacological changes during the study period should be described or, at minimum, acknowledged as a potential confound.

### **8. EDITORIAL RECOMMENDATION**

**RECOMMENDATION: MAJOR REVISIONS**

This paper addresses a clinically important and underserved area with a theoretically coherent and practically designed intervention. The session completion data are promising, and the qualitative findings are valuable. However, the quantitative outcome data are substantially undermined by the retrospective design, unvalidated primary outcome measure, delayed and incomplete survey return, and absence of prospective baseline seizure measurement. The Discussion overreaches the study's capacity to support efficacy conclusions.

The manuscript has genuine potential as a feasibility and acceptability report — and that is precisely how it should be reframed throughout. If the authors address the major methodological concerns identified above, temper the language of the Discussion to reflect the study's observational and retrospective character, and provide more rigorous qualitative methodology reporting, this paper could make a meaningful contribution to the FS treatment literature and serve as a valuable precursor to the authors' ongoing RCT.

The authors are encouraged to resubmit a revised manuscript with a detailed point-by-point response to each concern raised in this review.

*End of Review — Prepared by Expert Peer Reviewer in Neurology, Epilepsy, and Clinical Trial Methodology*

Review for AI-generated paper supplied by Epileptologist B

**PEER REVIEW REPORT**

*Confidential — For Editorial Use Only*

| **Manuscript** | *Randomized Controlled Pilot Trial of Daily Chicken Soup in Drug-Resistant Focal Epilepsy (Brief Communication)* |
| --- | --- |
| **Study Type** | Single-center, parallel-group, open-label randomized controlled pilot trial |
| **Sample Size** | N = 30 randomized (15 per arm); 21 evaluable at primary analysis |
| **Reviewer Expertise** | Neurology / Epilepsy / Clinical Trial Methodology |

**EDITORIAL RECOMMENDATION: MAJOR REVISIONS**

*The manuscript is transparently written and reports a null result honestly, both of which are commendable. However, severe differential attrition (47% vs. 13%), the absence of a baseline characteristics table, critical ambiguity in how the outcome direction is described, a missing reference, an inadequately justified biological mechanism, and the lack of supplementary analyses (per-protocol, sensitivity) require substantial revision before the paper can be considered for publication.*

### **SECTION 1: OVERVIEW AND SIGNIFICANCE OF THE RESEARCH QUESTION**

The manuscript presents a parallel-group randomized controlled pilot trial evaluating whether daily chicken soup supplementation reduces seizure frequency in adults with drug-resistant focal epilepsy (DRE). The clinical problem addressed — seizure burden in the approximately one-third of people with epilepsy who remain uncontrolled on antiseizure medications — is of genuine and substantial public health importance. The motivation to explore low-cost, accessible adjunctive interventions is intellectually sound given that many DRE patients are not candidates for surgery or neuromodulation.

However, the biological rationale for the specific intervention under study is weak. The proposed mechanistic pathway — (1) chicken soup has anti-inflammatory properties in vitro; (2) neuroinflammation contributes to epileptogenesis; (3) therefore, chicken soup may reduce seizures — involves two inferential leaps without supporting in vivo, animal-model, or pharmacokinetic data. The sole in vitro citation (Rennard et al., 2000) demonstrated inhibition of neutrophil chemotaxis in a laboratory setting. There is no evidence that bioavailable anti-inflammatory compounds reach the central nervous system at concentrations relevant to seizure threshold modulation. This gap substantially weakens the scientific premise of the study and must be addressed more explicitly in the Introduction.

The labeling of this as a pilot trial is appropriate given its scale. However, the stated rationale for conducting a pilot — and what specific parameters this pilot was intended to estimate for powering a future definitive trial (e.g., expected effect size, standard deviation, attrition rate, feasibility) — is not articulated. A pilot trial without explicit feasibility objectives lacks a clear purpose beyond hypothesis generation, which may limit its contribution to the literature.

### **SECTION 2: METHODOLOGY — STRENGTHS AND CRITICAL CONCERNS**

#### **2.1 Strengths**

- Computer-generated, block-randomized allocation stratified by baseline seizure frequency is appropriate and reduces confounding by seizure severity.
- Prospective, standardized seizure diaries during both baseline and treatment periods are correctly used.
- A 2-month baseline period provides a reasonable estimate of pretreatment seizure rate, though 3 months would have been more robust given the known variability of seizure counts.
- Pre-specification of the primary outcome metric (percentage change in monthly seizure rate) and analysis approach (Mann-Whitney U, LOCF for withdrawals) is stated and appropriate in principle.
- The choice of Mann-Whitney U for non-normally distributed seizure-count data is statistically defensible.
- Ethical conduct statements, conflict of interest disclosures, and funding transparency are adequately reported.

#### **2.2 Critical Methodological Concerns**

**Major Concern #1 — Severe and Differential Attrition**

The most serious methodological problem in this trial is the markedly asymmetric dropout rate: 46.7% (7/15) in the chicken soup arm versus 13.3% (2/15) in the control arm. This is not merely a quantitative concern; the reasons for withdrawal are entirely absent from the manuscript. In dietary intervention trials, dropout commonly occurs because: (a) the intervention is burdensome, unpalatable, or introduces adverse effects; or (b) participants perceive no benefit and disengage. In this trial, nearly half the chicken soup participants withdrew, which may itself constitute indirect evidence that the intervention was either poorly tolerated or ineffective from participants' subjective perspectives.

The application of Last Observation Carried Forward (LOCF) under these conditions is critically problematic. LOCF assumes that withdrawn participants' last available seizure rates represent their counterfactual outcome at trial completion. This assumption is implausible when dropout is informative — i.e., when withdrawal is related to treatment outcome. Reference 13 (Mallinckrodt et al., 2001), which the authors themselves cite, explicitly advocates for mixed-effects models over LOCF precisely because LOCF fails under informative dropout. The authors must address this contradiction.

At minimum, a per-protocol analysis (restricted to completers) and a sensitivity analysis using multiple imputation under a range of assumptions about the missing data mechanism must be provided as supplementary material. The manuscript should also include a CONSORT-style participant flow diagram and a table listing reasons for withdrawal per arm.

**Major Concern #2 — Absence of a Baseline Characteristics Table**

The manuscript contains no table presenting participant demographics, epilepsy characteristics, antiseizure medication burden, or baseline seizure rates by group. Without this, it is impossible to assess whether randomization achieved balance, whether the two groups are clinically comparable, or whether any observed between-group difference in outcomes could be attributed to a confounding baseline variable. This is a fundamental reporting requirement for any randomized trial. A Table 2 reporting baseline characteristics (age, sex, epilepsy duration, seizure type(s), number of prior ASMs, current ASM regimen, baseline monthly seizure rate) must be added.

**Major Concern #3 — Paradoxical and Insufficiently Explained Control Group Outcome**

The median percentage change in the control arm is reported as −39.47% — indicating that participants who continued their usual diet experienced a median seizure frequency increase of approximately 39% during the treatment period. This finding is highly counterintuitive and unexplained. In placebo-controlled epilepsy trials, control arms typically show modest improvements (5–20% reduction) due to regression to the mean, placebo response, and the Hawthorne effect. A near-40% worsening in the control group requires explicit explanation and is a potential red flag for informative attrition bias: participants in the control arm who were experiencing worsening seizures may have remained in the trial precisely because they were seeking help, whereas those doing well may have been less motivated to remain, inflating the apparent worsening.

Furthermore, the abstract states that 'the between-group distribution of seizure change favored chicken soup numerically.' This framing may mislead readers. The numerical apparent 'advantage' for chicken soup derives entirely from an unexplained deterioration in the control group, not from a meaningful improvement in the treatment group (chicken soup median: +7.69%). This distinction must be made explicit and prominently in both the abstract and discussion.

**Major Concern #4 — Intervention Lacks Standardization**

'Chicken soup once daily' is not a defined, reproducible intervention. The manuscript does not specify: (a) whether the soup was home-prepared, commercially produced, or standardized by the research team; (b) its composition (broth concentration, fat content, vegetable content, volume per serving); (c) its sodium content, which may be pharmacologically relevant in patients already on multiple medications; (d) any dietary or nutritional guidance provided to participants; or (e) how adherence was assessed or monitored. Without a standardized and measurable intervention, the trial cannot be replicated, and the intervention variable is confounded by enormous heterogeneity. A detailed description of the intervention following TIDieR (Template for Intervention Description and Replication) guidelines must be provided.

**Major Concern #5 — Undefined Pilot Study Objectives**

Pilot trials exist to estimate parameters for a subsequent definitive trial — primarily feasibility (recruitment rate, retention, intervention adherence) and preliminary effect size with confidence intervals for sample size calculation. The manuscript does not state what feasibility endpoints were prespecified, does not report recruitment rates, does not provide 95% confidence intervals around the primary effect estimate, and does not use the results to generate a sample size calculation for a future trial. If the study is to be presented as a pilot, it must deliver pilot-relevant outputs.

**Moderate Concern #6 — Open-Label Design and Reporting Bias**

The absence of blinding and placebo control in a seizure-frequency trial introduces multiple sources of bias: differential diary completion vigilance, expectation effects, social desirability bias in reporting, and differential healthcare-seeking behavior between arms. While the authors acknowledge this limitation, they do not discuss the known magnitude of placebo response in epilepsy clinical trials (typically 10–20% responder rates in control arms of double-blind trials), which further contextualizes the unexpected −39.47% in the control group. This context should be added to the discussion.

**Moderate Concern #7 — Participant Age Range Includes Minors**

The eligibility criterion specifies 'aged 16 years or older,' yet the methods section does not address the specific regulatory or ethical considerations applicable to the 16–17-year age bracket. In most jurisdictions, participants in this age range require parental or guardian co-consent. Whether any such participants were enrolled, and how consent was handled, should be clarified.

**Minor Concern #8 — Missing Reference #12**

The reference list skips from reference 11 to reference 13; reference 12 is absent entirely. This creates a gap in the citation record and suggests a citation was deleted during revision without updating numbering. All in-text citations must be cross-checked and renumbered accordingly.

**Minor Concern #9 — Baseline Period Duration**

A 2-month baseline period, while commonly used, may underestimate the true baseline seizure rate for patients with highly variable seizure counts. A 3-month baseline, consistent with the screening period already completed, would have provided a more stable estimate and is used in higher-quality DRE trials. The authors should acknowledge this as a limitation and discuss its potential impact on effect estimates.

### **SECTION 3: KEY FINDINGS AND EVIDENCE SUPPORTING THEM**

The primary analysis (Mann-Whitney U = 60.0, p = 0.1389) and secondary analysis (50% responder rate: 15.4% vs. 14.3%, Fisher exact p = 1.0) consistently and unambiguously indicate no statistically significant benefit of chicken soup over control. The authors interpret this correctly as a null result, and the abstract and discussion language is appropriately conservative.

However, two presentation issues must be corrected:

- The abstract language 'the between-group distribution of seizure change favored chicken soup numerically' is potentially misleading (see Major Concern #3 above). It should be rewritten to clarify that this numerical difference is driven by unexpected control-arm deterioration, not by clinically meaningful improvement in the treatment arm.
- No confidence intervals are reported for any effect estimates. For a pilot trial where imprecision is expected and must be quantified, 95% CIs are essential. The Mann-Whitney U statistic should be supplemented with a Hodges-Lehmann estimator and 95% CI for the median difference. Responder rates should include exact 95% CIs (Clopper-Pearson).

The 50% responder rate of ~15% in both arms is consistent with published placebo response rates in double-blind epilepsy trials, which further argues against any specific treatment effect of chicken soup.

### **SECTION 4: GAPS IN LITERATURE REVIEW AND OVER-INTERPRETATION RISKS**

#### **4.1 Literature Gaps**

The Introduction omits several bodies of literature that are directly relevant and would contextualize the study more rigorously:

- Placebo response rates in epilepsy clinical trials: The literature on placebo and nocebo effects in DRE trials is well-characterized (e.g., Rheims et al., 2008, Lancet Neurology) and should be cited. The observed ~15% responder rate in both arms aligns with this literature and is informative.
- Adult dietary interventions beyond the ketogenic diet: The modified Atkins diet, low glycemic index treatment (LGIT), and medium chain triglyceride diet have RCT or prospective data in adults with DRE and deserve mention to situate this work within the broader dietary intervention literature.
- Neuroinflammation as a seizure biomarker: The Introduction cites neuroinflammation as the mechanistic bridge, but does not cite any evidence that dietary anti-inflammatory interventions (with established anti-inflammatory profiles, unlike chicken soup) influence neuroinflammatory biomarkers or seizure outcomes in humans. This gap makes the mechanistic chain purely speculative.
- Regression to the mean in epilepsy trials: Patients are typically enrolled during periods of elevated seizure frequency ('enrichment bias'), and spontaneous improvement unrelated to treatment is expected. The Introduction should acknowledge this phenomenon as a key reason placebo-controlled design matters.
- TIDieR reporting guidelines for dietary interventions: The absence of any reference to intervention reporting standards (Hoffmann et al., 2014, BMJ) is a notable omission for a dietary trial.

#### **4.2 Risk of Over-Interpretation**

The manuscript is generally well-guarded against over-interpretation in its current form. The authors explicitly label findings as 'hypothesis-generating only' and state that data 'do not establish efficacy.' These formulations are appropriate and should be preserved in any revision. The one area of residual over-interpretation risk is the use of phrases like 'favored chicken soup numerically' in the abstract without adequate qualification — this should be revised as noted above.

The Discussion appropriately identifies that future trials would need 'a more rigorous design, including clear allocation concealment, a matched control condition, prespecified adherence monitoring, and an analysis strategy less sensitive to informative dropout.' These are all valid and important conditions. The authors might additionally state whether, in their view, the results from this trial — given the severe attrition — are sufficient to justify the cost and participant burden of a larger, properly designed trial.

### **SECTION 5: METHODOLOGICAL AND PRACTICAL CONTRIBUTIONS**

Despite its significant limitations, the manuscript makes the following modest contributions:

- It provides a rare example of an honestly reported null result in a dietary intervention trial for epilepsy. Publication bias against null results is well-documented in the epilepsy literature, and transparent reporting of negative or inconclusive data is of genuine value.
- It documents the feasibility challenge of a dietary intervention trial in DRE, specifically the high attrition rate in the intervention arm (~47%), which is itself informative for future trial designers and argues strongly against pursuing a larger trial with the same design.
- The use of a prospective 3-month screening period to confirm eligibility before a 2-month baseline, while resource-intensive, is methodologically sound and reduces the misclassification of transiently high-seizure-frequency patients as DRE.

The practical contribution to clinical practice is minimal and correctly characterized as such by the authors: chicken soup cannot be recommended as an adjunctive treatment for DRE on the basis of these data.

### **SECTION 6: STRUCTURED COMMENTS TO THE AUTHORS**

The following numbered comments are provided for systematic response. Each comment is categorized as Essential (must be addressed before the paper can be reconsidered) or Recommended (important for quality but may be addressed with explanation if infeasible).

| **C1** | **Essential — Attrition Bias —** Provide a CONSORT participant flow diagram showing screening, eligibility, randomization, dropout (with reasons per arm), and analysis populations. Add a supplementary per-protocol analysis restricted to completers, and a sensitivity analysis using multiple imputation under missing-at-random and missing-not-at-random assumptions. Discuss explicitly why LOCF is used despite its known limitations under informative dropout, given that the cited Mallinckrodt (2001) reference itself recommends mixed-effects models. |
| --- | --- |

| **C2** | **Essential — Baseline Characteristics Table —** Add a table reporting demographic and clinical characteristics by randomized arm (age, sex, epilepsy duration, focal seizure subtype, etiology if known, number of prior and current ASMs, baseline monthly seizure rate, any relevant comorbidities). Without this, the adequacy of randomization and the comparability of groups cannot be assessed. |
| --- | --- |

| **C3** | **Essential — Paradoxical Control Arm Deterioration —** The median −39.47% change in the control arm (indicating seizure worsening) is unexpected and requires detailed explanation. Investigate and report whether this result is attributable to: (a) informative retention bias (those doing worse remaining in the control arm); (b) chance in a very small sample; (c) unusually low baseline seizure rates in control completers inflating the negative percentage. Include individual-level data (median plot or forest-style individual response plot) in supplementary material to allow readers to assess this. |
| --- | --- |

| **C4** | **Essential — Rewrite Abstract Framing —** The statement 'the between-group distribution of seizure change favored chicken soup numerically' misrepresents the result pattern and should be rewritten to accurately state that the apparent numerical difference is driven by unexplained worsening in the control arm, not by clinically meaningful improvement in the treatment arm (median change in chicken soup arm: +7.69%). |
| --- | --- |

| **C5** | **Essential — Intervention Standardization —** The intervention must be defined in reproducible detail following TIDieR guidelines: specify soup type (commercial or standardized recipe), composition, volume, sodium content, preparation method, who administered or supplied it, and how adherence was monitored (e.g., diary, container return, or other method). |
| --- | --- |

| **C6** | **Essential — Pilot Objectives and Confidence Intervals —** State explicitly what feasibility parameters this pilot was designed to estimate. Provide 95% confidence intervals for all effect estimates (Hodges-Lehmann estimator for the median difference; Clopper-Pearson CIs for responder rates). If feasible, provide a sample size calculation for a hypothetical future definitive trial based on the observed effect estimate and variance, with appropriate caveats given the attrition. |
| --- | --- |

| **C7** | **Essential — Missing Reference 12 —** Reference 12 is absent from the reference list. Verify all in-text citations, identify the missing reference, and update the list accordingly with correct sequential numbering throughout the manuscript. |
| --- | --- |

| **C8** | **Recommended — Biological Rationale —** Substantially expand the mechanistic rationale or moderate its ambition. The current argument (in vitro neutrophil chemotaxis inhibition → neuroinflammation reduction → seizure reduction) requires CNS penetration of bioactive compounds from dietary chicken soup at therapeutically relevant concentrations. Either cite evidence that this occurs or clearly acknowledge that the mechanistic link is highly speculative and untested, and frame the study as exploratory accordingly. |
| --- | --- |

| **C9** | **Recommended — Placebo Response Contextualization —** Add a sentence in the Discussion citing published placebo and control arm responder rates from double-blind epilepsy trials. The observed ~15% responder rates in both arms of this trial are consistent with that literature, which further contextualizes the null result. |
| --- | --- |

| **C10** | **Recommended — Minor Age Eligibility Clarification —** Clarify the consent procedures for participants aged 16–17 years, including whether parental or guardian co-consent was obtained per local regulation and whether any such participants were enrolled. |
| --- | --- |

| **C11** | **Recommended — Adverse Event Reporting —** Report any adverse events, tolerability issues, or dietary concerns (e.g., sodium intake, allergic reactions, gastrointestinal effects) in the intervention arm. The high dropout rate in the chicken soup arm raises the possibility of tolerability contributing to withdrawal, which has direct implications for feasibility of a future trial. |
| --- | --- |

| **C12** | **Recommended — Broader Dietary Literature —** Briefly cite the modified Atkins diet and LGIT evidence in adults with DRE in the Introduction to situate the study within the full dietary intervention landscape, not only the ketogenic diet. |
| --- | --- |

### **SUMMARY SCORECARD**

| **Domain** | **Assessment** | **Required Action** |
| --- | --- | --- |
| Clinical importance of topic | High | None |
| Biological rationale | Weak — speculative CNS link | Strengthen or moderate (C8) |
| Randomization quality | Adequate | Clarify allocation concealment |
| Attrition management | Critical flaw — 47% vs 13% | Sensitivity analyses required (C1) |
| Intervention standardization | Absent | TIDieR description required (C5) |
| Outcome reporting | Partially misleading | Rewrite abstract framing (C4) |
| Statistical methods | Appropriate but incomplete | Add CIs; sensitivity analysis (C6) |
| Baseline comparability | Unreported | Add Table 2 (C2) |
| Literature review | Incomplete | Expand dietary & placebo lit (C12, C9) |
| Conclusion appropriateness | Appropriately conservative | Preserve; minor clarifications |
| Reference integrity | Reference 12 missing | Correct and renumber (C7) |

*Final Recommendation: MAJOR REVISIONS. The authors are encouraged to respond point-by-point to each numbered comment. Comments C1–C7 are essential and must be addressed in full for the manuscript to be reconsidered. If the differential attrition bias cannot be adequately addressed through supplementary analyses and the control-arm deterioration cannot be explained, the editorial team should consider whether the study provides sufficient inferential value for publication.*

Review for human-generated paper supplied by Epileptologist B

**PEER REVIEW REPORT**

*Confidential — For Editorial Use Only*

| **Manuscript Title** | Early seizure freedom with [18F]fluorodopa PET response after isocitrate dehydrogenase inhibition with vorasidenib: First case report |
| --- | --- |
| **Article Type** | Brief Communication / Case Report |
| **Reviewer Expertise** | Neurology, Epilepsy, Neuro-Oncology, Clinical Trials Methodology |
| **Review Date** | May 8, 2026 |

**OVERALL SUMMARY**

This case report describes a 52-year-old male with grade 2 IDH1 R132H-mutant, 1p/19q-codeleted oligodendroglioma who achieved seizure freedom within one month of initiating vorasidenib, accompanied by a metabolic partial response on [18F]fluorodopa (F-DOPA) PET at 3 and 6 months despite stable disease on MRI. The manuscript addresses two clinically relevant and largely unanswered questions in the management of IDH-mutant low-grade gliomas: (1) whether IDH inhibition can improve seizure control, and (2) whether amino acid PET can detect early treatment response ahead of conventional MRI.

The topic is timely, given vorasidenib's recent FDA approval and the conspicuous exclusion of patients with uncontrolled seizures from the pivotal INDIGO trial. The authors also draw attention to a potentially important confound — the known capacity of F-DOPA PET to capture ictal/peri-ictal metabolic activity — yet do not fully resolve the interpretive tension this creates. Several methodological and interpretive concerns must be addressed before the manuscript is suitable for publication.

**1. CORE RESEARCH QUESTION AND SIGNIFICANCE**

**1.1 Strengths**

- The central dual question — vorasidenib and seizure control, plus F-DOPA PET as an early response biomarker — is well-motivated by a genuine gap in the literature arising from the INDIGO trial's exclusion of patients with uncontrolled seizures.
- The mechanistic rationale linking IDH-mutant D-2-hydroxyglutarate (2-HG) production to epileptogenesis (glutamate mimicry, mTOR hyperactivation) is plausible and appropriately cited.
- The case is embedded in an ongoing prospective institutional cohort, lending greater context and implying that systematic data collection is underway.
- The first-report claim of vorasidenib + seizure freedom + F-DOPA PET response appears defensible given the current literature and is genuinely novel.

**1.2 Concerns**

- **CRITICAL:** Scope inflation:
- The abstract states this report 'highlights the potential of vorasidenib for seizure management and the value of F-DOPA PET for early treatment assessment.' This language implies a level of evidence that a single uncontrolled case report cannot provide. The authors should constrain language throughout to hypothesis-generating and exploratory conclusions.
- **MINOR:** Mechanistic underdevelopment:
- The mechanistic discussion reduces epileptogenesis in IDH-mutant gliomas to 2-HG mimicking glutamate. However, 2-HG's effects on GABAergic interneurons, neuronal hyperexcitability via mTOR pathway activation (Mortazavi et al., already cited), and epigenetic dysregulation affecting ion channel expression are underexplored. A more nuanced mechanistic framing would strengthen the scientific contribution.

**2. STRENGTH AND VALIDITY OF METHODOLOGY**

**2.1 Seizure Assessment**

| **MAJOR** | Absence of objective electrophysiological validation of seizure freedom is the most significant methodological gap in this manuscript. |
| --- | --- |

- Seizure frequency and freedom rely exclusively on a patient-completed diary reviewed at monthly visits. While patient diaries are standard in clinical practice, they are subject to recall bias, under-reporting (particularly of brief auras or nocturnal events), and cannot exclude subclinical electrographic seizures.
- No ambulatory EEG, video-EEG monitoring, or neurophysiological endpoint was employed at any timepoint. This is particularly relevant because the patient's seizures had prominent subjective components (epigastric aura, paresthesias) that are difficult to distinguish from non-epileptic events without EEG correlation.
- The paper defines seizure freedom as 'complete absence of seizures or auras according to ILAE outcome classification,' which is appropriate. However, the follow-up duration of 8 months is modest; the Engel/ILAE sustained seizure freedom benchmark is commonly applied at 12 months. The temporal claim of 'early' freedom is reasonable but the durability remains unestablished.
- Critically, spontaneous fluctuation in seizure frequency is common in grade 2 gliomas, particularly in the peri-operative period and during tumor stabilization. Without a formal run-in period or pre-treatment seizure frequency baseline over several months, the attribution of seizure freedom specifically to vorasidenib cannot be made with confidence. This must be explicitly and prominently acknowledged as a limitation.

**2.2 F-DOPA PET Methodology**

| **MAJOR** | The manuscript reports F-DOPA PET as evidence of tumor metabolic response, but does not adequately address the critical confound that F-DOPA PET uptake may partly reflect seizure-related metabolic activity rather than tumor biology alone. |
| --- | --- |

- The authors themselves cite the Morana et al. (2018) case report demonstrating increased F-DOPA uptake in epileptogenic tissue and peritumoral parenchyma during status epilepticus. Given that the present patient was seizing approximately twice weekly prior to vorasidenib and became seizure-free on therapy, a substantial proportion of the dramatic MTV reduction (16.3 cm³ → 8.0 cm³ → 4.0 cm³) may reflect the cessation of peri-ictal amino acid hypermetabolism rather than — or in addition to — tumor metabolic response. This is a fundamental interpretive challenge that is only briefly mentioned and must be addressed prominently.
- The dissociation between SUVmax (unchanged at 3 months: 3.2 → 3.2; declining only at 6 months: 2.5) and MTV (reduced by ~51% at 3 months) is striking and unexplained. If vorasidenib were reducing tumor metabolic activity uniformly, one would expect parallel reductions in both metrics. The selective MTV reduction without corresponding SUVmax change could indicate peripheral shrinkage of a seizure-active metabolic rim rather than central tumor cell activity reduction. This interpretation should be discussed.
- **MAJOR:** Essential PET acquisition details are absent:
- Scanner model, acquisition protocol, and reconstruction algorithm are not reported.
- Time from radiotracer injection to scan initiation is not stated.
- Patient fasting status prior to each scan is not reported — this is standard for amino acid PET to minimize background variation.
- No information is provided on whether the patient was in a seizure-free period (e.g., >24 hours without a seizure event) at the time of baseline PET. If a seizure occurred in close temporal proximity to the baseline scan, the baseline MTV may be systematically inflated, amplifying the apparent treatment response.
- Inter-observer variability and intra-observer reproducibility for MTV segmentation using the 1.6x background threshold are not reported. MTV is highly sensitive to threshold choice and scanner-specific calibration. Without reproducibility data or sensitivity analysis, the quantitative precision of the reported MTV values cannot be assessed.
- While PET RANO 1.0 criteria are appropriately cited for response classification, the authors should clarify whether the imaging protocol meets all PET RANO 1.0 technical requirements (standardized acquisition conditions, background reference region selection, etc.).

**2.3 MRI Volumetry**

- The 3D FLAIR volumetric measurements (32.2 → 31.5 → 28.1 cm³) are reported as 'nonsignificant.' In a single patient, formal statistical significance testing is not applicable. The authors likely mean that the change does not meet RANO criteria for partial response (≥50% volume reduction). This should be stated explicitly.
- The approximately 12.7% volume reduction from baseline to 6 months is not trivial. While below the RANO partial response threshold, this magnitude of change warrants comment in the context of the concurrent F-DOPA PET response.
- No information on volumetric measurement uncertainty or inter-rater reproducibility for MRI segmentation is provided.

**2.4 Blood 2-HG Monitoring**

- The authors state that serial blood 2-HG monitoring is 'ongoing at the authors' institution' but report no data. Given the central mechanistic argument linking 2-HG to both seizure generation and tumor metabolism, the absence of any pharmacodynamic biomarker data is a notable gap. Even preliminary or descriptive 2-HG values would substantially strengthen the report's mechanistic contribution.

**3. KEY FINDINGS AND EVIDENCE QUALITY**

**3.1 Seizure Freedom**

The observation of seizure freedom from the first month of vorasidenib, without any change in ASM, is the most clinically compelling finding. The mechanism is plausible and the timing is consistent with the pharmacodynamics of IDH inhibition. However, the evidence quality is limited by: (a) reliance on subjective patient diary, (b) absence of EEG confirmation, (c) inability to exclude spontaneous fluctuation or natural disease trajectory, and (d) short follow-up relative to established seizure freedom benchmarks. The finding should be presented as a preliminary signal warranting prospective study, not as evidence of efficacy.

**3.2 F-DOPA PET Response**

The reported F-DOPA PET partial response (50% MTV reduction at 3 months, >75% at 6 months) is striking but must be interpreted with caution given: (a) the unresolved seizure-activity confound described above, (b) the absence of standardized acquisition details, and (c) the divergence between MTV and SUVmax trajectories. The finding does advance the hypothesis that F-DOPA PET can provide earlier evidence of biological change than MRI volumetry in vorasidenib-treated patients, consistent with the FET PET data from Galldiks et al. (already cited). The extension of this observation from FET to F-DOPA PET is a meaningful incremental contribution.

**3.3 MRI Stability**

The MRI-stable but PET-responsive pattern is a potentially important observation supporting the use of functional imaging for early treatment assessment. However, the 6-month follow-up is insufficient to determine whether this represents a genuinely dissociated response or a temporal lag in volumetric change. Longer follow-up is needed before firm conclusions can be drawn about the relative predictive value of PET vs. MRI in this therapeutic context.

**4. LITERATURE REVIEW AND CONCEPTUAL GAPS**

**4.1 Existing Gaps in the Literature Review**

- The manuscript does not reference registry-based or observational data on seizure outcomes in vorasidenib-treated patients outside the INDIGO trial (if any exist at institutional or expanded-access program level). Readers should be informed about the totality of available evidence.
- The review of seizure outcomes with prior glioma-directed therapies (radiation, PCV, temozolomide) correctly cites Koekkoek et al. and Bruno et al., but does not address whether IDH inhibitors in non-glioma contexts (e.g., enasidenib/ivosidenib in AML) have produced any neurological or seizure-related observations that could inform mechanistic understanding.
- The claim that '2-HG is exported into the microenvironment, and the excitatory effects on neurons are rapid and do not seem to depend on a direct action on ionotropic glutamate receptors' is presented without adequate primary citation or mechanistic detail. The distinction between ionotropic and metabotropic glutamate receptor involvement, and the role of NMDA receptor modulation, should be clarified or cited more precisely.
- The potential interaction between levetiracetam and vorasidenib via hepatic enzyme induction/inhibition pathways is not addressed. Levetiracetam is not a major CYP substrate, but the absence of comment on pharmacokinetic interactions is an oversight in a case report focused on a pharmacological intervention.
- The broader landscape of amino acid PET tracers in glioma response assessment (MET, FET, FACBC) and their comparison to F-DOPA in this indication is not contextualised. Given that the Galldiks et al. (2024) paper used FET rather than F-DOPA, the specific advantages and limitations of F-DOPA in this context should be discussed.

**5. MAJOR LIMITATIONS AND INTERPRETIVE CONCERNS**

**5.1 Attribution and Causality**

| **CRITICAL** | The manuscript insufficiently addresses alternative explanations for both the seizure freedom and the PET response. A case report cannot establish causality, but a rigorous report must systematically evaluate and transparently acknowledge confounds. |
| --- | --- |

- For seizure freedom: natural history fluctuation, placebo effect, regression to the mean after a high-frequency period, and pharmacokinetic interactions with levetiracetam are not discussed as alternative explanations.
- For PET response: the seizure-activity confound (discussed above) is the most significant, but disease natural history and intra-scanner variability should also be addressed.

**5.2 Follow-up Duration**

- Eight months of seizure-free follow-up is insufficient to draw conclusions about sustained seizure control. Grade 2 gliomas are slow-growing tumors with prolonged clinical courses; minimum 12- to 24-month follow-up is conventionally reported in the seizure outcomes literature for this population.

**5.3 Generalizability**

- The patient's tumor characteristics are favorable: grade 2, 1p/19q-codeleted oligodendroglioma (the most treatment-responsive IDH-mutant glioma subtype), low Ki-67, intact CDKN2A/B, no prior radiotherapy or chemotherapy. These features limit generalizability to astrocytomas (IDH-mutant, non-codeleted), higher-grade tumors, or post-adjuvant-therapy patients.

**5.4 Single-ASM Monotherapy Baseline**

- The patient elected not to add a second ASM, which, while simplifying attribution analysis, also represents a deviation from standard clinical escalation practice. The decision-making process and the clinical reasoning for not escalating ASM prior to initiating vorasidenib should be stated explicitly.

**5.5 Redaction of Dates**

- The redaction of all specific dates (denoted 'XX XXXX' throughout) makes it impossible to reconstruct the clinical timeline, including the interval between symptom onset and surgery, between surgery and vorasidenib initiation, and between vorasidenib initiation and each imaging timepoint. While patient anonymisation is necessary, the authors should report time intervals in months (e.g., 'vorasidenib was initiated 3 months after surgery') rather than masking the timeline entirely. This is a minimum reporting standard for a case report.

**6. METHODOLOGICAL AND PRACTICAL CONTRIBUTIONS**

- This case report makes three contributions of varying strength:
- First report of seizure freedom under vorasidenib documented with standard ILAE outcome criteria. This is clinically valuable as a hypothesis-generating signal that should inform the design of formal seizure endpoints in future trials.
- First application of F-DOPA PET (as opposed to FET PET) for monitoring vorasidenib response, extending the pharmacological imaging literature for this new therapy.
- Raises the hypothesis that F-DOPA PET uptake in epileptogenic gliomas may have a seizure-activity component, with potential implications for PET-based response interpretation in this population.
- The manuscript falls short of demonstrating a methodologically rigorous case report standard in its current form, particularly regarding PET acquisition documentation, seizure outcome objectification, and confound analysis.

**7. SPECIFIC COMMENTS TO THE AUTHORS**

**7.1 Abstract**

- Line: 'F-DOPA PET revealed a significant decrease in tracer uptake starting from the third month' — SUVmax was unchanged at 3 months (3.2 → 3.2); only MTV decreased. The word 'significant' cannot be applied to a single patient without statistical framing. Revise to accurately distinguish between the metrics and their trajectories.
- The abstract should explicitly note the seizure-activity confound as a limitation of the PET interpretation.

**7.2 Introduction**

- The FDA approval year appears as 'XXXX' — this should be populated (2024) in the final version.
- 'D-2-hydroxyglutarate...plays a key role in epileptogenesis by mimicking the excitatory neurotransmitter glutamate' — this is a simplification. Please revise to acknowledge additional mechanisms (mTOR, epigenetic dysregulation of ion channels) with appropriate citations.

**7.3 Materials and Methods**

- Add PET acquisition parameters: scanner model, injected activity, scan duration, time post-injection, reconstruction method, and attenuation correction approach.
- State whether baseline PET was acquired during a seizure-free interval. If not, this must be noted as a potential source of inflated baseline MTV.
- Report inter-observer variability for MTV segmentation or confirm single-rater measurement with stated limitations.
- The ILAE reference (Beniczky et al., 2025) postdates the clinical events described and may not correspond to the classification used at the time of care. Clarify which version of the ILAE classification was prospectively applied.

**7.4 Results**

- Specify time intervals in months rather than redacted date placeholders for all clinical events.
- Report the precise timing of baseline PET relative to surgery and vorasidenib initiation.
- The 28.1 cm³ FLAIR volume at 6 months represents a ~12.7% reduction from baseline (32.2 cm³). This should be reported as a percentage change alongside the absolute values.
- Describe what the patient was doing in terms of seizure activity in the days prior to each PET scan. Was the patient seizure-free before each scan? This is critical for interpreting baseline vs. on-treatment PET.

**7.5 Discussion**

- Prominently and explicitly discuss the seizure-activity confound in F-DOPA PET interpretation. The current treatment (two sentences at the end of one paragraph) is insufficient given its centrality to the study's main claims.
- Add a paragraph on alternative explanations for seizure freedom (natural history, placebo, regression to the mean) and explicitly state why these are considered less likely (or acknowledge that they cannot be excluded).
- Discuss why F-DOPA PET was selected over FET PET in the context of the existing Galldiks et al. FET data. What is the added value or specific rationale for F-DOPA in this population?
- Comment on the potential pharmacokinetic interaction between levetiracetam and vorasidenib, even if only to state that no clinically significant interaction is expected.
- The section on blood 2-HG monitoring being 'ongoing' should either be removed from this case report (since no data are presented) or reframed to explain why it was not yet available for this index case.
- Future direction sentence: 'Future trials...should investigate the efficacy of IDH inhibitors on pharmacoresistant seizures' — consider specifying that this would require randomized or at minimum controlled designs given the confounds illustrated by this very case.

**8. EDITORIAL RECOMMENDATION**

**EDITORIAL RECOMMENDATION**

**MAJOR REVISIONS**

The manuscript addresses a clinically important and timely question and makes a genuine first-report contribution. However, it requires substantial revision before it can be considered for publication. The priority issues are:

- Seizure-activity confound in F-DOPA PET interpretation must be addressed prominently and systematically — this is the most critical scientific concern.
- PET acquisition methodology must be fully reported in accordance with PET RANO 1.0 and standard nuclear medicine reporting requirements.
- The clinical timeline must be reported in months rather than redacted date placeholders.
- Claims of 'significance' and causal language must be revised to reflect the hypothesis-generating nature of a single case report.
- Alternative explanations for seizure freedom must be explicitly discussed.
- The mechanistic narrative on 2-HG and epileptogenesis should be expanded and more precisely cited.

*If these concerns are adequately addressed, the revised manuscript may represent a worthwhile contribution to the emerging literature on vorasidenib's non-oncological effects and the utility of amino acid PET for early treatment monitoring in IDH-mutant gliomas.*

2.3 **Editorial Decisions**

The full text of each the editorial decision for each epileptologists’ papers are below.

*Editorial Decisions A*

Epileptologist A’s AI-Generated Paper Decision

**Editorial Decision: Major Revisions**

**Both documents converge on the same conclusion, which I agree with: this manuscript requires Major Revisions before it can be considered for publication in *Epilepsia*.**

**Rationale for Decision**

**The topic is important and appropriate for *Epilepsia*—clinician stigma toward functional seizures (FS) versus epilepsy is clinically meaningful and aligns with the journal’s scope. The within-subject vignette design is conceptually strong, and the reported findings are potentially impactful.**

**However, there are multiple high-severity methodological and analytic concerns that threaten the validity of the central conclusions, not just presentation or clarity issues.**

**Key Determinants (Decision-Driving Issues)**

**1. Measurement validity is currently unclear (critical flaw)**

- **The AQ instrument is mischaracterized and/or incorrectly scored, with internal inconsistencies between AQ-27 vs AQ-9 structure, Likert scaling, and composite ranges.review_57**
- **This directly affects all primary outcomes, meaning results cannot be trusted without re-specification and likely re-analysis.**

**➡️ This alone warrants major revision, potentially full reanalysis.**

**2. Regression model contains a structural impossibility**

- **“Prior education” is listed as 100% present (no variance) but included as a statistically significant predictor.review_57+1**
- **A zero-variance predictor cannot produce a coefficient, indicating:**
  - **Data coding error, or**
  - **Table/reporting error, or**
  - **Model mis-specification**

**➡️ This is a fatal internal inconsistency that must be resolved before interpretation.**

**3. Vignette confounding undermines the main causal/interpretive claim**

- **The epilepsy and FS vignettes differ across multiple clinically meaningful dimensions (duration, semiology, awareness, EEG, etc.), not just diagnostic label.review_57+1**
- **Therefore:**
  - **Differences in “stigma” domains (e.g., dangerousness, fear, responsibility) cannot be attributed to diagnosis alone.**

**➡️ This does not invalidate the study entirely, but it forces a major reframing of conclusions.**

**4. Statistical rigor is insufficient**

- **No multiple-comparison correction across 9 AQ domains.peer_review_manuscript_57**
- **Suspicious uniform p-value reporting (all 0.001).peer_review_manuscript_57**
- **Overfitted regression relative to sample size (n=75, multiple predictors).peer_review_manuscript_57**
- **No clearly defined primary endpoint, and unclear composite score validity.review_57**

**➡️ These issues collectively raise risk of Type I error and model instability.**

**5. Over-interpretation of findings**

- **Claims of:**
  - **“Validation” of patient experiences**
  - **“Protective” effects of fellowship**
  - **Prescriptive educational recommendations**
- **These exceed what can be inferred from a single-center, cross-sectional vignette survey.review_57+1**

**➡️ Requires systematic downgrading to associative and hypothesis-generating language.**

**6. Qualitative component lacks methodological transparency**

- **No description of coding framework, coders, reliability, or analytic approach.
  ➡️ Limits interpretability and reproducibility.review_57+1**

**Summary Judgment**

- **Scientific premise: Strong**
- **Design concept: Appropriate but imperfect**
- **Execution: Flawed in several key domains**
- **Data validity: Currently uncertain**
- **Interpretation: Overstated**

**👉 Therefore: Major Revisions (not Reject)
Because:**

- **The issues appear fixable with transparent reporting and reanalysis,**
- **The dataset likely remains usable,**
- **The topic is important and publishable if addressed rigorously.**

**Concerns About the Peer Reviews**

**Overall, the reviews are high quality, consistent, and appropriate. However, a few points to note as editor:**

**1. Potential overreach in statistical critique (minor concern)**

- **The reviewer invokes strict rules like 10–15 observations per predictor.peer_review_manuscript_57**
  - **While directionally correct, this is a rule of thumb, not an absolute requirement.**
  - **A more nuanced framing (e.g., penalized models, shrinkage, or exploratory analyses) could have been suggested.**

**👉 Not incorrect, but somewhat rigid framing.**

**2. Mixed stance on AQ validity**

- **The reviewer critiques:**
  - **Low Cronbach’s alpha in some domains**
  - **Use of a composite score**
- **However, these are secondary concerns relative to the more fundamental issue (instrument mis-specification).**

**👉 Slight mismatch in prioritization, but not problematic.**

**3. Very strong language around vignette confounding**

- **The vignette issue is correctly identified as major, but describing it as a “fundamental threat” that might invalidate central conclusions could be slightly overstated.**
  - **The design still supports comparison of responses to two clinically realistic scenarios, just not pure label effects.**

**👉 Should be reframed as limits causal attribution, not invalidates all findings.**

**4. Appropriate but extensive scope of critique**

- **The review is exceptionally thorough, bordering on prescriptive (e.g., literature expansions, additional variables).**
- **Some requested additions (e.g., burnout, race/ethnicity) may extend beyond the scope of the original study.**

**👉 These should be interpreted as suggestions, not requirements for acceptance.**

**Editorial Recommendation to Authors (if conveyed)**

**Focus on:**

1. **Clarifying and correcting AQ instrument and scoring**
2. **Resolving regression inconsistencies (especially prior education)**
3. **Applying appropriate statistical corrections and reporting**
4. **Reframing conclusions to align with design limitations**
5. **Expanding methods transparency (especially qualitative + sampling)**
6. **Explicitly acknowledging vignette confounding as central limitation**

**If these are addressed rigorously, this manuscript could become a publishable and meaningful contribution to Epilepsia.**

Epileptologist A’s Human-Generated Paper Decision

**Editorial Decision: Major Revisions**

**I agree with the reviewers’ consensus: this manuscript requires Major Revisions before it could be considered for publication in *Epilepsia*.**

**Rationale for Decision**

**This is a clinically important and timely topic—brief, scalable interventions for functional seizures (FS) are a major unmet need. The Re-PROGRAM concept, telehealth delivery, and high session completion rate are meaningful strengths.**

**However, as currently presented, the manuscript substantially overreaches the evidentiary limits of its design, and there are several methodological and reporting deficiencies that materially limit interpretability.**

**Key Determinants (Decision-Driving Issues)**

**1. Fundamental mismatch between design and claims (critical)**

- **The study is an uncontrolled, retrospective service evaluation, with:**
  - **No control group**
  - **No randomization**
  - **No prospective baseline seizure diary**
  - **Variable follow-up timing**
- **Yet the manuscript repeatedly implies efficacy (e.g., seizure reduction, improved functioning).review_84**

**➡️ This is the primary issue driving the decision.
The manuscript must be fully reframed as feasibility/acceptability only.**

**2. Outcome measurement is weak and potentially unreliable**

- **Primary instrument (FSPICS):**
  - **Unvalidated**
  - **Retrospective pre/post comparison**
  - **Administered a median of 229 days after interventionreview_84+1**
- **No:**
  - **Prospective seizure tracking**
  - **Standardized outcome measures**
  - **Quantification of seizure reduction magnitude**

**➡️ This creates substantial recall and measurement bias, limiting any inference about treatment effect.**

**3. Severe risk of selection and response bias**

- **Only 16/29 (55%) returned surveys, with only 12 completereview_84+1**
- **Outcomes for others:**
  - **Chart review (non-equivalent data source)**
  - **4 patients lost to follow-up**

**➡️ The reported “improvement rates” are highly sensitive to denominator choice and likely overestimated.**

**4. Participant flow and baseline characterization are inadequate**

- **No clear reporting of:**
  - **Screened vs eligible vs enrolled population**
  - **Decliners or exclusions**
- **Minimal baseline clinical data:**
  - **No seizure burden**
  - **No psychiatric comorbidity**
  - **No concurrent treatments or medication changesreview_84**

**➡️ This prevents interpretation of feasibility, selection bias, and generalizability.**

**5. Statistical analysis is insufficient**

- **Minimal inferential framework (single Fisher test of limited relevance)review_84+1**
- **No:**
  - **Effect sizes**
  - **Confidence intervals**
  - **Structured descriptive statistics of outcomes**

**➡️ Results are descriptively suggestive but analytically underdeveloped.**

**6. Qualitative methods lack rigor**

- **Thematic analysis described without:**
  - **Coding framework**
  - **Number of coders**
  - **Independence from intervention delivery**
  - **Handling of negative/neutral responsesreview_84+1**

**➡️ This weakens a component that is otherwise one of the manuscript’s strengths.**

**7. Reproducibility and reporting gaps**

- **Missing or incomplete:**
  - **Intervention materials (supplements not provided)**
  - **Fidelity monitoring**
  - **Therapist training/supervision detailsreview_84**

**➡️ Limits reproducibility and evaluation of scalability claims.**

**8. Ethics and disclosure omissions (serious)**

- **No clear:**
  - **IRB/ethics status**
  - **Consent procedures**
  - **Safety/event monitoring**
  - **Funding or conflict-of-interest disclosuresreview_84**

**➡️ These are mandatory for publication and must be addressed.**

**9. Overstated mechanistic claims**

- **Assertions about:**
  - **Brain connectivity**
  - **neuroinflammation**
  - **BDNF, autonomic regulation, etc.review_84**
- **None are measured in this study.**

**➡️ Mechanistic framing must be explicitly hypothetical, not inferential.**

**Summary Judgment**

| **Domain** | **Assessment** |
| --- | --- |
| **Clinical importance** | **High** |
| **Innovation** | **Moderate–high** |
| **Internal validity** | **Limited** |
| **Reporting rigor** | **Incomplete** |
| **Interpretability** | **Compromised by bias and measurement** |

**👉 Decision: Major Revisions (not Reject)**

**This remains potentially publishable because:**

- **The feasibility signal (100% completion) is notable**
- **The intervention concept is relevant**
- **The dataset can support a well-framed descriptive study**

**—but only if the manuscript is substantially reframed and clarified.**

**Concerns About the Peer Reviews**

**Overall, both reviews are high-quality, rigorous, and consistent. I agree with the vast majority of their assessments. A few editorial observations:**

**1. Appropriate emphasis on feasibility reframing (strength)**

**Both reviewers correctly and consistently identify that:**

- **The study must be reframed away from efficacy claims**
- **Toward feasibility and acceptability**

**👉 This is the most important and appropriate critique.**

**2. Strong (appropriate) critique of outcome measurement**

- **The concern regarding FSPICS validity and recall bias is well-justified.peer_review_manuscript_84**
- **The reviewers appropriately emphasize:**
  - **Lack of psychometric validation**
  - **Long recall interval**

**👉 This is a central methodological issue, not overemphasized.**

**3. Potential overreach: requiring psychometric validation**

- **One reviewer suggests:**
  - **Reporting full psychometric properties (Cronbach’s alpha, validity, etc.)**
- **For a retrospective service evaluation, this may be unrealistic post hoc**

**👉 More appropriate requirement:**

- **Acknowledge lack of validation**
- **Use instrument descriptively**
- **Avoid treating it as a validated outcome**

**4. Slightly rigid expectations for statistical analysis**

- **Suggestions like:**
  - **Wilcoxon testing**
  - **formal inferential comparisons**
- **In such a small, biased, retrospective dataset, purely descriptive analysis may actually be preferable.**

**👉 Not incorrect, but could risk overformalizing weak data.**

**5. Extensive scope of requested additions**

- **Requests for:**
  - **Expanded literature context**
  - **predictors of response**
  - **broader mechanistic discussions**

**👉 These are helpful but not strictly required for publication of a feasibility study.**

**6. Appropriate but strong tone on response rate**

- **The critique of 55% survey response rate as inadequate is valid in the context of:**
  - **Primary outcome measurement**

**👉 However, for real-world clinical program evaluations, this is not unusual—should be framed as:**

- **Limitation, not disqualifier.**

**Editorial Guidance to Authors (if conveyed)**

**To reach acceptability, the authors should prioritize:**

**Essential (must fix)**

1. **Remove all efficacy language**
2. **Reframe as feasibility/acceptability study**
3. **Provide full participant flow + denominators**
4. **Separate survey vs chart-review outcomes**
5. **Add ethics, consent, and disclosure statements**
6. **Clarify outcome definitions and limitations of measurement**
7. **Substantially expand qualitative methods OR reframe as descriptive feedback**

**Strongly recommended**

- **Add baseline clinical characteristics**
- **Clarify concurrent treatments**
- **Provide intervention detail/fidelity information**
- **Present results with conservative denominators (N=29)**

**Optional / lower priority**

- **Formal statistical testing**
- **Expanded mechanistic discussion**
- **Full psychometric validation (better suited for future work)**

**Bottom Line**

**This manuscript has clear potential as a feasibility study, but not as an efficacy study.**

**If the authors reframe rigorously and improve transparency, it could become a useful early-stage contribution to FS intervention literature—particularly as a precursor to their planned RCT.**

*Editorial Decisions B*

Epileptologist B’s AI-Generated Paper Decision

Decision: REJECT WITH REVIEW

Editorial rationale (200-300 words):
This manuscript reports a small, single-center, open-label randomized pilot trial of daily chicken soup versus usual diet in adults with drug-resistant focal epilepsy. The epilepsy population is relevant, and the authors appropriately report a null result without claiming efficacy. However, the manuscript does not meet Epilepsia’s threshold for scientific contribution or internal validity. The primary result is non-significant, the responder analysis is neutral, and the apparent directional difference is driven largely by worsening in the control arm rather than clinically meaningful improvement in the intervention arm. The trial is further compromised by severe differential attrition, with 7/15 discontinuations in the chicken soup arm versus 2/15 in control, analyzed using LOCF despite likely informative missingness. A randomized control participant was excluded from the percentage-change analysis because baseline seizure rate was zero, adding fragility to an already underpowered analysis.

The reviewers correctly recognized the manuscript’s transparent tone, but their major validity concerns are decisive. Reviewer 1 identified fundamental reporting and interpretive gaps: no baseline characteristics table, no withdrawal reasons, no CONSORT flow, no standardized intervention description, no confidence intervals, unexplained control-arm deterioration, and a missing reference.   Reviewer 2 similarly emphasized differential attrition, LOCF, primary-outcome instability, and overstatement of directional effects.   Although some issues are technically correctable, revision would not overcome the trial’s limited biological rationale, open-label usual-diet control, small sample, high attrition, and low international readership value. The manuscript is therefore not suitable for revision at Epilepsia.

Top decisive reasons:

1. Severe differential attrition and LOCF-based handling of likely informative dropout undermine internal validity.
2. The intervention is biologically speculative and insufficiently standardized, limiting reproducibility and interpretability.
3. The null, underpowered, open-label pilot offers limited contribution for Epilepsia’s readership beyond documenting an inadequately informative feasibility exercise.

Required actions or core reasons:

- Core design limitations are not remediable within revision: small underpowered sample, open-label usual-diet control, and severe attrition.
- Essential reporting omissions prevent adequate evaluation of randomization balance, intervention fidelity, adherence, tolerability, and missing-data mechanisms.
- The manuscript’s novelty and scientific priority are insufficient for Epilepsia given the weak rationale and inconclusive findings.

Reviewer disagreement synthesis:
Both reviewers agreed that the paper is transparent and conservatively written, and both identified differential attrition, LOCF, and overly favorable directional language as major concerns. Reviewer 1 provided a more extensive list of essential revisions and recommended major revision; Reviewer 2 was somewhat more permissive but raised the same central validity issues. The decisive editorial judgment is that these concerns reflect core limitations rather than merely fixable reporting defects.

Confidence: High

Epileptologist B’s Human-Generated Paper Decision

Decision: MAJOR REVISIONS

Editorial rationale (200-300 words):
The manuscript reports a single 52-year-old patient with IDH1 R132H-mutant, 1p/19q-codeleted grade 2 oligodendroglioma, persistent postoperative focal seizures despite levetiracetam, subsequent vorasidenib 40 mg daily, seizure freedom after one month without ASM change, and F-DOPA PET MTV reduction from 16.3 to 8.0 to 4.0 cm³ while MRI remained stable by RANO, with 8 months reported seizure freedom.   The case is relevant to Epilepsia because seizure control in IDH-mutant glioma is clinically important, and seizure outcomes were not adequately addressed in the pivotal vorasidenib setting.   The apparent first-report combination of early seizure freedom and F-DOPA PET response is novel enough to justify continued consideration as a Brief Communication. Ethical reporting is acceptable at this stage, with local ethics protocol and written consent stated.   However, the manuscript is not close to publishable as submitted. Its central clinical outcome is based on patient diary review only, without objective EEG validation, caregiver corroboration, or a robust pre-treatment seizure baseline; spontaneous fluctuation, postoperative evolution, regression to the mean, and placebo effects remain plausible.   The imaging claim also requires substantial repair: F-DOPA uptake may reflect seizure-related/peri-ictal metabolic activity, baseline seizure timing is not reported, key PET acquisition and segmentation reproducibility details are absent, and the SUVmax/MTV divergence weakens a simple tumor-response interpretation.   The reviewers were broadly supportive of the topic, but Reviewer 1’s concrete validity concerns carry more weight than the more optimistic framing of Reviewer 2. These defects are serious but potentially fixable through reporting, reframing, and interpretive restraint rather than new data collection. Therefore, major revision—not minor revision or rejection—is the appropriate threshold.

Top decisive reasons:

1. The case is timely, epilepsy-relevant, and plausibly novel, but the evidence remains a single uncontrolled observation.
2. The seizure outcome and PET-response interpretation are vulnerable to major confounding.
3. The manuscript needs substantial reporting and interpretive revision before it can meet Epilepsia’s standards.

Required actions or core reasons:

- Reframe all causal, efficacy, and “significant” claims as hypothesis-generating observations.
- Provide full PET acquisition, processing, background-reference, and MTV segmentation/reproducibility details.
- Address the seizure-activity confound in F-DOPA PET interpretation, including seizure timing before each scan and the SUVmax/MTV divergence.
- Clarify seizure outcome ascertainment, including aura freedom, duration of continuous seizure freedom, diary source/corroboration, baseline seizure pattern, and limitations from lack of EEG validation.
- Report the clinical timeline and imaging intervals clearly in months, and express MRI/PET changes as absolute and percentage changes without inappropriate statistical language.
- Discuss alternative explanations for seizure freedom and PET change, including postoperative evolution, regression to the mean, natural history, placebo effects, and medication interaction considerations.

Reviewer disagreement synthesis:
Both reviewers agreed that the topic is timely, clinically relevant, and potentially publishable after revision. Both identified the need for more cautious causal language, clearer seizure characterization, and better handling of F-DOPA PET specificity. Reviewer 1 judged the methodological and interpretive problems to require major revision, emphasizing PET confounding, missing acquisition details, lack of objective seizure validation, and incomplete timeline reporting. Reviewer 2 was more favorable but raised overlapping concerns. The decisive issues are the concrete threats to validity, not overall enthusiasm.

Confidence: High

*Editorial Decisions* C

Epileptologist C’s AI-Generated Paper Decision

Editorial Decision: Major Revisions Required

Decision Rationale (Internal)

Both reviewers independently arrived at the same conclusion: the manuscript addresses a clinically relevant and timely question — nurse-led telephone counseling targeting sleep deprivation and alcohol use as seizure triggers — and the magnitude of the reported effect is potentially meaningful. However, both raise substantively overlapping and independently identified concerns across design validity, statistical integrity, outcome ascertainment, and interpretive framing. The concerns are correctable but are too numerous and fundamental to constitute minor revisions. The work is not suitable for rejection, as the clinical premise is sound and both reviewers expressed genuine interest in seeing it published after revision. A decision of Major Revisions is therefore appropriate.

Letter to Authors

Re: Manuscript #28 — Decision: Major Revisions Required

Dear Authors,

Thank you for submitting your manuscript describing the STOP IT nurse-led telephone counseling intervention for adults with epilepsy who self-report sleep deprivation or alcohol use as seizure triggers. The topic is clinically important and well-suited to Epilepsia. Both reviewers recognized the pragmatic value of the intervention and the clinical meaningfulness of the reported effect size. However, they each independently identified overlapping and substantive concerns that must be addressed before the manuscript can be considered for acceptance. A decision of Major Revisions has therefore been rendered.

The full reviewer comments are attached. Below is a synthesis of the most critical issues requiring your attention.

I. Study Design Transparency and Control Group Definition

Both reviewers raised concerns about whether the control group was truly contemporaneous, prospectively enrolled, and assessed under equivalent conditions. The current description — controls seen "prior to implementation" or by non-participating providers — suggests a quasi-experimental or provider-level allocation design, which carries meaningful risk of secular trend, selection, and provider bias. You must:

Clarify exact enrollment periods for both groups

Confirm whether controls were consented and followed prospectively with identical procedures

Include a participant flow diagram (screening → eligibility → enrollment → allocation → follow-up → analysis)

II. Causal Language and Interpretive Framing

This is the single most consistent criticism across both reviews. The nonrandomized design, absence of an attention-matched control, and reliance on self-reported outcomes mean that the intervention group received substantially more clinical contact than controls — making this effectively a comparison of structured counseling plus attention versus standard care alone. The manuscript cannot definitively attribute seizure reduction to trigger-specific behavioral modification.

Throughout the Abstract, Key Points, Discussion, and Conclusion, language such as "effective adjunct," "significant improvements," and "clinically meaningful improvements" must be revised to associative framing (e.g., "was associated with," "observed in the context of"). Conclusions must explicitly acknowledge that a randomized controlled trial is needed before causal efficacy can be claimed.

III. Statistical Reporting Errors and Model Specification

Reviewer 1 identified a specific and important statistical inconsistency: the reported p-value for the 50% responder comparison (p = 0.008) appears inconsistent with a two-sided Fisher exact test (which yields approximately p = 0.017), and is instead consistent with a one-sided test. If two-sided testing was prespecified in the Methods, this must be corrected throughout. All p-values should be recalculated and reconciled with the stated analytic approach.

Additionally, both reviewers flagged that the multivariable logistic regression model is likely overfit, given approximately 27 total responders and at least six model terms. Please provide the full model specification, consider a more parsimonious or penalized approach, and present the adjusted analysis as exploratory.

IV. Missing Data Handling

The Methods state that patients lost to follow-up were excluded from the primary analysis and included in a last-observation-carried-forward (LOCF) sensitivity analysis. However, the Results report responder rates using denominators of 40 per group, despite documented losses to follow-up. The LOCF sensitivity analysis results are not reported.

Please provide: consistent and transparent denominators for every outcome; the results of all sensitivity analyses; and a discussion of why LOCF may be particularly problematic in this context (e.g., regression to the mean at baseline).

V. Behavioral Outcomes Definition and Mediation

The central mechanistic premise of the paper — that seizure reduction was mediated by behavioral change — is not adequately supported. Terms such as "consistent adherence to target bedtime" and "meaningful reduction or cessation of alcohol consumption" are undefined and not operationalized. Behavioral outcomes for the control group are not reported.

Please: define all behavioral outcomes quantitatively; report them for both groups; and, if mediation analysis is not feasible, explicitly refrain from implying that seizure reduction was mechanistically caused by improved sleep or alcohol behavior.

VI. Intervention Reproducibility

The manuscript proposes a scalable clinical framework, yet the intervention description lacks sufficient detail for replication — specifically, the counseling script or manual, nurse training requirements, call duration, fidelity monitoring procedures, and criteria for motivational counseling. A supplemental protocol or structured checklist should be added.

VII. Concomitant Treatment Changes

Neither ASM changes nor other epilepsy-management changes during the 12-month follow-up period are reported by group. These are plausible alternative explanations for seizure reduction and must be addressed — either with descriptive data by group, or with sensitivity analyses excluding participants who underwent major treatment changes.

VIII. Placeholders and Registration

The manuscript contains multiple unfilled placeholders (study dates, IRB protocol number, author names, ClinicalTrials.gov identifier). These must be completed before resubmission. Please also provide the trial registration date relative to enrollment start, the prespecified primary and secondary outcomes, and any protocol deviations. A completed TREND checklist should be included or cited.

Additional Issues

In addition to the above, please also address the following:

Figure 1 appears to be missing from the submitted document; only the legend is present. Please include the full figure with labeled axes, units, and sample-size denominators

Confidence intervals should be added for the primary outcome effect estimates; p-values alone are insufficient in a small study with skewed seizure-frequency data

Table 1 baseline p-values should specify the tests used and, ideally, be supplemented or replaced with standardized mean differences

Drug-resistant epilepsy should be formally defined in the Methods using a standard (e.g., ILAE) criterion

The Abstract's phrasing of "identical triggers" should be revised to reflect group-level similarity rather than individual matching

Consider whether exploratory subgroup analyses by epilepsy syndrome (particularly generalized genetic epilepsy vs. focal) are feasible, given that alcohol and sleep deprivation may have differential relevance across syndromes

Mechanistic claims in the Introduction regarding GABA-mediated tonic inhibition and cortical excitability should be verified against the cited references or softened

Summary

This manuscript presents a promising, clinically practical, and potentially impactful intervention. Both reviewers expressed genuine interest in seeing it published, and the editorial team shares that view. However, the revisions requested above are substantive and non-trivial. A revised manuscript will be subject to re-review. We encourage you to respond to each point systematically in a detailed response-to-reviewers document.

We look forward to receiving your revision.

Sincerely,

The Editors, Epilepsia

Epileptologist C’s Human-Generated Paper Decision

**Editorial Decision: Major Revisions**

**Journal: *Epilepsia***
**Manuscript Reference:** Article 31
**Re:** Long-term outcomes of seizures induced during the "Pocket Monster" television broadcast

Dear Authors,

Thank you for submitting your manuscript to *Epilepsia*. Your work addresses a historically significant and clinically relevant topic — the long-term trajectories of patients who experienced visually induced seizures during the 1997 "Pocket Monster" broadcast. Two expert reviewers with relevant expertise have now evaluated this submission. Both reviewers acknowledge the descriptive value of the dataset and the clinical interest of the 5-year follow-up comparison between patients with and without pre-existing epilepsy. However, both have identified substantial concerns that must be fully addressed before the manuscript can be considered further for publication. On this basis, I am returning the manuscript with a decision of **Major Revisions Required**.

Please be advised that the scope and number of concerns raised are extensive, and the revised manuscript will undergo re-review. I draw your attention in particular to the following priority issues:

**Priority Issue 1: Originality and Potential Redundant Publication *(Reviewer 2 — Critical)***

This is the most serious concern raised and must be addressed transparently before all others. Reviewer 2 notes that the manuscript does not clearly distinguish its contribution from the previously published 5-year follow-up reports (your references 7 and 8), which appear to use the same 91-patient cohort and the same 25 recurrent cases. *Epilepsia* places high importance on scientific novelty and publication integrity. You must explicitly state in your revision whether this constitutes a secondary analysis of a previously published cohort, identify which data and analyses are entirely new, describe any overlap in tables, results, or text with prior publications, and articulate the distinct scientific question this paper addresses. Without a clear and satisfactory response to this point, the manuscript cannot proceed to further review.

**Priority Issue 2: Critical Internal Inconsistency in Table 1 *(Reviewer 2 — Critical)***

Reviewer 2 identifies a likely reversal of the LRE/GE counts in the "Epilepsy syndrome" row of Table 1 — a finding that is central to the manuscript's conclusions. As written, the table appears to report the opposite direction of the association described in the Results, Discussion, Abstract, and Conclusions. This must be thoroughly verified and corrected, with all downstream text and the p-value recalculated accordingly.

**Priority Issue 3: Incomplete Methods and Unresolved Placeholders *(Reviewer 2 — Major)***

The manuscript contains unreplaced placeholders ("XX/XXXX," "XXXX," "XXX") for dates and locations, lacks a participant flow diagram, does not explain the 12 patients without follow-up data, and does not clearly define inclusion/exclusion criteria or ascertainment procedures. These omissions are not acceptable in a manuscript submitted for peer review and must be fully resolved.

**Priority Issue 4: Unexplained Corrections to Previously Published Data *(Reviewer 2 — Major)***

The statement that "several corrections were made on the patients' data from the original study" is not explained. The nature, number, and adjudication of these corrections must be specified, as they may affect group assignment, syndrome classification, or recurrence status.

**Priority Issue 5: Selection Bias Must Be Foregrounded *(Both Reviewers — Major)***

Both reviewers emphasize that restricting the analysis to the 25 patients with seizure recurrence — while omitting the 66 who did not recur — constitutes a major design limitation that is not adequately communicated. This selection must be prominently stated in the Abstract, Methods, Results, and Discussion, and all claims about "patients with visually induced seizures" must be reframed to refer only to the recurrent subset.

**Priority Issue 6: Overstatement of Conclusions, Particularly Regarding Treatment *(Both Reviewers — Major)***

Both reviewers flag that the conclusion that results "will be useful in order to determine the treatment" of patients with visually induced seizures is not supported by a study that includes only recurrent cases, does not compare treatment strategies, and does not assess treatment outcomes. This claim and similar overstatements must be substantially qualified or removed.

**Additional Major Issues to Address:**

- **Small sample and statistical approach:** With n=25 (12 vs. 13), multiple unadjusted comparisons are statistically unreliable. Both reviewers strongly recommend reframing the manuscript as a descriptive, hypothesis-generating study, reporting effect sizes and confidence intervals, and de-emphasizing p-values. (*Both Reviewers*)
- **Non-standardized EEG and photosensitivity assessment:** IPS protocols varied across hospitals, undermining comparability of PPR rates — a central construct. Conclusions regarding photosensitivity must be softened, and protocol variation must be described and prominently acknowledged. (*Both Reviewers*)
- **Outdated classification systems:** The use of 1981 and 1989 ILAE classification frameworks should be explicitly justified and, where feasible, mapped to current ILAE terminology. (*Both Reviewers*)
- **Speculative mechanistic interpretation:** The Discussion's suggestions that the Pocket Monster broadcast "unmasked latent epilepsy" or "coincided with active epileptogenicity" are counterfactual claims the study design cannot support. These must be recast as hypotheses. (*Both Reviewers*)
- **Confounding by treatment and age:** Near-universal AED exposure in the Epilepsy group versus minimal exposure in the Nonepilepsy group is a major confounder that is not addressed. Either limited adjusted analyses should be performed, or the limitation must be explicitly acknowledged with causal language removed. (*Reviewer 2*)
- **Ethics statement:** No IRB/ethics committee approval statement or consent/waiver description is present. This is mandatory for *Epilepsia* and must be added. (*Reviewer 2*)
- **Syndrome classification reproducibility:** Diagnostic criteria, timing of syndrome assignment, and whether central adjudication was performed must be specified. (*Reviewer 2*)

**Minor Issues to Address:**

- Restructure the Abstract to properly describe design, cohort, and comparison groups, reserving results for the Results section.
- Replace "mental retardation" with "intellectual disability" throughout.
- Relabel "Nonepilepsy group" to "no prior epilepsy group" or equivalent to prevent misreading.
- Provide complete denominators and missing-data counts for all rows in Table 1.
- Justify and/or present continuous data for the age <12 years cutoff.
- Define AED exposure categories precisely (new, continued, adjusted).
- Add a dedicated Limitations paragraph to the Discussion.
- Update and broaden the reference list to include contemporary literature on photosensitive epilepsy and current classification frameworks.
- Revise imprecise or grammatically unclear sentences throughout.

**Editorial Summary:**

This dataset has genuine descriptive and historical value for the field of photosensitive epilepsy. However, in its current form the manuscript has critical issues regarding potential redundant publication, an apparent reversal of a central result in Table 1, incomplete methods, overextended statistical inference, and conclusions that exceed what the data can support. The revision must be substantive. A manuscript reframed as a clearly labeled descriptive secondary analysis — with transparent acknowledgment of its limitations, corrected tables, resolved originality concerns, and appropriately tempered conclusions — has the potential to make a useful contribution to *Epilepsia*.

Please submit your point-by-point response to all reviewer and editor comments along with your revised manuscript.

Sincerely,
**The Editors, *Epilepsia***
